# Global pricing of AWaRe (Access, Watch, Reserve) antibiotics: implications of the UNGA-AMR 70% Access target on national pharmaceutical expenditure

**DOI:** 10.64898/2026.02.12.26346187

**Authors:** Kasim Allel, Filip Djukic, Mike Thorn, Aislinn Cook, Peter Stephens, Suzannah Chapman, Anand Balachandran, Michele Cecchini, Elizabeth Tayler, Jennifer Cohn, Alexandra Cameron, Benedikt Huttner, Michael Sharland, Koen B. Pouwels

**Author notes:** Corresponding author: Radcliffe Primary Care Building, Radcliffe Observatory Quarter, Woodstock Rd, Oxford OX2 6GG. These authors contributed equally (Michael Sharland, Koen B. Pouwels).

## Abstract

**Background:** The United Nations General Assembly High-level Meeting on Antimicrobial Resistance (UNGA HLM-AMR) committed to a target that 70% of global human antibiotic use (ABU) should be from the Access group of the WHO AWaRe system.

**Methods:** We used 2019 IQVIA MIDAS® global ABU Quarterly value sales, volumes (kg/SU) and average ex-manufacturer prices to evaluate price per daily defined dose (DDD) by AWaRe group across countries. IQVIA MIDAS volumes/value data reflect public, private, or mixed sectors. We estimated potential national pharmaceutical expenditure savings if i) the UNGA 70% Access target was met, and ii) national ABU aligned with the WHO Model List of Essential Medicines (EML). We evaluated 7-day treatment prices for common oral and parenteral antibiotics across AWaRe groups. We measured affordability in middle-income countries (MICs) by income group, as the percentage of the population at risk of falling below national poverty lines if paying out-of-pocket, using income distributions and generalised beta distributions of the second kind. Prices were reported in 2019 international dollars (I$).

**Results:** Volume-weighted ex-manufacturer prices per DDD were lower for Access (I$1·2, IQR I$0·7) than Watch (I$2·6, IQR I$2·1) and highest (I$83·8, IQR I$80·9) for Reserve antibiotics. Lower prices were seen in high-income countries for Access antibiotics. Meeting the 70% Access target could save countries I$0·1 million–I$4·9 billion annually. Global savings could reach I$10·4 billion if only WHO EML-listed antibiotics were used. Seven-day parenteral meropenem could put 7% (IQR 9%) of the population in MICs at risk of impoverishment.

**Conclusion:** Antibiotic policies focused on achieving the UNGA-AMR 70% Access target could generate significant potential national and global expenditure savings.

**Funding:** This work was supported by the Wellcome Trust (304681/Z/23/Z) as part of the Antibiotic Data to Inform Local Action (ADILA) project and the Global Antibiotic Policy initiative (GAPi) project (RES 2024-495).

## Introduction

Optimising antibiotic use (ABU) is a major focus of the Global Action Plan to combat Antimicrobial Resistance (AMR). About a third of the 186 countries (96% of World Health Organisation ‘WHO’ member states) have reported they had limited national policies for appropriate ABU, following the Tracking AMR Country Self-Assessment Survey.^1^ There is a complex balance of controlling the overuse of antibiotics combined with ensuring appropriate access to essential antibiotics. The 2022 WHO AWaRe (Access, Watch, Reserve)^2^ antibiotic book provides detailed guidance on selecting the appropriate drug, dose, and duration of the 41 antibiotics on the WHO Model Essential Medicine List (EML), treating the 35 most common infections in both the primary healthcare and hospital settings. Access antibiotics are generally narrow spectrum antibiotics with lower resistance potential and a better safety profile than broader spectrum Watch antibiotics.^2,3^ Due to the continued rise in global ABU, particularly with a marked increase in Watch compared to Access antibiotics, the proportion of countries where the WHO’s 2019-2023 General Programme of Work target of 60% Access was met reduced from 76% in 2000 to 55% in 2015.^4^ A recent global study found a further 11% increase in ABU between 2016 and 2023, with projections indicating a persistent increase of up to 52% in global use by 2030 if no reductions occurred in low- and middle-income countries (LMICs).^5^ The 2024 UNGA-AMR committed to a global target that 70% of all ABU in humans should be Access by 2030,^6^ reflecting that over 90% of antibiotics are used in primary care where most infections can still be effectively treated with Access antibiotics.

Despite a projected $412 billion annual loss in healthcare costs due to AMR by 2030,^7^ there is limited literature on the potential economic impacts of optimising ABU.^3,8,9^ We hypothesised that Watch antibiotics are generally more expensive than Access antibiotics and are frequently over used, especially in primary care. Therefore, if countries move closer to achieving the UNGA target by encouraging the use of Access antibiotics where appropriate in place of Watch, this could result in substantial pharmaceutical expenditure savings. Additional savings, while also improving equitable access to essential medicines could come from promoting the use of antibiotics included in the EML and implementing external/international reference pricing policies that align national procurement prices with regional median prices.^3,8^ Reference pricing is particularly relevant for pricing single-source, on-patent medicines, and less so for multisource, off-patent products.

## Methods

### Study design and selection of main data source

We conducted a retrospective comparative study on the global pricing of 267 antibiotics using the WHO AWaRe system, including the 41 antibiotics on the 2023 WHO Model Essential Medicine List (EML).^10^ Following a systematic search of existing data sets, we selected IQVIA MIDAS Quarterly sales data for 2019 for its depth and broader country coverage. IQVIA MIDAShas been used in other global pharmaceutical pricing studies.^11-16^ IQVIA’s national audits and MIDAS reflect local industry-standard pack prices, which may represent either list prices or average invoice prices, depending on country-specific reporting practices. These do not capture net prices realised by manufacturers. Sales values in IQVIA audits are derived by applying reported prices to product volume data. To standardise comparisons, IQVIA MIDASadjusts national audit sales values using a single average industry margin. Actual margins, which may vary by product, are not disclosed. Furthermore, clawbacks, discounts, and rebates remain unknown, while public-private sector representativeness vary by country, potentially leading to an overestimation of prices (Text A1).

We sourced 2019 IQVIA MIDASQuarterly data, the last full year prior to COVID-19, covering oral (e.g., capsules, liquids and tablets) and intravenous (e.g., injections and solutions) antibiotics. This dataset provides national pricing and uses information for antibiotics across 73 countries, representing all WHO^17^ regions and World Bank^18^ income groups, except for low-income countries (Table A1, Figure A1).

### Main variable: Prices for AWaRe antibiotic by DDD

We calculated the country-level ex-manufacturer price per DDD sold into the retail-hospital sector for all available antibiotics following a two-tier approach. Where actual ex-manufacturer prices were not available, IQVIA MIDASapproximates them by adjusting retail or wholesale prices using estimated mark-up factors based on local pricing structures. Initially, we calculated total DDDs per antibiotic package or product. Subsequently, we calculated ABU (in units of DDD, derived from the originally reported volume in kg/SU) for a given package and then divided the ex-manufacturer price for that package by the calculated ABU to derive a price per DDD unit. See more details in Supplementary appendix Text A2 and Table A2 for price specification by country.

### Antibiotic price verification and cross-referencing

We used the WHO Health Action International project on medicine prices and availability,^19^ which provides the most recently available multiple LMICs data on the lowest country-level prices from 2004 to 2011. We focused on the most consistently reported antibiotics with no missing data (i.e., amoxicillin and ciprofloxacin) across 10 middle-income countries ‘MICs’ (Text A4). We also compared IQVIA MIDAS’s volume-weighted prices per DDD of commonly prescribed AWaRe antibiotics (amoxicillin, azithromycin, ceftriaxone, ciprofloxacin, clarithromycin, gentamicin, meropenem, linezolid, and colistin) against country-specific sources, documented in the WHO’s Medicines Prices and Market Information Sources database,^20^ where country information were available (N=8 countries; Text A5). A volume-weighted price is the average price of a medicine adjusted for the volume purchased, providing greater weight to prices from larger procurement volumes. All respective prices were inflated and converted to 2019 I$. Results are presented in Text A4-5.

### Potential national pharmaceutical expenditure savings of implementing the 70% Access target

ABU was measured in total DDDs per antibiotic and country. Subsequently, we estimated volume-weighted ex-manufacturer prices per DDD across different country-antibiotic combinations based on the WHO’s AWaRe system (Figures A2-4).^10^ We quantified median ex-manufacturer prices per DDD by country, WHO region^17^ and income group.^18^ We also calculated the percentage of ABU by AWaRe group at a country level to estimate changes if they implemented varying levels of ABU targets. We estimated a range of 60%, 65%, 70%, 75%, and 80% of national antibiotic use being Access antibiotics. This was then multiplied by the difference in ex-manufacturer prices between Access and Watch antibiotics (assuming total antibiotic use was the same), to estimate potential savings (total and per capita). We then created a tool for visualisation of a country’s potential pharmaceutical expenditure savings to account for different pricing strategies and improvement in ABU around the 70% Access target (https://bit.ly/3WN1wZR). When available, savings were compared to national pharmaceutical expenditures per capita.^21^

### Potential national pharmaceutical expenditure savings of using WHO Model EML-listed antibiotics

Using the same approach as above, we employed a stratified analysis to estimate antibiotic prices based on inclusion in the 2023 WHO Model EML (Figure A5 and Tables A3-7).^9^ Pharmaceutical expenditure savings were calculated under the scenario where countries aligned exclusively with WHO Model (rather than national) EML-listed antibiotics.

### Potential national pharmaceutical expenditure savings of using regional reference prices

We estimated the potential savings in pharmaceutical expenditure through regional referencing pricing, defined here as all countries paying the same price as the median ex-manufacturer price for Access and Watch antibiotics within their specific WHO region.

### Potential patient level expenditure impact of AWaRe antibiotic treatment

To assess whether patients could afford a full treatment course for minor and severe infections when paying out-of-pocket for antibiotics, we assessed the price and affordability of (Scenario 1) oral primary care treatment of the most common infections, and (Scenario 2) systemic antibiotic hospital treatment for sepsis of unknown origin across country populations, following the WHO AWaRe antibiotic book guidance.^2,22^

Scenario 1: We quantified 7-day hypothetical monotherapy treatment prices for oral AWaRe book Access (amoxicillin, amoxicillin + clavulanic acid, cefalexin, doxycycline, nitrofurantoin, phenoxymethylpenicillin), Watch (azithromycin, cefixime, ciprofloxacin, clindamycin), and Reserve (linezolid) antibiotics for most common bacterial infections (Text A6). We employed a conservative approach, pricing single-agent courses using ex-manufacturer prices.

Scenario 2: We estimated the empirical treatment price of the AWaRe book antibiotics recommended for the treatment of sepsis for adults and neonates (Text A7), generating for adults a 4-tiered (ceftriaxone + amikacin/gentamicin, piperacillin + tazobactam, meropenem, and colistin) and for neonates a 5-tiered group analysis (ampicillin/benzylpenicillin + gentamicin [Access], ceftriaxone [Watch], piperacillin + tazobactam [Watch], meropenem [Watch], and colistin [Reserve]).

We subsequently estimated affordability of Scenario 1 and 2 using estimated national income distributions generated by fitting generalised beta distributions of the second kind^23^ utilising 2019 country gross national income (GNI) per capita, Gini index, and income shares in deciles from the world income inequality database.^24^ Building upon income distributions, we estimated the proportions of country population, based on annual income distributions, that could be at risk of impoverishment – using national poverty lines – if they were to pay for the these antibiotics out-of-pocket. We focused on MICs only, assuming that, in most HICs, antibiotic prices are largely covered by health systems.

All prices were expressed in purchasing power parity ‘PPP’-adjusted 2019 international dollars (I$), utilising 2019’s OECD exchange^25^ and PPP^26^ rates. All analyses were performed in R software v2022. The script utilised is available on GitHub (https://bit.ly/3LF24L0).

## Results

### Prices of AWaRe antibiotic by DDD

Volume-weighted ex-manufacturer median prices by DDD were I$1·2 (p25^th^ I$1·0, p75^th^ I$1·7) for Access, I$2·6 (p25^th^ I$1·8, p75^th^ I$3·9) for Watch, and I$83·8 (p25^th^ I$52·6, p75^th^ I$133·6) for Reserve antibiotics (Table 1). Median Access prices were lower for HICs (n=40 countries) and upper middle-income countries (UMICs, n=20), I$1·1 and I$1·3, respectively, compared to I$1·5 in lower middle-income countries (LMC, n=13). Southeast Asia (n=5 countries) and the Western Pacific (n=11) had the lowest median prices for Access antibiotics (I$1·1 and I$1·1). Among Watch antibiotics, the Eastern Mediterranean (n=9 countries) and Western Pacific regions (n=11) reported the highest median prices (I$3·5 and I$3·6, respectively), while Southeast Asia the lowest (I$1·4) (Table 1, Figure A6). Among price variability explained by volume and antibiotic type, on average, 65% was attributed to procured volume and 35% to molecule differences (Text A8). New Zealand had among the lowest prices for Access and Watch (Figure 1, Table A8). The differences were due to procured volumes, antibiotic-mix, and infectious disease burdens. Eight countries had Access-to-Watch price ratios ≥1(Figure A7 and Table A9). Overall, the most expensive antibiotics in each AWaRe group were ampicillin sulbactam, cefazedone, mecillinam (Access), fidaxomicin, temocillin (Watch), and tigecycline, daptomycin (Reserve) (Table A10), with marked heterogeneity in pricing noted between HICs and MICs across most antibiotics, especially for Reserve (Figure 2). Parenteral antibiotics were generally more expensive compared to oral (Figure A8 and Table A11-2). Prices per DDD for oral and parenteral were I$1·0 (IQR I$0·6) and I$8·4 (IQR I$6·9) for Access, I$1·5 (IQR I$0·9) and I$21·7 (IQR I$21·7) for Watch, and I$68·1 (IQR I$115·8) and I$121·4 (IQR I$84·7) for Reserve, respectively, emphasising the significant potential pharmaceutical expenditure savings associated with step down to oral antibiotics. Country-aggregated antibiotic expenditure per capita was highest in France, Saudi Arabia and the US (I$17·6-I$152·9) and lowest in Australia, India, Peru, South Africa, Venezuela, and Vietnam (below I$7·9) (Figure A9, panel A).

**Table 1.** Volume-weighted ex-manufacturer price (I$) per DDD across AWaRe groups, by WHO-region and income group (n=73 countries using IQVIA MIDAS Quarterly sales data for the year 2019)

| AWaRe category/<br>Region or<br>income groups | Sampled<br>population-<br>weighted GDP<br>per capita<sup>†</sup> | Overall<br>population-<br>weighted GDP<br>per capita<sup>†</sup> | A. Access antibiotics |  |  |  | B. Watch antibiotics |  |  |  | C. Reserve antibiotics |  |  |  |
| --- | --- | --- | --- | --- | --- | --- | --- | --- | --- | --- | --- | --- | --- | --- |
|  |  |  | Median | p25<sup>th</sup> | p75<sup>th</sup> | IQR | Median | p25<sup>th</sup> | p75<sup>th</sup> | IQR | Median | p25<sup>th</sup> | p75<sup>th</sup> | IQR |
| All | 21,104 | 17,963 | 1.2 | 1.0 | 1.7 | 0.7 | 2.6 | 1.8 | 3.9 | 2.1 | 83.8 | 52.6 | 133.6 | 80.9 |
| **WHO Region** |  |  |  |  |  |  |  |  |  |  |  |  |  |  |
| AFR, n=2 | 13,627 | 4,487 | 1.1 | 0.9 | 1.3 | 0.4 | 3.2 | 3.1 | 3.2 | 0.1 | 45.9 | 45.9 | 45.9 | 45.9 |
| AMR, n=13 | 36,390 | 34,231 | 1.3 | 1.1 | 2.1 | 0.9 | 2.2 | 1.2 | 4.8 | 3.6 | 117.1 | 64.6 | 194.4 | 129.8 |
| EMR, n=9 | 13,238 | 11,838 | 1.6 | 1.3 | 1.9 | 0.7 | 3.5 | 3.1 | 3.9 | 0.9 | 143.2 | 36.3 | 245.0 | 208.7 |
| EUR, n=33 | 42,490 | 38,432 | 1.2 | 1.0 | 1.5 | 0.5 | 3.1 | 2.2 | 3.7 | 1.5 | 91.4 | 62.3 | 133.8 | 71.4 |
| SEAR, n=5 | 8,299 | 8,187 | 1.1 | 0.9 | 1.2 | 0.3 | 1.4 | 1.1 | 4.6 | 3.5 | 21.6 | 7.1 | 107.8 | 100.7 |
| WPR, n=11 | 20,109 | 19,844 | 1.1 | 0.8 | 1.5 | 0.8 | 3.6 | 2.6 | 4.9 | 2.4 | 101.4 | 84.6 | 148.5 | 63.9 |
| **Income group** |  |  |  |  |  |  |  |  |  |  |  |  |  |  |
| HIC, n=40 | 51,076 | 50,844 | 1.1 | 0.9 | 1.4 | 0.5 | 3.1 | 2.3 | 4.7 | 2.4 | 100.8 | 69.5 | 152.3 | 82.8 |
| UMC, n=20 | 18,096 | 17,001 | 1.3 | 1.1 | 1.8 | 0.7 | 2.9 | 2.0 | 4.2 | 2.2 | 91.4 | 48.6 | 147.3 | 98.7 |
| LMC, n=13 | 7,760 | 6,976 | 1.5 | 1.2 | 1.6 | 0.4 | 2.9 | 1.5 | 3.6 | 2.1 | 96.6 | 7.1 | 165.1 | 157.9 |

Notes: WHO= World Health Organization. AFRO= African region, AMRO= Americas region, EMRO= East Mediterranean region, EURO= Europe, SEARO= Southeast Asia region, WPRO= Western-pacific region. HIC= High-income country, UMC= Upper middle-income country. LMC= Lower middle-income country. p25<sup>th</sup>= percentile 25<sup>th</sup>, p75<sup>th</sup>= percentile 75<sup>th</sup>. I\$ International dollars, 2019. n= number of observations/countries. GDP= Gross Domestic Product. <sup>†</sup> 2019 GDP per capita PPP (I\$) was weighted to each subgroup (all, WHO region, and Income groups) following population sizes of sampled countries (n=73) and overall (all countries worldwide) for comparison purposes; Data were extracted from the World Bank (https://data.worldbank.org). Source: Authors' analysis of IQVIA MIDAS Quarterly Sales data for the year 2019, reflecting estimates of real-world activity. Copyright IQVIA. All Rights Reserved.

**Figure 1.**
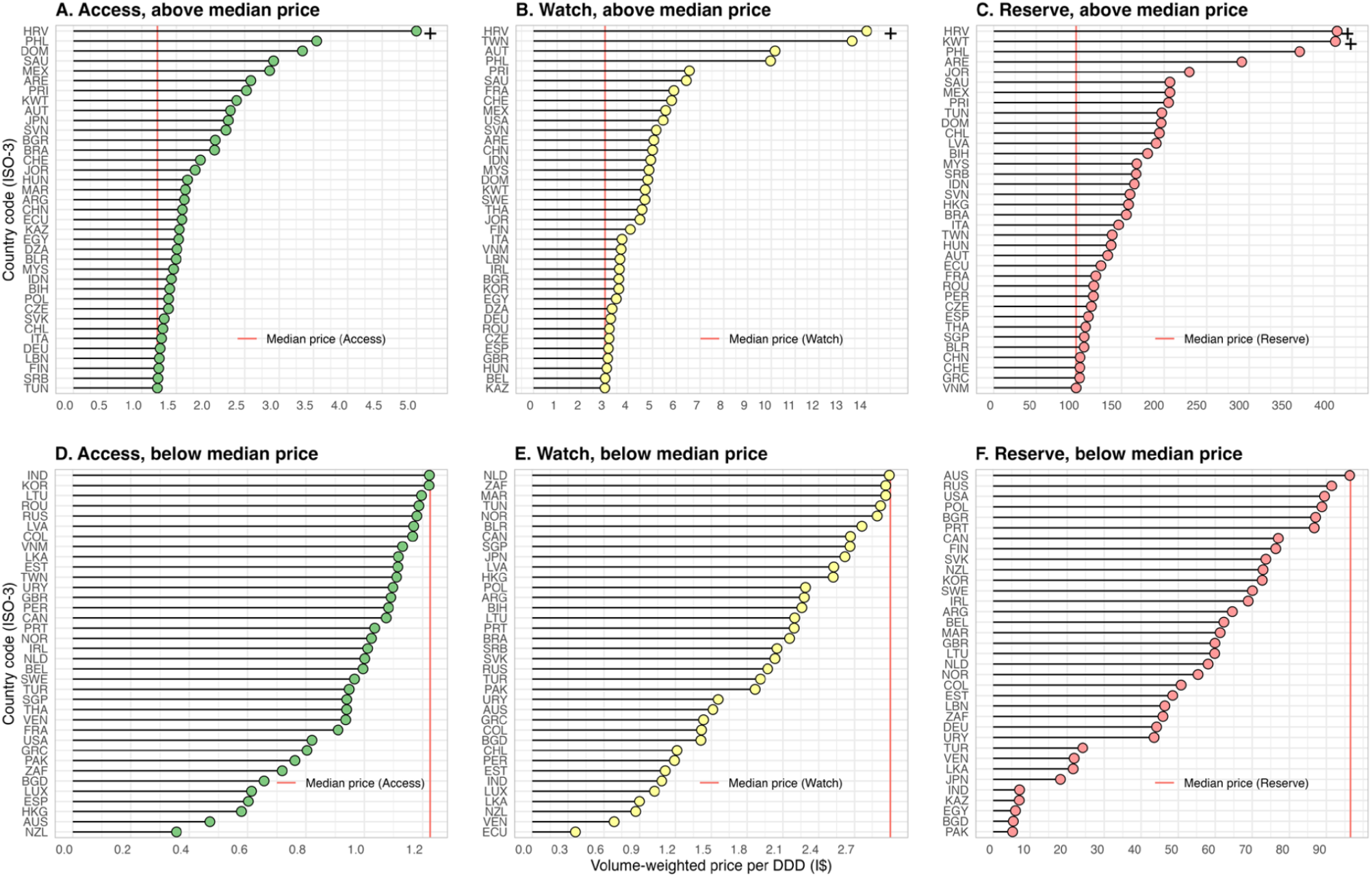
Volume-weighted ex-manufacturer prices (I$) per DDD across AWaRe groups, by country (n=73 countries using IQVIA MIDAS Quarterly sales data for the year 2019) Notes: DDD= Daily defined doses. I$= International dollars. Panel A, +HRV was I$11. Panel B, +HRV was I$30. Panel C, +HRV was I$1479 and +KWT was I$1257 per DDD. I$= International dollars, 2019. Country ISO-3 code come from the International Standard for country codes, see Supplementary Table A1 for full list. Source: Authors’ analysis of IQVIA MIDAS Quarterly Sales data for the year 2019, reflecting estimates of real-world activity. Copyright IQVIA. All Rights Reserved.

**Figure 2.**
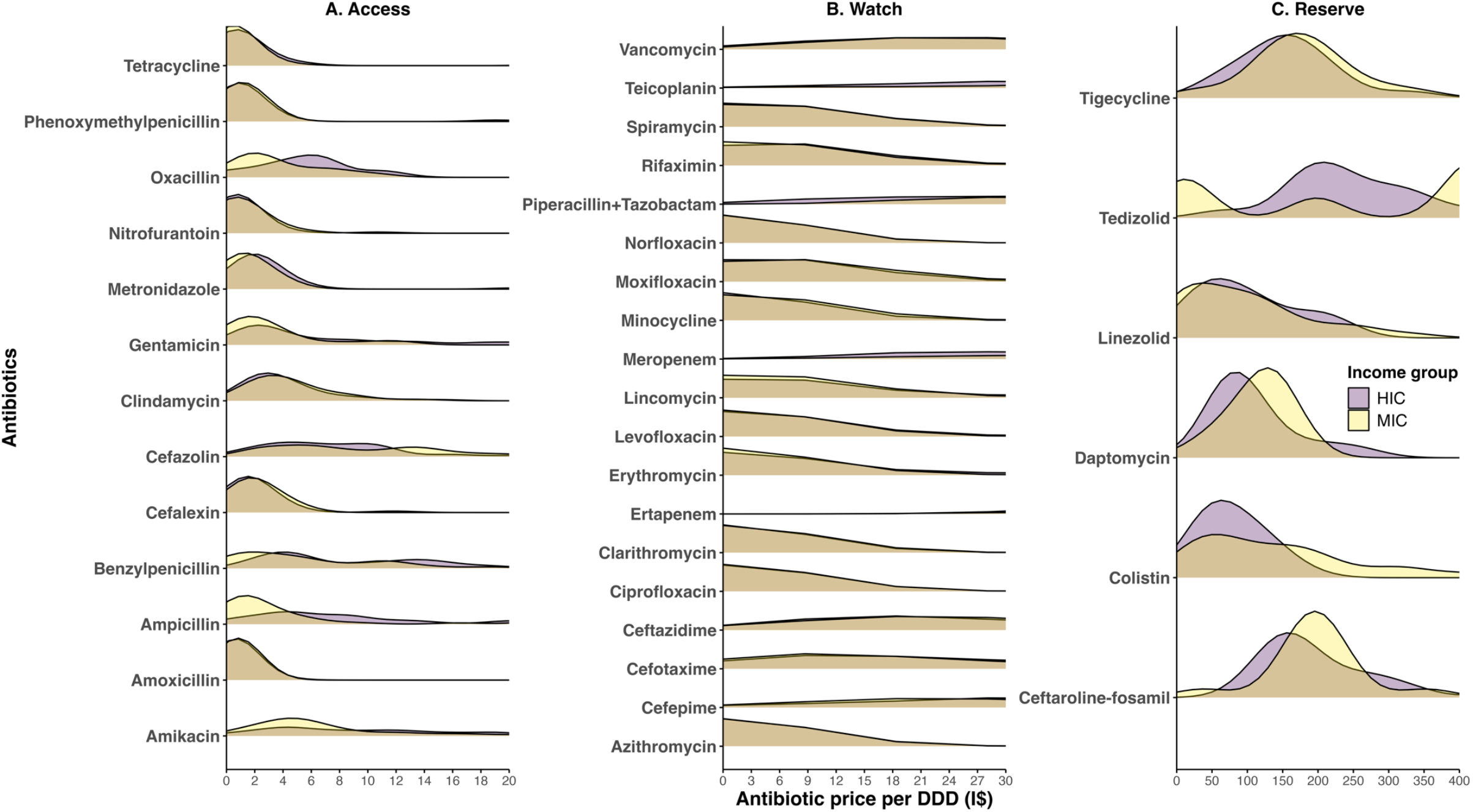
Distribution of volume-weighted ex-manufacturer price (I$) per DDD across leading antibiotics, segmented by AWaRe group and income level using IQVIA MIDAS Quarterly sales data for the year 2019 Notes: AWaRe= Access, Watch and Reserve. HIC= High-income country. MIC= Middle income countries. DDD= Daily defined doses. I$= International dollars, 2019. Source: Authors’ analysis of IQVIA MIDASQuarterly Sales data for the year 2019, reflecting estimates of real-world activity. Copyright IQVIA. All Rights Reserved.

### Potential pharmaceutical expenditure savings from achieving the 70% Access target

Access antibiotics had a median percentage of total ABU of 50% (IQR 26%) across the 73 countries included (Figure 3, panel A), highest in Africa (74%) and lowest in Southeast Asia (36%). Supplementary Text A9 provides specific analyses for ABU by country and AWaRe groups, with median use of 46% for Watch and 0·1% for Reserve antibiotics, the remaining share was for not recommended. Achieving higher national Access ABU use could deliver substantial financial savings, if Watch ABU is replaced by Access (Tables A13-4). For countries with Access ABU, as a percentage, below the 70% threshold and with higher ex-manufacturer-based antibiotic prices for Watch compared to Access (n=52, 71% of the 73 originally included countries), achieving a 70% Access ABU target could save, on average, I$1·6 per capita annually. Total savings could comprise I$8·4 billion annually across these 52 countries. We observed savings per capita ranging from I$4·5 to I$5·0 in Saudi Arabia, Italy, Korea, and Puerto Rico (see Figure 3B). An average saving of I$161·9 million per country (IQR I$111·5 million, median I$16·6 million) was identified, with China leading at I$4·9 billion. These savings would be equivalent to approximately 1% of total pharmaceutical health expenditures per capita in some countries (e.g., Italy, Romania, Korea) (Figure A10, Table A15).

**Figure 3.**
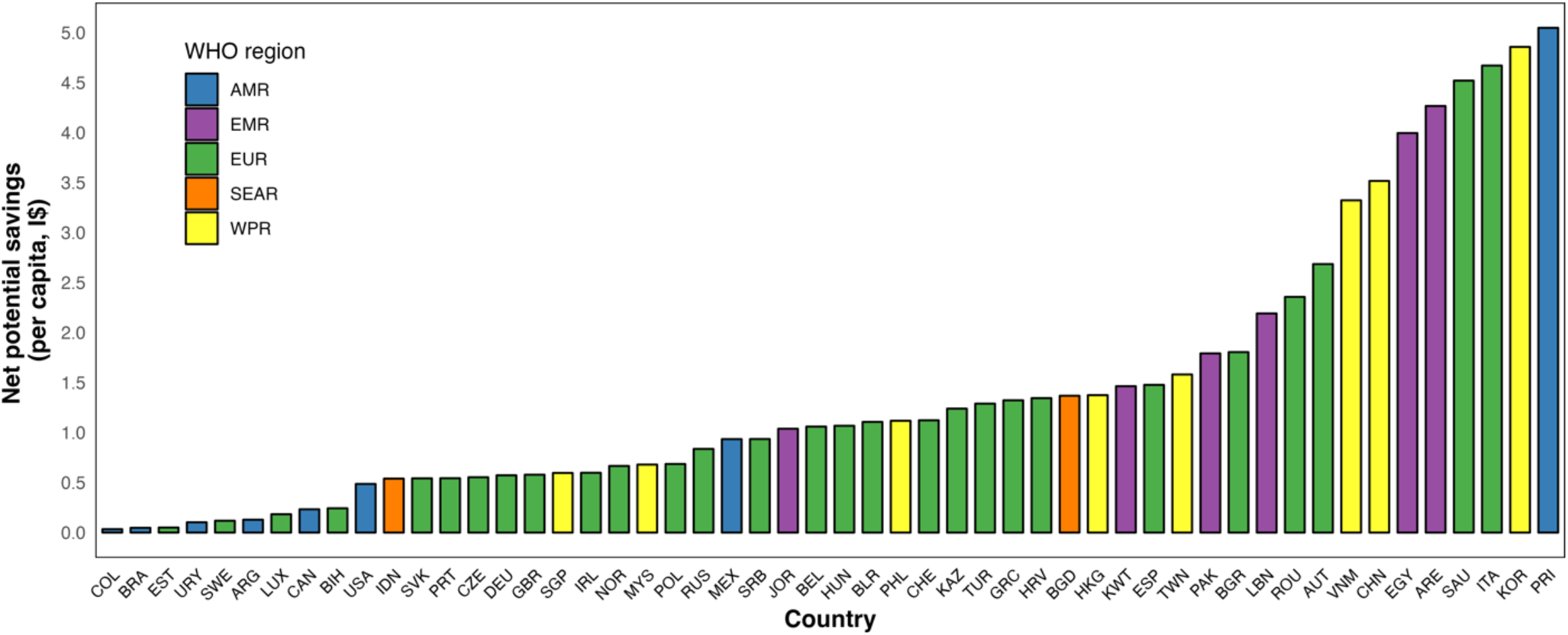
Net country level potential savings in antibiotic expenditure if the proportion of Access antibiotics reach the 70% target following antibiotic ex-manufacturer prices per DDD using IQVIA MIDAS Quarterly sales data for the year 2019 Notes: The percentage of volume-weighted antibiotics sold by AWaRe category (shares) can be found in Supplementary Text A10, Figure A10.1. WHO= World Health Organization. AFRO= African region, AMRO= Americas region, EMRO= East Mediterranean region, EURO= Europe, SEARO= Southeast Asia region, WPRO= Western-pacific region. UN-GA= United Nations General Assembly. AWaRe= Access, Watch and Reserve. I$= International dollars, 2019. Country ISO-3 code come from the International Standard for country codes, see Supplementary Table A1 for full list. Source: Authors’ analysis of IQVIA MIDAS Quarterly Sales data for the year 2019, reflecting estimates of real-world activity. Copyright IQVIA. All Rights Reserved.

### Prices and pharmaceutical expenditure savings associated with the use of WHO EML-listed AWaRe antibiotics

There was marked variation in the range of EML and non-EML antibiotics used by countries (Figure A9, panel B). On average across countries, 15 Access antibiotics were listed on the EML compared to only 6 that were not, while for Watch antibiotics, the figures were 11 EML listed compared 21 non-EML. Median prices of EML-listed antibiotics per DDD were I$1·2 (IQR I$0·6) for Access, I$2·6 (IQR I$2·6) for Watch, and I$84·5 (IQR I$94·2) for Reserve groups, respectively, compared to I$2·5 (IQR I$3·4) for Access, I$2.66 (IQR I$1·8) for Watch, and I$144·1 (IQR I$110·4) for Reserve per DDD for antibiotics not on the EML (Table A16-7). The Access-to-Watch price ratio was higher for EML-listed antibiotics: 0·4 (IQR 0·3) for HICs, 0·8 (IQR 0·2) for LMCs, and 0·7 (IQR 0·4) for UMCs (Figure A11, Table A17). The Americas had higher ratios (1·1), with Chile (1·4) and Ecuador (4·3), indicating that Access antibiotics were more expensive compared to Watch in some countries. Supplementary Text A10 provides more details on EML proportions and pricing by AWaRe group.

If countries only used WHO EML-listed antibiotics and achieved the 70% Access target, the pharmaceutical expenditure savings impact would be I$10·4 billion with an average of I$196·1 million per country (IQR I$73·5 million) (Tables A18).

### Pharmaceutical expenditure savings from aligning with AWaRe antibiotic regional median prices

The number of countries with expected savings would expand from 52 to 60 if pharmaceutical expenditure savings were estimated based on using regional referencing pricing across WHO regions. The eight countries currently employing a Watch<Access pricing strategy (i.e., Chile, Dominican Republic, Ecuador, India, Japan, Peru, Sri Lanka, and Venezuela, Figure A7 and Table A9) could potentially save a total of I$1·7 billion by implementing regional reference pricing. In 60 countries, (Table A19-20), overall savings could comprise I$7·8 billion among all countries using regional reference pricing, with average pharmaceutical expenditure savings of I$1·4 per capita and country (IQR I$1·5 per capita). Ecuador, Greece and Japan could have the largest per capita savings (between I$4·2 and I$7·2). Overall country pharmaceutical expenditure savings could be I$129·6 million (IQR I$59·4 million) per country, with I$2·8 billion and I$1·0 billion for China and India, respectively (Table A21).

On average, countries could reduce total antibiotic pharmaceutical spending by 9% (IQR 8%) by meeting the 70% Access target alone, 10% (IQR 10%) by using EML-listed antibiotics alone, and 11% (IQR 11%) by using regional reference pricing alone.

### Affordability of oral antibiotic monotherapy treatment course prices

Figures A13–23 show 7-day oral antibiotic treatment prices by country under Scenario 1 (i.e., hypothetical monotherapy relative treatment expenditures across Access, Watch and Reserve antibiotics). Treatment with Access antibiotics had median prices from I$3·1 (doxycycline, e.g., second choice for community-acquired pneumonia) or amoxicillin (I$5·0, e.g., for acute otitis) to I$14·3 (amoxicillin + clavulanic acid for acute sinusitis) (Tables A22-3). Watch-antibiotic treatments ranged from I$9·5 (azithromycin) to I$96·9 (clindamycin). Linezolid, a Reserve oral antibiotic, had a median treatment course price at I$780·5 (IQR 1154·9) for mild-severe bacterial infections.

Due to their relatively high price, clindamycin and linezolid may put individuals from MICs at risk of impoverishment due to out-of-pocket payment (Figure A24, Table A24). For instance, a 7-day clindamycin regimen could potentially put an average of 0·3% of the population (IQR 0·3%) at risk of being pushed into poverty across included MICs. This translates to approximately 493,544 individuals per country, totalling 15·3 million people across 31 MICs. South Africa, Venezuela, and Brazil face significantly higher impacts, with 1·8%, 1·4%, and 1·2% of their populations, respectively, at risk. Linezolid-based oral treatments could put 3·4% (IQR 3·4%) of the populations in the included MICs at risk of being pushed into poverty. On average, 4·8 million individuals per country (median: 264,980; IQR: 1·1 million) are at risk, amounting to a total of 144.5 million people across these MICs. The Philippines (30·0%, ≈32·5 million), India (12·7%, ≈33·9 million), and Jordan (10·2%, ≈1·0 million) have the largest proportion of their population at risk, although the risk is small as linezolid in the WHO AWaRe book is only indicated as targeted treatment for severe MRSA infection or multidrug-resistant tuberculosis.

### Affordability of intravenous (IV) antibiotics and sepsis treatment prices

Under Scenario 2 (i.e., 7-day adult IV treatments for sepsis of unknown origin in adults, the median price of meropenem and colistin courses was I$759·3 and I$557·7 in HICs, compared to I$1,567·1 and I$1,076·0 in MICs (Tables A25-7), with IQRs suggesting wide price variability, particularly for meropenem. Neonatal sepsis treatment showed similar price patterns with high-priced courses for piperacillin + tazobactam and meropenem, albeit at lower overall prices (Tables A25-6 and A28).

Out-of-pocket antibiotic expenses pose a risk of impoverishment for MIC’s populations, especially for 7-day adult sepsis treatments with meropenem and colistin (Figure A25). In the Philippines 27% and 28% of the populations (i.e., 30·1 and 31·7 million inhabitants) could be at risk of impoverishment if having to pay out of pocket for colistin and meropenem-based treatments, while in Pakistan, 21% and 16% are similarly vulnerable (36·1 and 46·9 million inhabitants). An average of 7% (IQR 9%) of MIC populations are at risk of impoverishment if they would have to pay out-of-pocket for meropenem treatment of sepsis (Table A29).

Meropenem prices could put an additional 19% of Argentina’s population (i.e., 8·6 million inhabitants), and 17% each in Brazil (i.e., 37·2 million inhabitants) and Bangladesh (i.e., 28·7 million inhabitants), 12% in India (168·9 million inhabitants), and 4·3% in China (61·0 million inhabitants) at risk of impoverishment. Overall, meropenem-based treatments could put 437·9 million individuals from the included MICs at risk of impoverishment (MICs country median 0·7 million, IQR 8·4 million individuals).

## Discussion

We have conducted a comprehensive global and national analysis of AWaRe antibiotic prices. Our findings confirm that Access antibiotics are generally cheaper than Watch antibiotics, supporting the AWaRe system’s emphasis on the optimal use of Access drugs as clinically effective,^10^ lower resistance selection,^27^ and more affordable options, particularly in primary care. This analysis strengthens the case for the UNGA AWaRe target of increasing Access antibiotic use to 70%, with potential substantial national savings in antibiotic-related pharmaceutical expenditures identified with meeting this target (I$161·9 million, on average). If countries also only used EML-listed antibiotics (i.e. the antibiotics included in the WHO AWaRe book), savings could increase to an average of I$196·1 million.

Southeast Asia had the lowest ex-manufacturer prices for Access and Watch antibiotics, while MICs and Eastern Mediterranean countries reported the highest. Despite China and India’s significant manufacturing capacity (producing 70% of global antibiotics),^28^ their volume-weighted prices per DDD were not the lowest. New Zealand had the lowest prices, likely due, in part, to effective price negotiations and competitive tendering,^29^ which provides 100% coverage compared to the OECD average of 58%.^30,31^ Moreover, New Zealand’s strategic use of generics likely contributes to their lower expenditures.

Although Access antibiotics are generally less expensive than Watch antibiotics, local use patterns can reverse this trend. In eight countries, the Access-to-Watch price ratio exceeded 1, with Venezuela at 1·3 and Ecuador showing the highest at 3·5. In Ecuador, clarithromycin (Watch) made up 63% of total ABU at I$0.04 per DDD, while amoxicillin and amoxicillin + clavulanic acid (Access) accounted for 36% of Access ABU at I$0.9 and I$3.3 per DDD, respectively.^32^ These patterns suggest that higher volume use of specific antibiotics may influence procurement choices and pricing, which can also shape future potential pharmaceutical expenditure savings opportunities in the context of achieving UNGA targets.

In many LMICs, out-of-pocket antibiotic expenditure are a large portion of income, imposing significant financial strain. We found that volume-weighted ex-manufacturer prices for Access antibiotics are often higher in MICs than in HICs, with considerable price variability.^33^ However, actual patient prices may be higher due to supply chain and procurement mark-ups (e.g., pricing margins, banking fees, import duties),^34^ which limits both affordability and access. A recent report highlighted that in many LMICs, individuals pay 20-30% more for basic medicines compared to HICs.^34^ National health policies could reduce this burden by prioritising Access antibiotics use where appropriate.

Variation exists by country in the medicines that are included or excluded in WHO Model EML and their pricing.^35^ The price variation for non-EML-listed antibiotics compared to EML-listed is substantial, particularly in MICs. This highlights the economic disparities in ensuring affordable access to antibiotics to the population, which hinders progress towards universal health coverage. Aligning National EML antibiotics to the WHO EML, where appropriate and encouraging the use of EML-listed antibiotics could help to mitigate drug shortages and strengthen supply chain resilience.^36^ While the global EML serves as a benchmark for drug demands, national lists may differ to reflect local epidemiology and needs.

Our study has several limitations. We focused solely on pharmaceutical expenditure; therefore, (i) we did not account for price changes due to volume shifts, and (ii) we excluded health and economic impacts beyond drug costs, such as changes in side-effect rates or treatment failures. All IQVIA MIDASprices are estimated, which do not necessarily accurately reflect ex-manufacturer real prices. Although we retrieved ex-manufacturer prices, price volatility influences antibiotic pricing in tender-based countries (e.g., Argentina, Brazil, Italy, UK),^37^ where formal bidding processes influences pricing. Only 73 countries were included, and these are not fully representative of WHO regions (e.g., Sub-Saharan Africa is excluded) or low-income groups. Data may not represent actual patterns of ABU in MICs,^38^ as it is derived from formal distribution channels and therefore excludes important components of the market such as informal sales, donations, or dispensing through religious and faith-based organisations.^39^ However, our relative estimates of AWaRe use were consistent with the 52% Access ABU reported in the 2022 Global antimicrobial resistance and use surveillance system GLASS ABU report.^40^ The percentage of populations at risk of impoverishment in MICs may be overestimated and not fully capture affordability in countries with comprehensive or partial public coverage for medicines. However, we did not include costs of care pathways which could increase out-of-pocket expenditure. Additionally, using ex-manufacturer-based prices may underestimate end-buyer retail prices, which could be, on average, 1·0–1·3 times higher, potentially due to premiums and distribution margins, with greater values in MICs (e.g., see a conservative exploratory analysis using Pharma14 in Text A11). Actual mark-ups may be up to 8-times greater in some settings.^41^

Understanding actual affordability challenges can offer crucial leverage in price negotiations, ensuring procurement strategies are effective and aligned with local economic realities. This approach could lead to more sustainable pricing models that reflect regional economic capacities, potentially reducing costs and enhancing access to essential medicines, which is supported in our analysis of regional reference pricing. Uniform price reduction assumptions may not estimate country-level differences in molecule pharmaceutical expenditures, volumes and clinical burdens, which need to be refined in future studies at a country level.

National and global antibiotic pricing policies should focus on affordability and access, developing a “Fair price of AWaRe antibiotics” at a country level. The WHO AWaRe system is a practical tool for estimating pharmaceutical expenditures and guiding future global and national antibiotic policies, emphasising the importance of considering ABU within the broader context of medicines policy.^42^ The tool developed in this study can help inform national estimates of potential pharmaceutical expenditure savings associated with implementing the 70% Access target into national AMR action plans, reviewing the antibiotics listed on National EMLs, considering reimbursement schemes to include Access antibiotics, and where applicable and feasible, external reference pricing to reduce ex-manufacturer prices and procurement expenditures. Future steps should focus on analysing the pricing policies contained in the WHO’s pharmaceutical pricing guidelines for their applicability/suitability to antibiotics in the AWaRe EML,^8^ alongside efforts to strengthen transparent market intelligence to support efficient procurement. For instance, aligning procurement with bulk-purchase strategies can significantly reduce antibiotic expenditures, easing financial pressures on healthcare systems and improving antibiotic accessibility.^43,44^ Other strategies include pooled procurement, competitive and split tendering, and optimising antibiotic portfolios towards rational set of products could help improve price negotiations and reliable supply.

At a medicine level, optimisation of AWaRe antibiotic procurement is also critical, re-evaluating pricing data to streamline antibiotic selection, reduce unnecessary stock keeping units (e.g., brand, dosage, pack size, strength) variation, and align procurement practices with the cost-effective and clinically appropriate options. To realise this pharmaceutical expenditure potential, antibiotic pricing and use should be analysed using local data sources, to better reflect national procurement policies, while ensuring Access group AWaRe antibiotics are affordable and available at the primary-care level.^45^ Development of enhanced antibiotic data monitoring, analysis and policies, aligned with the 2024 UNGA’s global Access target could lead to local pharmaceutical expenditure savings, reducing financial burden, and promote equitable access to essential antibiotics while mitigating AMR.

## Data Availability

All data produced in the present work are contained in the manuscript and Supplementary materials.

## Author contributions

Conception and design of the study (KA, KP, MS, FD). Data acquisition: (KA, MT, FD). Data cleaning: (KA, FD, MT). Statistical analysis and methodology: (KA, KP, MT, MS). Drafted the initial manuscript: (KA). Critically revised and approved the manuscript: All authors.

## Acknowledgements

We are grateful for the collaborators of the GAPi project for their contributions, and European Society Conference of Microbiology and Infectious Diseases (ESCMID) conference 2025 version (preliminary results were presented). The findings and conclusions in this report are those of the authors and do not necessarily represent the official position of WHO or any of the institutions mentioned. The opinions expressed and arguments employed herein are solely of the authors and do not necessarily reflect the official position of the OECD or of member countries of the OECD. The statements, findings, conclusions, views and opinions contained and expressed in this research article are based in part on data obtained under license from the following IQVIA information service(s): IQVIA MIDAS.

Copyright IQVIA. All Rights Reserved. The statements, findings, conclusions, views and opinions contained and expressed herein are not necessarily those of IQVIA or any of its affiliated or subsidiary entities

## Declarations

The authors declare no conflict of interest.

