## Supplementary files for "Global pricing of AWaRe (Access, Watch, Reserve) antibiotics: implications of the UNGA-AMR 70% Access target on national pharmaceutical expenditure"

**Table A29.** Percentage of the population at risk of falling below the national poverty line due to out-of-pocket expenditure for sepsis treatment of unknown causative agent among adults, by treatment group†, data from the IQVIA MIDAS Quarterly sales data for the year 2019 . 106

**Abbreviations**

AFRO= African region
ABU= Antibiotic use
AMR= Antimicrobial resistance
AMRO= WHO Americas regional office
AWaRe= Access, Watch, Reserve
DDD= Daily defined doses
EML= Essential Medicine List
EMRO= East Mediterranean region
EURO= Europe
GA= General Assembly
GDP= Gross domestic product
GLM= Generalised linear models
HIC= High-income countries
I$= International dollars
Int$= International dollars
IQR= Interquartile range
LMIC= Low- and middle-income countries
LMC= low middle-income countries
MIC= Middle-income countries
OECD= Organization for Economic Cooperation and Development
OOPE= out-of-pocket expenditure
PPP= purchase power parity
UMC= Upper middle-income countries
UN= United Nations
USD= United States Dollars
SEARO= Southeast Asia region
SD= Standard deviation
WB= World Bank
WHO= World Health Organization
WPRO= Western-pacific region.

**Text A1.** IQVIA MIDAS details

IQVIA MIDAS® Quarterly sales data provides valuable market insights, though its ability to fully capture actual pharmaceutical revenues can be influenced by diverse national pricing mechanisms and discount structures. In several European countries, clawbacks—calculated on total country or company revenues—are excluded from IQVIA MIDAS estimates. In the US, pharmacy benefit manager (PBM) discounts remain unavailable, further limiting accuracy. Additionally, volume discounts applied at the invoice level (rather than per pack) are not accounted for in any country. Countries might be subject to different VAT schemes. Moreover, bonus stock is inconsistently reported—counted as zero in some cases and at full price in others, as seen in non-bonus stock transactions, yet not reported. Variations in national pricing mechanisms and discount structures can lead to differences between manufacturers' actual revenues and those reported in audits, offering important context for interpreting IQVIA MIDAS's representation of pharmaceutical pricing and sales.

**Table A1.** List of countries included in IQVIA MIDAS Quarterly sales data for the year 2019, by ISO3 code and WHO region and income group (n=73)

| ISO3 code | Country | WHO region | Income group |
|---|---|---|---|
| ARE | United emirates states | EMR | HIC |
| ARG | Argentina | AMR | UMC |
| AUS | Australia | WPR | HIC |
| AUT | Austria | EUR | HIC |
| BEL | Belgium | EUR | HIC |
| BGD | Bangladesh | SEAR | LMC |
| BGR | Bulgaria | EUR | UMC |
| BIH | Bosnia | EUR | UMC |
| BLR | Belarus | EUR | LMC |
| BRA | Brazil | AMR | UMC |
| CAN | Canada | AMR | HIC |
| CHE | Switzerland | EUR | HIC |
| CHL | Chile | AMR | HIC |
| CHN | China | WPR | UMC |
| COL | Colombia | AMR | UMC |
| CZE | Czech Republic | EUR | HIC |
| DEU | Germany | EUR | HIC |
| DOM | Dominican Republic | AMR | UMC |
| DZA | Algeria | AFR | LMC |
| ECU | Ecuador | AMR | LMC |
| EGY | Egypt | EMR | LMC |
| ESP | Spain | EUR | HIC |
| EST | Estonia | EUR | HIC |
| FIN | Finland | EUR | HIC |
| FRA | France | EUR | HIC |
| GBR | United Kingdom | EUR | HIC |
| GRC | Greece | EUR | HIC |
| HKG | Hong Kong | WPR | HIC |
| HRV | Croatia | EUR | HIC |
| HUN | Hungary | EUR | HIC |
| IDN | Indonesia | SEAR | UMC |
| IND | India | SEAR | LMC |
| IRL | Ireland | EUR | HIC |
| ITA | Italy | EUR | HIC |
| JOR | Jordan | EMR | LMC |
| JPN | Japan | WPR | HIC |
| KAZ | Kazakhstan | EUR | UMC |
| KOR | Korea | WPR | HIC |
| KWT | Kuwait | EMR | HIC |
| LBN | Lebanon | EMR | UMC |
| LKA | Sri Lanka | SEAR | LMC |
| LTU | Lithuania | EUR | HIC |
| LUX | Luxembourg | EUR | HIC |
| LVA | Latvia | EUR | HIC |
| MAR | Morocco | EMR | LMC |

| | | | |
|---|---|---|---|
| MEX | Mexico | AMR | UMC |
| MYS | Malaysia | WPR | HIC |
| NLD | Netherlands | EUR | HIC |
| NOR | Norway | EUR | HIC |
| NZL | New Zealand | WPR | HIC |
| PAK | Pakistan | EMR | LMC |
| PER | Peru | AMR | UMC |
| PHL | Philippines | WPR | LMC |
| POL | Poland | EUR | HIC |
| PRI | Puerto Rico | AMR | HIC |
| PRT | Portugal | EUR | HIC |
| ROU | Romania | EUR | UMC |
| RUS | Russia | EUR | UMC |
| SAU | Saudi Arabia | EMR | HIC |
| SGP | Singapore | WPR | HIC |
| SRB | Serbia | EUR | HIC |
| SVK | Slovakia | EUR | UMC |
| SVN | Slovenia | EUR | UMC |
| SWE | Sweden | EUR | HIC |
| THA | Thailand | SEAR | HIC |
| TUN | Tunisia | AFR | LMC |
| TUR | Turkey | EUR | UMC |
| TWN | Taiwan | WPR | UMC |
| URY | Uruguay | AMR | HIC |
| USA | Unites States of America | AMR | HIC |
| VEN | Venezuela | AMR | UMC |
| VNM | Vietnam | WPR | LMC |
| ZAF | South Africa | AFR | UMC |

Notes: WHO= World Health Organization. AFRO= African region, AMRO= Americas region, EMRO= East Mediterranean region, EURO= Europe, SEARO= Southeast Asia region, WPRO= Western-pacific region. HIC= High-income country, UMC= Upper middle-income country. LMC= Lower middle-income country. Authors' analysis of IQVIA MIDAS Quarterly Sales data for the year 2019, reflecting estimates of real-world activity. Copyright IQVIA. All Rights Reserved.[1] Country ISO-3 Codes from the International Standard for country codes (https://www.iso.org/iso-3166-country-codes.html).

**Figure A1.** Countries included by income-level and WHO region (n=73), data from the IQVIA MIDAS Quarterly sales data for the year 2019

A

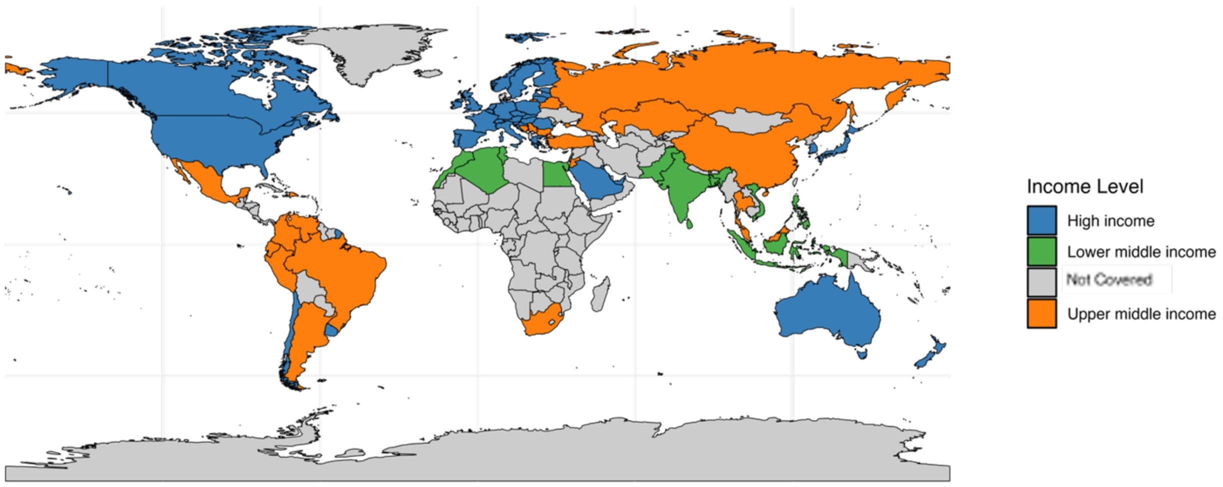

B

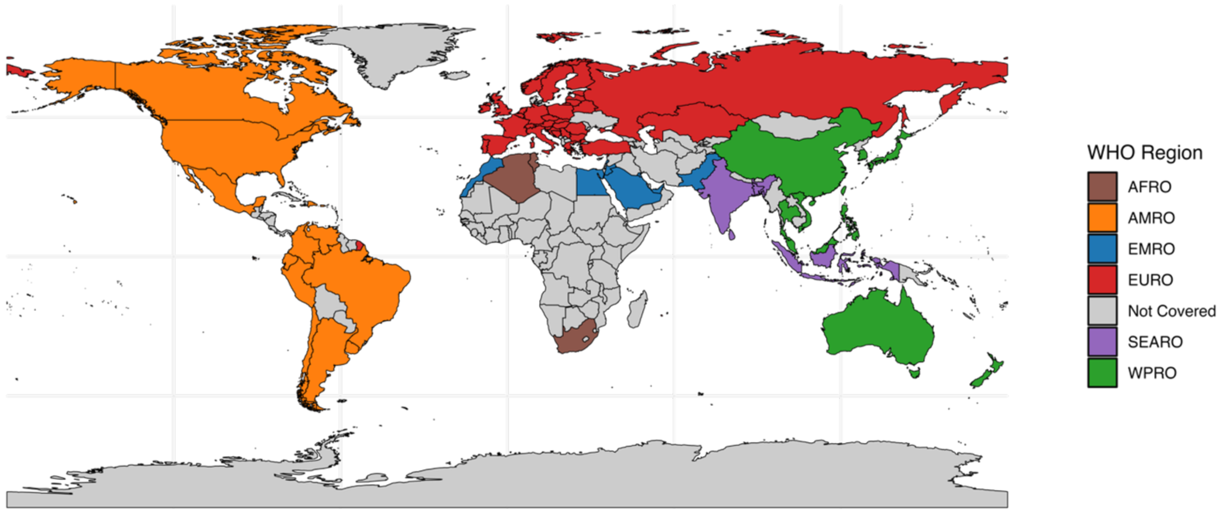

Notes: WHO= World Health Organization. AFRO= African region, AMRO= Americas region, EMRO= East Mediterranean region, EURO= Europe, SEARO= Southeast Asia region, WPRO= Western-pacific region. Countries 'Not covered' mean that they were not included in our study.

**Text A2.** Supplementary definitions

Antibiotic Use (ABU) is defined as: a metric of the quantity of a given antibiotic drug for a given location and time period, based on reported weights or international units (which may be derived from sales data, hospital data, and other sources), expressed in units of the World Health Organization's Defined Daily Dose (DDD) system. In its simplest form, ABU for a given combination of drug, location, and time period is calculated as the reported weight divided by the DDD. It is not intended that drug consumption can be inferred from this metric, although ancillary data in the original data sources may permit such inference. Instead, the metric is intended as a high-level indication of drug utilisation that can be used to inform policy decisions. The ADILA project proposed a new way to compute DDDs, and that's explained hereinafter.

### WHO DDD definitions

Defined Daily Dose (DDD) is defined on the WHO website[2] (https://www.whocc.no/ddd/definition_and_general_considera/) as follows:

> *The DDD is the assumed average maintenance dose per day for a drug used for its main indication in adults.*

For example, the WHO entry for demeclocycline on the ATC/DDD Index 2022 is:

| ATC Code | Name | DDD | U | Adm.R | Note |
|---|---|---|---|---|---|
| J01AA01 | demeclocycline | 0.6 | G | O | |

Where Adm.R is the "Route of administration" and 'O' means Oral (a full list of administration routes is included on the WHO's website here https://www.whocc.no/atc_ddd_alterations__cumulative/ddd_alterations/abbrevations/).

The WHO websites states the following:

> *The basic principle is to assign only one DDD per route of administration within an ATC code.* Prices are calculated over total packs in DDD sold.

Calculate average price per DDD
Dependent on distribution of pricing

$$Total\ DDD = \frac{Total\ Weight}{WHO\ DDD}$$

$$Total\ DDD = \frac{(Strength)(Pack\ Size)(Total\ Quantity\ of\ Packs\ Sold)}{WHO\ DDD}$$

$$Price\ per\ DDD = \frac{Average\ total\ Ex\ facotry\ Price\ 2019}{Total\ DDD}$$

*Mean/SD* *Median/IQR*

**Text A3.** List of countries included from WHO/HAI for which amoxicillin and ciprofloxacin lowest ex-manufacturer prices were available

Countries available (n=10): China, India, Russia, Indonesia, Brazil, Peru, Morocco, Lebanon, Philippines, Tunisia.

Source and more information: WHO/HAI.[3]

**Text A4.** Antibiotic price verification : WHO/HAI

Firstly, ex-manufacturer prices from WHO/HAI were, on average, 27% lower ($p25^{th}$ 10%, $p75^{th}$ 38%) for amoxicillin and 4% lower for Ciprofloxacin, compared to our volume-weighted ex-factory (ex-manufacturer) prices per DDD .Authors' analysis of IQVIA MIDAS Quarterly Sales data for the year 2019, reflecting estimates of real-world activity. Copyright IQVIA. All Rights Reserved.

**Table Text 4.1.** Minimum prices for Amoxicillin and Ciprofloxacin from WHO/HAI in US dollars not PPP, by country

| **Country** | **System** | **Access** Amoxicillin | | | **Watch** Ciprofloxacin | | |
|---|---|---|---|---|---|---|---|
| | | Dosage | Price | Currency ($) | Dosage | Price | Currency ($) |
| China | procurement public | 500mg cap | 0.075 | 2012 | 500mg cap | | |
| India | procurement public | 500mg cap | 0.035 | 2011 | 500mg cap | 0.027 | 2011 |
| Russia | procurement public | 500mg cap | 0.088 | 2011 | 500mg cap | 0.079 | 2011 |
| Indonesia | procurement public | 500mg cap | 0.041 | 2010 | 500mg cap | 0.202 | 2010 |
| Brazil | procurement public | 500mg cap | 0.043 | 2008 | 500mg cap | 0.077 | 2008 |
| Peru | procurement public | 500mg cap | 0.042 | 2005 | 500mg cap | 0.024 | 2005 |
| Morocco | procurement public | 500mg cap | 0.047 | 2004 | 500mg cap | 0.809 | 2004 |
| Lebanon | procurement public | 500mg cap | 0.044 | 2004 | 500mg cap | | |
| Philippines | procurement public | 500mg cap | 0.071 | 2002 | 500mg cap | 0.152 | 2008 |
| Tunisia | procurement public | 500mg cap | 0.047 | 2004 | 500mg cap | 0.161 | 2004 |

Notes: mg= milligrams.

**Table Text 4.2.** Minimum prices for Amoxicillin and Ciprofloxacin from WHO/HAI, by country

| Country | System | Amoxicillin<br>Difference (%) compared to IQVIA MIDAS prices | Ciprofloxacin<br>Difference (%) compared to IQVIA MIDAS prices |
|---|---|---|---|
| China | procurement public | 14.87% | |
| India | procurement public | -36.15% | -10.24% |
| Russia | procurement public | -7.13% | 5.44% |
| Indonesia | procurement public | -26.26% | 24.01% |
| Brazil | procurement public | -75.10% | -78.44% |
| Peru | procurement public | -21.80% | -77.20% |
| Morocco | procurement public | -57.81% | 83.23% |
| Lebanon | procurement public | -39.12% | |
| Philippines | procurement public | -19.20% | -37.88% |
| Tunisia | procurement public | -2.50% | 59.43% |

Notes: DDD= Daily defined dose. PPP= Purchase power parity. Procurement prices might be different to ex-manufacturer prices. Authors' analysis of IQVIA MIDAS Quarterly Sales data for the year 2019, reflecting estimates of real-world activity. Copyright IQVIA. All Rights Reserved.

### **Text A5.** Antibiotic price verification : WHO's Medicines Prices and Market Information Sources database

We compared IQVIA MIDAS® Quarterly sales data's volume-weighted prices per DDD of common prescribed antibiotics (Amoxicillin, Azithromycin, Ceftriaxone, Ciprofloxacin, Clarithromycin, Gentamicin, Meropenem, Linezolid, and Colistin) against single-entry prices in country-specific sources, documented in the WHO's Medicines Prices and Market Information Sources database.[4] Based on Table A5.1 and our systematic review of the WHO dataset, we identified Australia, Belgium, Brazil, Chile, Slovakia, South Africa, and Switzerland as countries reporting complete price data, including medicines specification, prices and their formulations.

**Table A5.1.** Selected countries reporting procurement prices from the WHO's Medicines Prices and Market Information Sources database

| Country/ Organization | Price Type | Price Link | Price Comment |
|---|---|---|---|
| Australia | Manufacturer List Price | https://www.pbs.gov.au/info/industry/pricing/ex-manufacturer-price | Approved Ex-Manufacturer Price |
| Belgium | Manufacturer List Price | https://webappsa.riziv-inami.fgov.be/SSPWebApplicationPublic/fr/Public/ProductSearch | Prix ex-usine |
| Brazil | Manufacturer List Price | https://www.gov.br/anvisa/pt-br/assuntos/medicamentos/cmed/precos | PF: maximum selling price that must be practiced by companies producing, importing or distributing medicines for pharmacies, drugstores, hospitals, clinics and for governments (when the PMVG is not applicable); |
| Brazil | Procurement/Tender Award Price | https://paineldeprecos.planejamento.gov.br/analise-materiais | # of purchase processes/value of approved purchases (Search by Nome do Material) |
| Chile | Manufacturer List Price | https://www.cenabast.cl/observatorio-de-precios-internacionales-2/ | International Reference Prices (Argentina, Brazil, Chile, Colombia, Uruguay) |
| Chile | Procurement/Tender Award Price | https://www.cenabast.cl/compras-cenabast | Quantities and unit prices |
| Slovakia | Manufacturer List Price | https://www.health.gov.sk/?zoznamy-uradne-urcenych-cien | Úradne určená cena lieku (The officially determined price of the drug: For the purposes of this law, the officially determined price of a drug is understood as the price of the drug from the manufacturer or importer, which cannot be exceeded during the first sale of the drug in the territory of the Slovak Republic or during the subsequent sale of the drug to the holder of a permit for wholesale distribution of drugs) |
| South Africa | Manufacturer List Price | https://www.health.gov.za/nhi-pee/ | ex-manufacturer price private sectors |
| South Africa | Procurement/Tender Award Price | http://www.health.gov.za/tenders/ | Master Health Product List tab |
| Switzerland | Manufacturer List Price | http://www.xn--spezialittenliste-yqb.ch/Default.aspx | Manufacturer Price (Fabrikabgabepreis, excl VAT) |
| United Kingdom | Procurement/Tender Award Price | https://www.gov.uk/government/publications/drugs-and-pharmaceutical-electronic-market-information-emit | hospital-sector; 4-month rolling average |

Notes: Data were sourced from direct country links, provided by the World Health Organization 'WHO'. https://www.who.int/teams/health-product-and-policy-standards/medicines-selection-ip-and-affordability/affordability-pricing/med-price-info-source

**Summary of the results**

Table A5.2 presents the results, followed by a country-by-country explanation below. Positive percentage values, as variation, reflect higher prices in country-specific sources compared to IQVIA MIDAS Quarterly sales data for the year 2019.

In Australia, in five of the antibiotics, the range was from +15.2% (Ciprofloxacin) to −34.8% (Azithromycin). Gentamicin, Clarithromycin, and Amoxicillin were also priced below IQVIA, with an average variation of –13.64%, reflecting generally lower local prices—likely due to the use of approved ex-manufactory pricing in national sources.

For Belgium, prices for Amoxicillin (+166.8%), Azithromycin (+132%), and Ciprofloxacin (+56.4%) were substantially higher than IQVIA. Meropenem (−27.5%) and Colistin (−48.8%) were lower. The average variance was +50.25%, with some antibiotics well above IQVIA costs.

Variability was high in Brazil for prices. Azithromycin (+85.6%) and Gentamicin (+30.9%) were higher than IQVIA, whereas Ciprofloxacin (−72.4%), Linezolid (−15.1%), and Meropenem (−9.2%) were lower. The average variation was +3.78%, which reflects a balance almost equidistant from over- and under-estimation.

In Chile, all the antibiotics were lower in price compared to IQVIA, including Amoxicillin (−38.4%), Clarithromycin (−36.2%), and Ciprofloxacin (−6.2%). Ceftriaxone (+4.2%) was the only one that had a small rise. Overall change averaged –15.27%, which means usually lower public procurement prices.

There was a mixed pattern in Slovakia. Ciprofloxacin (+25.8%) and Amoxicillin (+14.5%) were more expensive than IQVIA, but Meropenem (−34.3%) and Linezolid (−2.6%) were less expensive. The average divergence was +0.85%, showing tight overall correspondence with IQVIA figures.

In the United Kingdom, prices differed. Much lower values were for Azithromycin (–60.4%) and Ciprofloxacin (–60.5%), whereas for Gentamicin (+87.4%) and Clarithromycin (+1.9%) they were higher. Overall, variation was –9.10%, which indicated mixed pricing compared to IQVIA.

In Switzerland, country-specific pricing was low overall, with significant differences for Ceftriaxone (–55.5%), Linezolid (–43.8%), and Clarithromycin (–17.5%). Slight variations were seen only in Ciprofloxacin (+4.2%) and Clarithromycin (+1.9%). The average variation was –23.06%.

In South Africa, the greatest deviation was witnessed. Amoxicillin (+297.6%) and Azithromycin (+363.9%) were far more expensive compared to IQVIA MIDAS, while Meropenem (−67.6%) and Clarithromycin (−41.5%) were far less expensive. The overall deviation was +82.71%, indicative of a highly unstable market environment. However, these prices were drawn from private sector sources, which may account for the observed variability.

Authors' analysis of IQVIA MIDAS Quarterly Sales data for the year 2019, reflecting estimates of real-world activity. Copyright IQVIA. All Rights Reserved.

**Table A5.2.** Country-specific prices per DDD for commonly used antibiotics, based on external sources aligned with the WHO ex-manufacturer list, shown in comparison to IQVIA MIDAS Quarterly sales data for the year 2019

| Antibiotic | Australia | | | Belgium | | | Brazil | | | Chile | | |
|---|---|---|---|---|---|---|---|---|---|---|---|---|
| | Type | Price | Variation (%) | Type | Price | Variation (%) | Type | Price | Variation (%) | Type | Price | Variation (%) |
| Amoxicillin | Capsule 250 mg (as trihydrate), 20 quantity, oral | 0.28 | 12.5 | AMOXICILLINE SANDOZ 500 mg (SANDOZ) 16 | 1.59 | 166.75 | 500 MG CAP DURA CT BL AL PLAS PVC/PVDC TRANS X 20 | 2.37 | -4.29 | AMOXICILIN A 500 MG CAJ 21 CP | 0.53 | -38.43 |
| Ceftriaxone | | | | CEFTRIAXONE FRESENIUS KABI 1 g (FRESENIUS KABI) 10 injectieflacons 1 g poeder voor oplossing voor injectie | 12.31 | 1.42 | 1G PO SOL INJ IV CX 50 FA VD AMB | 23.28 | 4.20 | | | |
| Azithromycin | Tablet 500 mg (as dihydrate), oral | 0.82 | -34.8 | ZITROMAX 500 mg (Pfizer) 3 filmomhulde tabletten | 2.23 | 131.97 | 500 MG COM REV CT BL AL PLAS OPC X 2 | 2.85 | 85.59 | AZITROMICI NA 500 MG CAJ 6 CM REC | 0.35 | -0.75 |
| Ciprofloxacin | Tablet 250 mg (as hydrochloride), oral | 0.58 | 15.2 | CIPROXINE 500 (BAYER) 20 tabletten, oral | 1.82 | 56.40 | 2,0 MG/ML SOL INJ CX FR PLAS PE TRANS SIST FECH X 100 ML, parenteral | 8.98 | -72.44 | CIPROFLOXA CINO 500 MG CAJ 20 CM REC | 0.49 | -6.17 |
| Clarithromycin | Tablet 250 mg, oral | 0.27 | -31.1 | BICLAR 500 FORTE (VIATRIS) 10 omhulde tabletten | 1.26 | 74.51 | 500 MG COM REV CT BL AL PLAS OPC X 10 | 3.91 | -1.38 | CLARITROMI CINA 500 MG CAJ 14 CM REC | 0.59 | -36.21 |
| Gentamicin | Injection 80 mg (as sulfate) in 2 mL, parenteral | 2.20 | -30.0 | | | | 10 MG/ML SOL INJ CX 100 AMP VD AMB X 1 ML | 4.40 | 30.93 | | | |
| Meropenem | | | | MEROPENEM FRESENIUS KABI 500 mg (FRESENIUS KABI) 1 injectieflacon 20 ml poeder voor oplossing voor infusie en injectie | 32.83 | -27.49 | 500MG PÓ SOL INJ IV CX 10 FA VD TRANS X 30ML | 182.36 | -9.23 | | | |

| | | | | | | | | | |
|---|---|---|---|---|---|---|---|---|---|
| Linezolid | | | | | | | 2 MG/ML SOL INJ INFUS IV CX 10 ENV PLAS OPC X BOLS PLAS TRANS X 300 ML, parenteral | 155.46 | -15.06 |
| Colistin | | | | COLISTINEB 2.000.000 I.E. (TEVA PHARMA BELGIUM), 10 injectieflacons 2000000 IU, 2000000 IU - Colistimethaat, natrium- | 27.57 | -48.82 | 1.000.000 UI PO SOL INFUS IV/INAL CX 10 FA VD TRANS, parenteral | 40.27 | 13.13 |

| **Antibiotic** | **Slovakia** | | | **UK** | | | **Switzerland** | | | **South Africa** | | |
|---|---|---|---|---|---|---|---|---|---|---|---|---|
| | Type | Price | Variation (%) | Type | Price | Variation (%) | Type | Price | Variation (%) | Type | Price | Variation (%) |
| Amoxicillin | | | | Amoxicillin 250mg capsules / Packsize 21 | 0.17 | -43.82 | Amoxicillin Axapharm, Disp Tabl 500 mg, 20 Stk | 0.62 | -22.8 | Amoxicillin 875 mg TAB 10, 1 pack | 2.01 | 297.61 |
| Ceftriaxone | Ceftriaxon Kabi 1 g plv inj 10x1 g | 16.9 | 14.5 | Ceftriaxone 250mg powder for solution for injection vials / Packsize 1 | 9.92 | -60.36 | Ceftriaxon Labatec, Trockensub 1 g, Durchstf 1 Stk | 10.23 | -55.5 | 500 mg INJ pack size 2 quantity 1 | 3.90 | 25.33 |
| Azithromycin | | | | Azithromycin 250mg capsules / Packsize 6 | 0.87 | 18.91 | Zithromax, Filmtabl 250 mg, 4 Stk | 1.70 | -2.7 | Azithromycin dihydrate, 200mg/5ml, dosage form: SUSpack size:15, quantity=1. | 5.82 | 363.91 |
| Ciprofloxacin | Ciprofloxacin Kabi 200 mg/100 ml sol inf 10x100 ml/200 mg (fla) | 26.2 | 25.8 | Ciprofloxacin 250mg tablets / Packsize 10 | 0.20 | -60.51 | Cip eco, Filmtabl 250 mg, 6 Stk | 2.31 | 4.2 | Ciprofloxacin 500 mg TAB 10, pack size 1 | 0.47 | -7.51 |
| Clarithromycin | | | | Clarithromycin 250mg tablets / Packsize 14 | 0.29 | 1.90 | Clarithromycin Sandoz, Filmtabl 250 mg, 14 Stk | 1.19 | -17.5 | Clarithromycin 500 mg TAB 10 | 1.06 | -41.47 |

| | | | | | | | | | | | | |
|---|---|---|---|---|---|---|---|---|---|---|---|---|
| Gentamicin | | | | Gentamicin 20mg/2ml solution for injection ampoules / Packsize 5 | 13.16 | 87.41 | | | | Gentamicin 20 mg/2ml INJ pack size: 10 | 1.54 | 16.10 |
| Meropenem | | 32.0 | -34.3 | | | | Meronem, Trockensub 500 mg i.v, Durchstf 10 Stk | 52.65 | -27.0 | Meropenem Anhydrous 1000 mg/30ml INJ 10 | 17.53 | -67.57 |
| Linezolid | Linezolid Kabi 2 mg/ml sol inf 10x300 ml/600 mg (fl'.LDPE) | 102.4 | -2.6 | | | | Linezolid Pfizer, Inf Lös 600 mg/300ml, Freeflex 10 Stk | 70.62 | -43.8 | linezolid 600 mg TAB 10, oral | 24.43 | 61.95 |
| Colistin | | | | | | | | | | | | |

Notes: Blank cells indicate missing data. Positive percentage values reflect higher prices in country-specific sources compared to IQVIA MIDAS. WHO= World Health Organization. Source: https://www.who.int/teams/health-product-and-policy-standards/medicines-selection-ip-and-affordability/affordability-pricing/med-price-info-source

**Text A6.** Full list of antibiotic treatment course prices per antibiotic, dosage, indication and frequency.

Following the WHO-EMl-AWaRe book (https://www.who.int/publications/i/item/9789240062382), we estimated the price for antibiotic treatment courses following most reported infection diagnosis in line with indications, dosages and frequency per highlighted in Table below.
Pricing treatment courses for most common infections and most reported oral antibiotics across countries, following the AWaRe book (int$).

| Antibiotic treatment course | AWaRe category | Treatment | Indications |
|---|---|---|---|
| Amoxicillin | Access | 500mg, every 8 hours for 5 days | Pharyngitis, Acute otitis media, COPD exacerbation (mild) |
| Amoxicillin + Clavulanic acid | Access | 500mg + 125mg, every 8 hours for 5 days | Acute otitis media, Acute sinusitis |
| Cefalexin | Access | 500mg, every 8 hours for 5 days | Respiratory infections |
| Doxycycline | Access | 500mg, every 12 hours for 5 days | CAP (mild), COPD exacerbation (mild) |
| Nitrofurantoin | Access | 50mg every 6 hours for 5 days | Lower UTIs |
| Phenoxymethylpenicillin | Access | 500mg, every 6 hours for 5 days | Respiratory infections |
| Azithromycin | Watch | 500mg, single dose for 4 days | Enteric fever(mild), Infectious acute diarrhoea |
| Cefixime | Watch | 400mg single dose for 3 days | Bacterial Infections |
| Ciprofloxacin | Watch | 500mg, every 12 hours for 7 days | Upper UTI (mild), Enteric fever (mild), Febrile neutropenia (low risk) |
| Clindamycin | Watch | 600mg every 8 hours for 4 weeks | Bacterial Infections |
| Linezolid | Reserve | 600mg every 12 hours for 10 days | Mild-severe bacterial infections |

Notes: UTI= Urinary tract infections. IQR= Interquartile range. P25th= 25th percentile. Mg= milligrams. COPD= Chronic obstructive pulmonary disease. CAP= Community-acquired pneumonia.

**Text A7.** Exemplar of antibiotic treatment for sepsis among adults and paediatrics following WHO treatment.

We used the WHO's guidelines to follow an empirical treatment for adults and paediatrics for unknown cause of sepsis.[5] For the calculation of treatment pricess, we quantified human body weight (in kilograms) following 70 kilograms among adults and 3.4 kilograms among children,[6] and converted the below indications (antibiotic dosage and frequency) to WHO's daily defined doses (DDD) using the WHO's ATC/DDD index.[2] Treatments regimens are in line with WHO/AWaRe book and recent studies.[7,8]

| Treatment | Population | Antibiotic | Duration |
|---|---|---|---|
| Sepsis (i) | Adults | **Ia.** Ceftriaxone 2g every 24hrs IV **COMBINED WITH** (Amikacin 15 mg/kg every 24 hrs IV OR Gentamicin 5 mg/kg every 24 hrs IV)<br><br>**OR**<br><br>**Ib.** Cefotaxime 2 g every 8 hrs IV **COMBINED WITH** (Amikacin 15 mg/kg every 24 hrs IV OR Gentamicin 5 mg/kg every 24 hrs IV) | 7 days |
| Sepsis (ii) | Adults | **II.** Piperacillin+tazobactam 4 g+500 mg q6h IV | 7 days |
| Sepsis (iii) | Adults | **III.** Meropenem 2 g q8h IV | 7 days |
| Sepsis (iv) | Adults | **IV.** Colistin<br>Loading dose: 300 mg CBA (9 million IU CMS)<br>• Maintenance dose: 150 mg CBA<br>(4.5 million IU CMS) q12h | 7 days |
| Sepsis (i) | Paediatric | **I.** (Ampicillin 50 mg/kg/dose every 8 hrs IV **OR** Benzylpenicillin 30 mg/kg/dose (50 000 IU/kg/dose) every 8 hrs IV)<br>**COMBINED WITH** (Gentamicin 7.5 mg/kg/dose every 24 hrs IV) | 7 days |
| Sepsis (ii) | Paediatric | **II.** Cefotaxime 50 mg/kg/dose every 8 hrs IV **OR** Ceftriaxone 80 mg/kg/dose every 24 hrs IV | 7 days |
| Sepsis (iii) | Paediatric | **III.** Piperacillin+tazobactam 100 mg/kg/dose of piperacillin component q8h IV | 7 days |
| Sepsis (iv) | Paediatric | **IV.** Meropenem 40 mg/kg/dose q8h IV | 7 days |
| Sepsis (v) | Paediatric | **V.** Colistin 1.25-2.5 mg/kg/dose CBA<br>(37 500-75 000 IU/kg/dose CMS) q12h | 7 days |

Notes: We followed the AWaRe book and WHO guidelines https://app.firstline.org/en/clients/276-world-health-organization. WHO= World Health Organization. g= grams, mg= milligrams. IV= Intravenous. Hrs= hours. Kg= kilogram.

**Figure A2.** Available Access antibiotics per country in IQVIA MIDAS Quarterly sales data for the year 2019

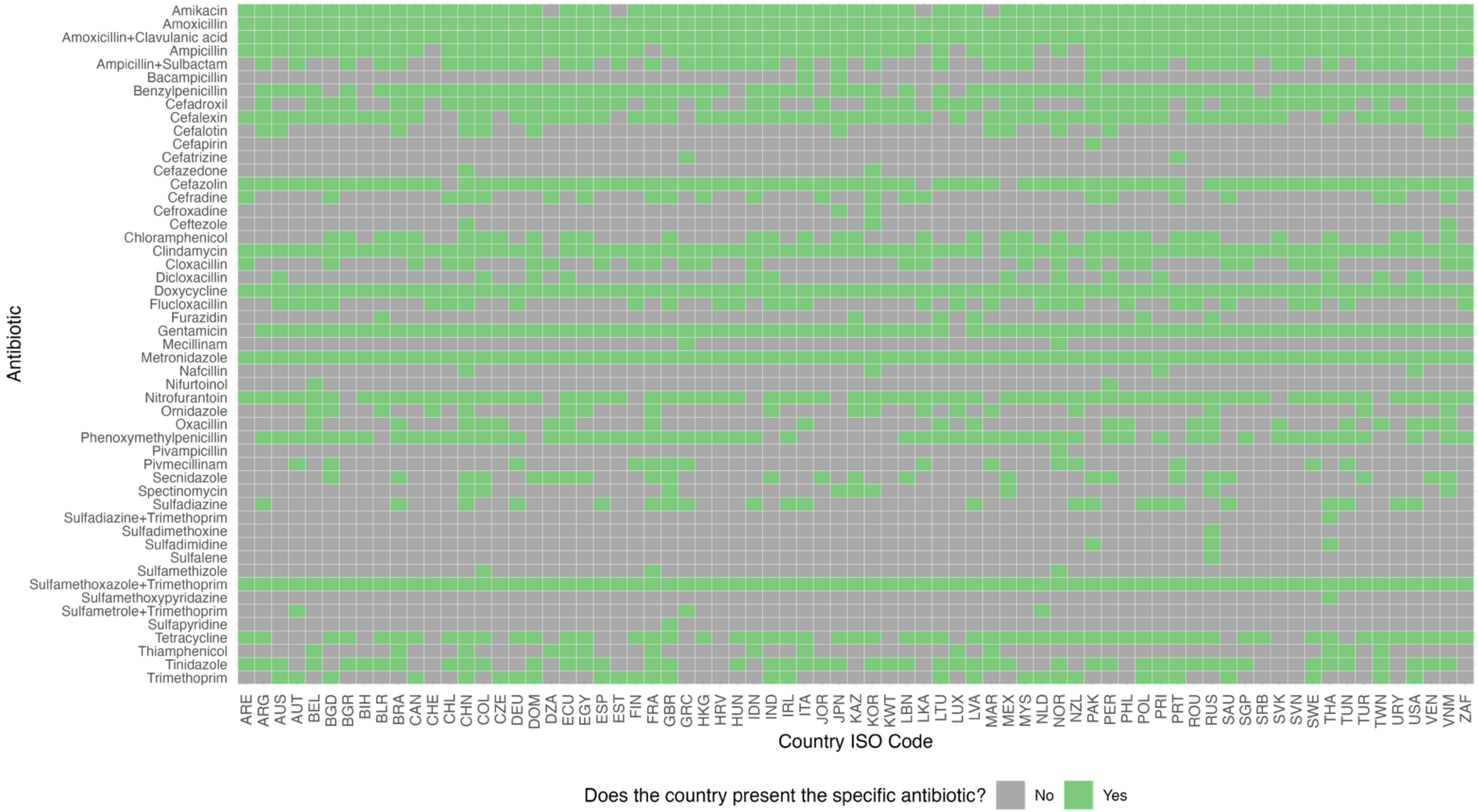

Notes: Access is based on AWaRe system classification.[8] Table A1 indicates the abbreviations of country-ISO3-codes. Authors' analysis of IQVIA MIDAS Quarterly Sales data for the year 2019, reflecting estimates of real-world activity. Copyright IQVIA. All Rights Reserved

**Figure A3.** Available Watch antibiotics per country in IQVIA MIDAS Quarterly sales data for the year 2019

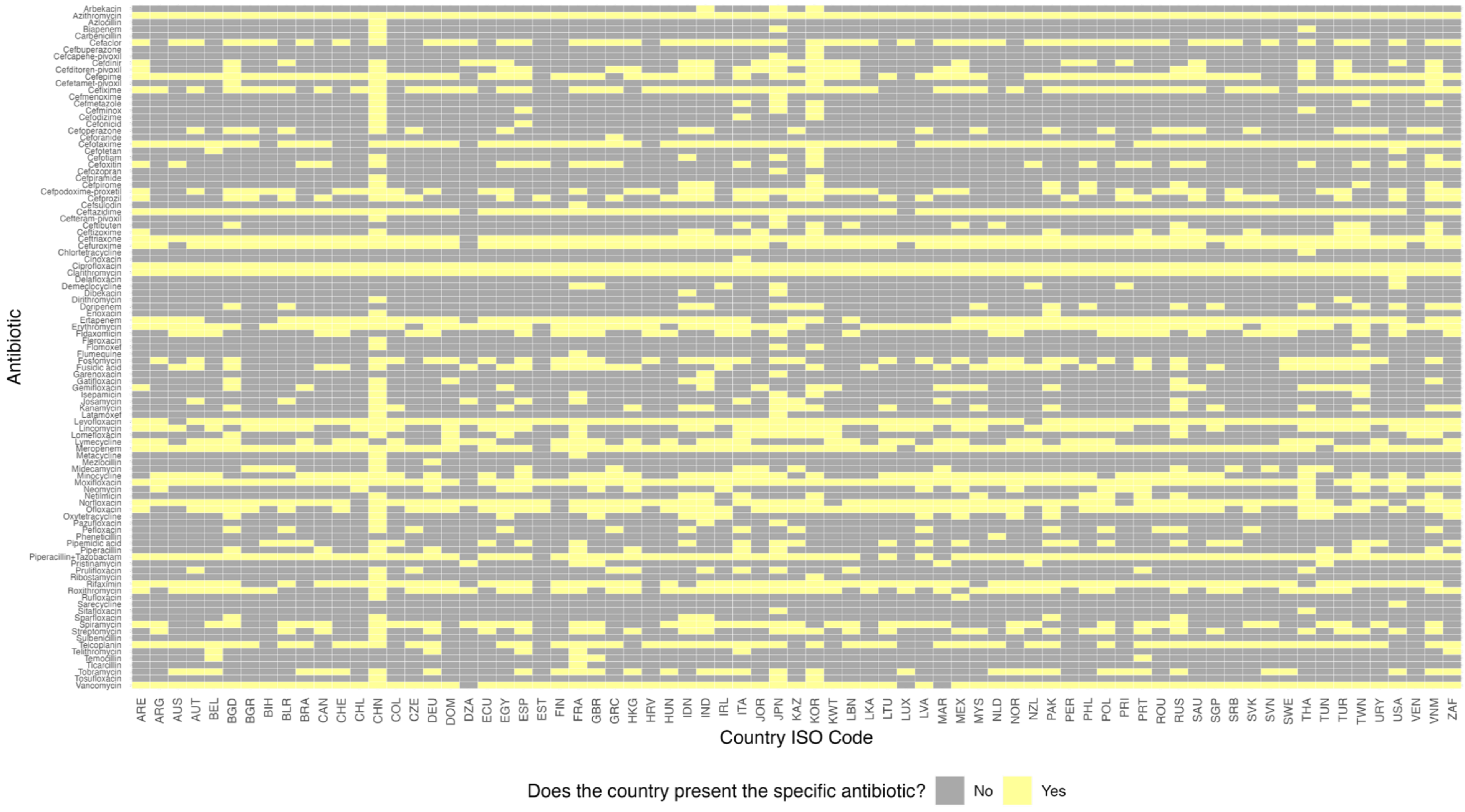

Notes: Watch is based on AWaRe system classification.[8] Table A1 indicates the abbreviations of country-ISO3-codes. Authors' analysis of IQVIA MIDAS Quarterly Sales data for the year 2019, reflecting estimates of real-world activity. Copyright IQVIA. All Rights Reserved

**Figure A4.** Available Reserve antibiotics per country in IQVIA MIDAS Quarterly sales data for the year 2019

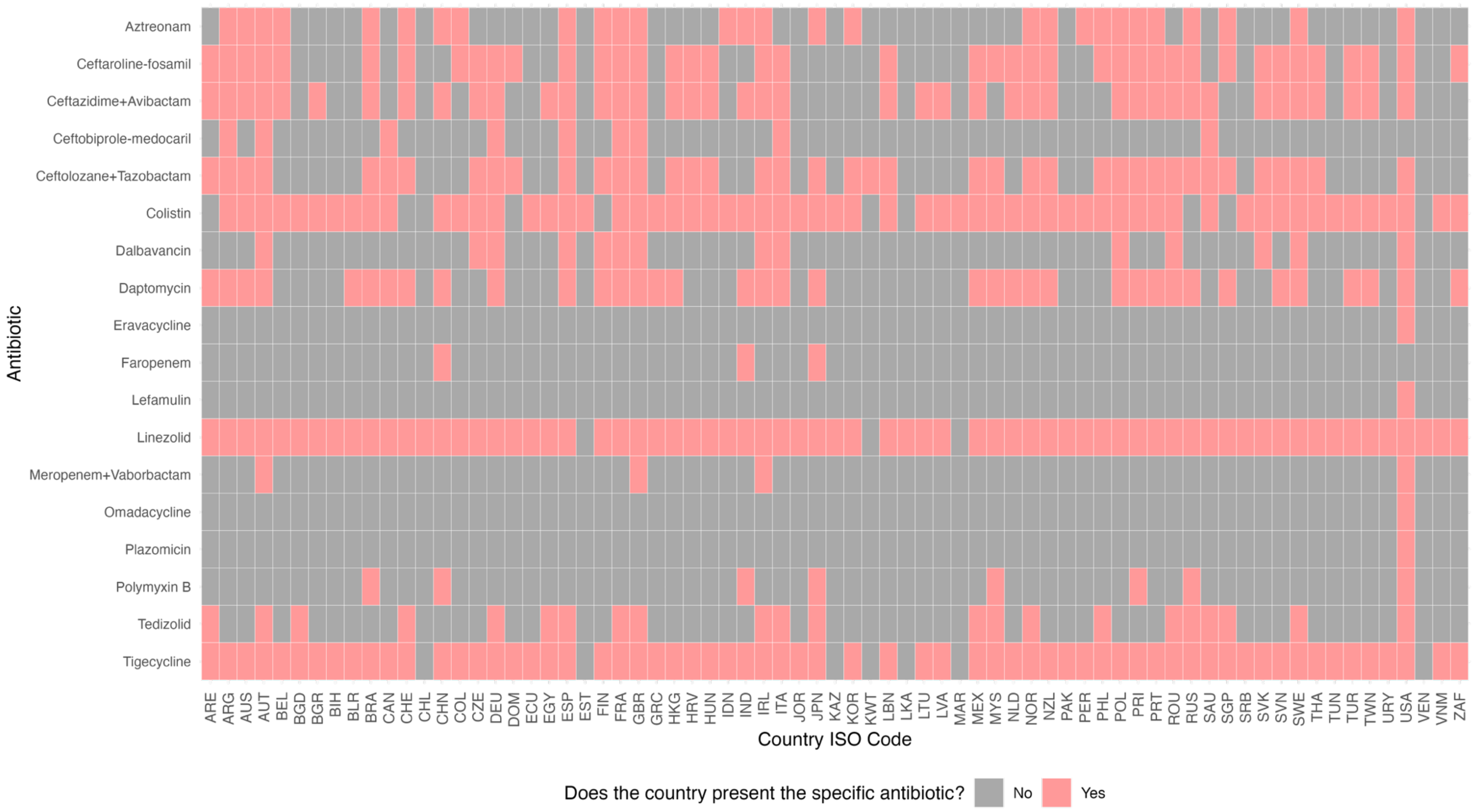

Notes: Reserve is based on AWaRe system classification.[8] Authors' analysis of IQVIA MIDAS Quarterly Sales data for the year 2019, reflecting estimates of real-world activity. Copyright IQVIA. All Rights Reserved

**Figure A5.** Available antibiotics pertaining to the EML book in IQVIA MIDAS 2019

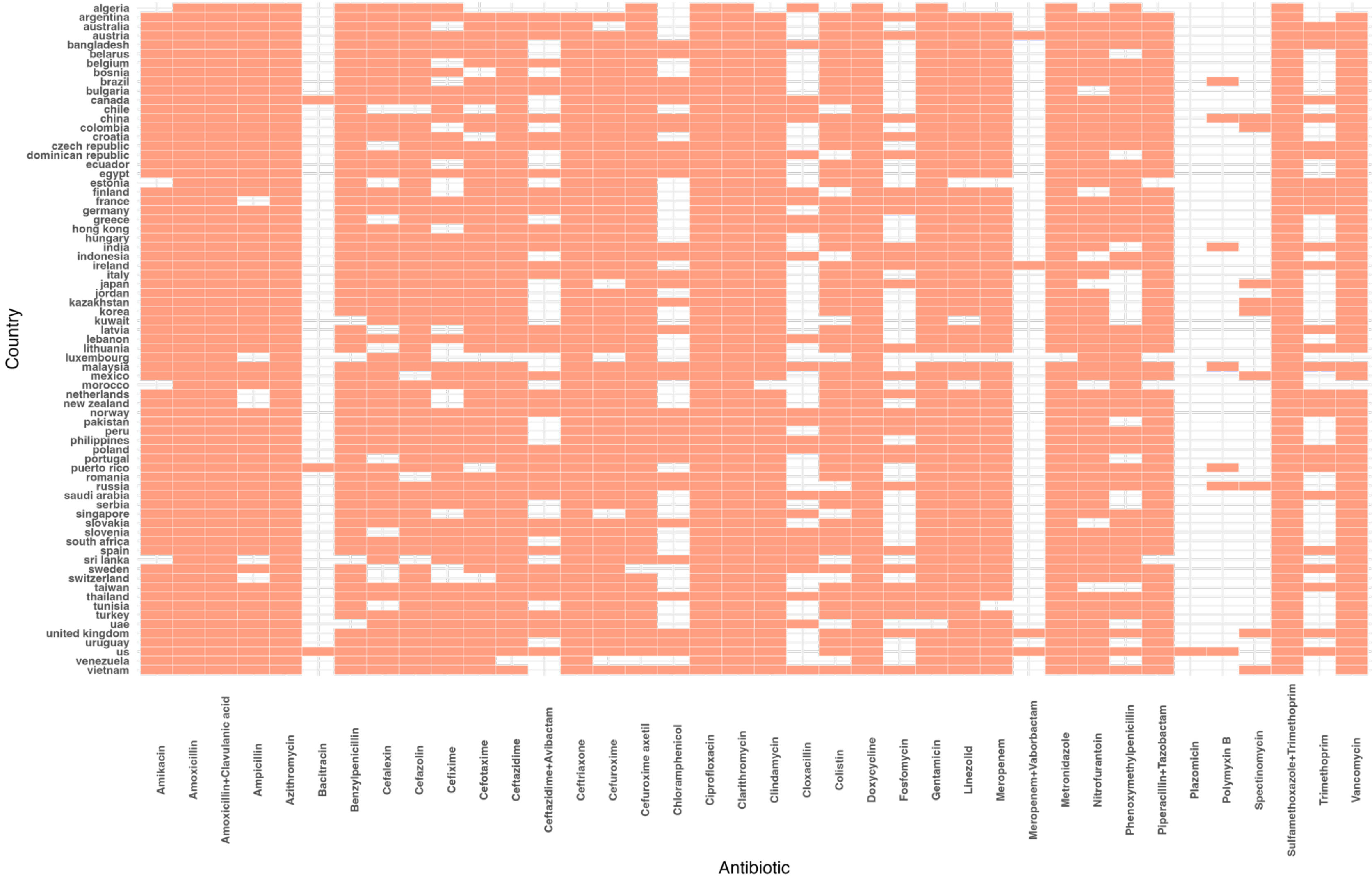

Notes: EML= Essential Medicine List. Authors' analysis of IQVIA MIDAS Quarterly Sales data for the year 2019, reflecting estimates of real-world activity. Copyright IQVIA. All Rights Reserved

**Table A3.** Antibiotics classified as Access following the EML book, data from the IQVIA MIDAS Quarterly sales data for the year of 2019

| Antibiotic | EML book | Antibiotic | EML book[9] |
|---|---|---|---|
| Amikacin | YES | Nafcillin | NO |
| Amoxicillin | YES | Nifurtoinol | NO |
| Amoxicillin+Clavulanic acid | YES | Nitrofurantoin | YES |
| Ampicillin | YES | Ornidazole | NO |
| Ampicillin+Sulbactam | NO | Oxacillin | NO |
| Bacampicillin | NO | Phenoxymethylpenicillin | YES |
| Benzylpenicillin | YES | Pivampicillin | NO |
| Cefadroxil | NO | Pivmecillinam | NO |
| Cefalexin | YES | Secnidazole | NO |
| Cefalotin | NO | Spectinomycin | YES |
| Cefapirin | NO | Sulbactam | NO |
| Cefatrizine | NO | Sulfadiazine | NO |
| Cefazedone | NO | Sulfadiazine+Trimethoprim | NO |
| Cefazolin | YES | Sulfadimethoxine | NO |
| Cefradine | NO | Sulfadimidine | NO |
| Cefroxadine | NO | Sulfalene | NO |
| Ceftezole | NO | Sulfamethizole | NO |
| Chloramphenicol | YES | Sulfamethoxazole | NO |
| Clindamycin | YES | Sulfamethoxazole+Trimethoprim | YES |
| Cloxacillin | YES | Sulfamethoxypyridazine | NO |
| Dicloxacillin | NO | Sulfametrole+Trimethoprim | NO |
| Doxycycline | YES | Sulfapyridine | NO |
| Flucloxacillin | NO | Tetracycline | NO |
| Furazidin | NO | Thiamphenicol | NO |
| Gentamicin | YES | Tinidazole | NO |
| Mecillinam | NO | Trimethoprim | YES |
| Metronidazole | YES | | |

Notes: EML= Essential Medicine List. Authors' analysis of IQVIA MIDAS Quarterly Sales data for the year 2019, reflecting estimates of real-world activity. Copyright IQVIA. All Rights Reserved.

**Table A4.** Antibiotics classified as Watch following the EML book, data from the IQVIA MIDAS Quarterly sales data for the year 2019

| Antibiotic | EML book | Antibiotic | EML book | Antibiotic | EML book[9] |
|---|---|---|---|---|---|
| Arbekacin | NO | Cinoxacin | NO | Pazufloxacin | NO |
| Azithromycin | YES | Ciprofloxacin | YES | Pefloxacin | NO |
| Azlocillin | NO | Clarithromycin | YES | Pheneticillin | NO |
| Biapenem | NO | Delafloxacin | NO | Pipemidic acid | NO |
| Carbenicillin | NO | Demeclocycline | NO | Piperacillin | NO |
| Cefaclor | NO | Dibekacin | NO | Piperacillin+Tazobactam | YES |
| Cefamandole | NO | Dirithromycin | NO | Pristinamycin | NO |
| Cefbuperazone | NO | Doripenem | NO | Prulifloxacin | NO |
| Cefcapene-pivoxil | NO | Enoxacin | NO | Ribostamycin | NO |
| Cefdinir | NO | Ertapenem | NO | Rifamycin | NO |
| Cefditoren-pivoxil | NO | Erythromycin | NO | Rifaximin | NO |
| Cefepime | NO | Fidaxomicin | NO | Roxithromycin | NO |
| Cefetamet-pivoxil | NO | Fleroxacin | NO | Rufloxacin | NO |
| Cefixime | YES | Flomoxef | NO | Sarecycline | NO |
| Cefmenoxime | NO | Flumequine | NO | Sitafloxacin | NO |
| Cefmetazole | NO | Fosfomycin | NO | Sparfloxacin | NO |
| Cefminox | NO | Fusidic acid | NO | Spiramycin | NO |
| Cefodizime | NO | Garenoxacin | NO | Streptomycin | NO |
| Cefonicid | NO | Gatifloxacin | NO | Sulbenicillin | NO |
| Cefoperazone | NO | Gemifloxacin | NO | Tebipenem | NO |
| Ceforanide | NO | Isepamicin | NO | Teicoplanin | NO |
| Cefotaxime | YES | Josamycin | NO | Telithromycin | NO |
| Cefotetan | NO | Kanamycin | NO | Temocillin | NO |
| Cefotiam | NO | Latamoxef | NO | Ticarcillin | NO |
| Cefoxitin | NO | Levofloxacin | NO | Tobramycin | NO |
| Cefozopran | NO | Lincomycin | NO | Tosufloxacin | NO |
| Cefpiramide | NO | Lomefloxacin | NO | Vancomycin | YES |
| Cefpirome | NO | Lymecycline | NO | | |
| Cefpodoxime-proxetil | NO | Meropenem | YES | | |
| Cefprozil | NO | Metacycline | NO | | |
| Cefsulodin | NO | Mezlocillin | NO | | |
| Ceftazidime | YES | Midecamycin | NO | | |
| Cefteram-pivoxil | NO | Minocycline | NO | | |
| Ceftibuten | NO | Moxifloxacin | NO | | |
| Ceftizoxime | NO | Neomycin | NO | | |
| Ceftriaxone | YES | Netilmicin | NO | | |
| Cefuroxime | YES | Norfloxacin | NO | | |
| Cefuroxime axetil | YES | Ofloxacin | NO | | |
| Chlortetracycline | NO | Oxytetracycline | NO | | |

Notes: EML= Essential Medicine List. Authors' analysis of IQVIA MIDAS Quarterly Sales data for the year 2019, reflecting estimates of real-world activity. Copyright IQVIA. All Rights Reserved.

**Table A5.** Antibiotics classified as Reserve following the EML book, data from the IQVIA MIDAS Quarterly sales data for the year 2019

| Antibiotic | EML book[9] |
|---|---|
| Aztreonam | NO |
| Ceftaroline-fosamil | NO |
| Ceftazidime+Avibactam | YES |
| Ceftobiprole-medocaril | NO |
| Ceftolozane+Tazobactam | NO |
| Colistin | YES |
| Dalbavancin | NO |
| Daptomycin | NO |
| Eravacycline | NO |
| Faropenem | NO |
| Fosfomycin | YES |
| Lefamulin | NO |
| Linezolid | YES |
| Meropenem+Vaborbactam | YES |
| Minocycline | NO |
| Omadacycline | NO |
| Oritavancin | NO |
| Plazomicin | YES |
| Polymyxin B | YES |
| Tedizolid | NO |
| Telavancin | NO |
| Tigecycline | NO |

Notes: EML= Essential Medicine List. Authors' analysis of IQVIA MIDAS Quarterly Sales data for the year 2019, reflecting estimates of real-world activity. Copyright IQVIA. All Rights Reserved.

**Table A6.** Antibiotics classified as unclassified following the EML book, data from the IQVIA MIDAS Quarterly sales data for the year 2019

| Antibiotic | Antibiotic | Antibiotic |
|---|---|---|
| Albendazole+Secnidazole | Ampicillin+Chlorphenamine+Lidocaine+Guaiacol+ Metamizole sodium | Cefotaxime+Lidocaine |
| Ambroxol+Amoxicillin | Ampicillin+Guaifenesin+ Metamizole sodium | Cefpirome+Sulbactam |
| Ambroxol+Amoxicillin+Clavulanic acid | Ampicillin+Lidocaine+Niaouli Oil | Cefpodoxime-proxetil+Lactobacillus acidophilus |
| Ambroxol+Ampicillin+Guaifenesin+Lidocaine+Cineole+Niaouli Oil | Ampicillin+Lidocaine+Terpin hydrate | Ceftriaxone+Lidocaine |
| Ambroxol+Azithromycin | Ampicillin+Phenazopyridine | Cefuroxime axetil+Clavulanic acid |
| Ambroxol+Cefaclor | Ampicillin+Probenecid | Cellulose+Doxycycline |
| Ambroxol+Cefadroxil | Arginine+Cefepime | Chloramphenicol+Chlorphenamine+Guaifenesin+Phenylephrine+Iodine+Oxeladin+Prednisolone |
| Ambroxol+Cefalexin | Azithromycin+Bacillus coagulans+Saccharomyces boulardii | Chloramphenicol+Citric acid+Paracetamol+Phenylephrine+Menthol |
| Ambroxol+Clarithromycin | Azithromycin+Lactobacillus acidophilus | Chloramphenicol+Methylthioninium |
| Ambroxol+Doxycycline | Azithromycin+Nimesulide | Chlorphenamine+Guaifenesin+Oxytetracycline+Metamizole sodium |
| Ambroxol+Gemifloxacin | Azithromycin+Zinc | Chlorphenamine+Lidocaine+Tetracycline+Ascorbic acid+Metamizole sodium |
| Ambroxol+Levofloxacin | Bacitracin | Ciprofloxacin+Flavoxate |
| Ambroxol+Roxithromycin | Benzylpenicillin+Ascorbic acid+Metamizole sodium | Ciprofloxacin+Phenazopyridine |
| Amoxicillin+Acetylcysteine | Benzylpenicillin+Lidocaine | Citric acid+Metronidazole+Sodium+Phosphoric acid |
| Amoxicillin+Bacillus coagulans | Benzylpenicillin+Procaine | Clarithromycin+Tinidazole+Lansoprazole |
| Amoxicillin+Bromelain | Benzylpenicillin+Tolycaine | Clarithromycin+Tinidazole+Omeprazole |
| Amoxicillin+Bromhexine | Bezoar bovis+Metronidazole | Colistin |
| Amoxicillin+Bromhexine+Chlorphenamine | Bismuth+Kanamycin+Pectin+Attapulgite+Dimevamide | Colistin+Kaolin+Pectin |
| Amoxicillin+Bromhexine+Oxolamine | Bismuth+Metronidazole+Tetracycline | Collagen+Gentamicin |
| Amoxicillin+Bromhexine+Phenylephrine+Dextromethorphan | Bismuth+Neomycin+Procaine+Menthol | Cyanocobalamin+Folic acid+Nicotinamide+Oxytetracycline+Pantothenic acid+Pyridoxine+Riboflavin+Thiamine |
| Amoxicillin+Brovanexine | Bromhexine+Cefaclor | Dextran+Linezolid |
| Amoxicillin+Carbocisteine | Bromhexine+Cefalexin | Doxycycline+Beta.Cyclodextrin |
| Amoxicillin+Clarithromycin+Esomeprazole | Bromhexine+Chlorphenamine+Erythromycin | Doxycycline+Beta.Cyclodextrin+Lactobacillus acidophilus |
| Amoxicillin+Clarithromycin+Omeprazole | Bromhexine+Erythromycin | Doxycycline+Lactobacillus acidophilus |
| Amoxicillin+Clavulanic acid+Lactobacillus acidophilus | Bromhexine+Tetracycline | Doxycycline+Lactobacillus lactis |

| | | |
|---|---|---|
| Amoxicillin+Diclofenac | Cefadroxil+Lactobacillus acidophilus | Doxycycline+Saccharomyces boulardii |
| Amoxicillin+Dicloxacillin+Lactobacillus acidophilus | Cefalexin+Carbocisteine | Doxycycline+Serrapeptase |
| Amoxicillin+Lactobacillus acidophilus | Cefalexin+Probenecid | Ethylmorphine+Guaifenesin+Tetracycline |
| Amoxicillin+Sulbactam | Cefazolin+Lidocaine | Flavoxate+Ofloxacin |
| Ampicillin+Bromhexine | Cefdinir+Lactobacillus acidophilus | Fluconazole+Ornidazole |
| Ampicillin+Bromhexine+Chlorphenamine+Lidocaine+Cineole+Clonixin+Niaouli Oil | Cefixime+Acetylcysteine | Fluconazole+Secnidazole |
| Ampicillin+Bromhexine+Guaifenesin+Eucalyptus globulus+Niaouli Oil | Cefixime+Lactobacillus acidophilus | Fluconazole+Tinidazole |
| Ampicillin+Bromhexine+Guaifenesin+Lidocaine+Cineole+Niaouli Oil | Cefixime+Lactobacillus lactis | Fosfomycin+Phenazopyridine |

Notes: These antibiotics are not yet classified by the EML book; EML Essential Medicine List. Authors' analysis of IQVIA MIDAS Quarterly Sales data for the year 2019, reflecting estimates of real-world activity. Copyright IQVIA. All Rights Reserved.

**Table A7.** Antibiotics classified as 'Not recommended' following the EML book, data from the IQVIA MIDAS Quarterly sales data for the year 2019

| Antibiotics |
| --- |
| Acetylspiramycin+Metronidazole |
| Amikacin+Cefepime |
| Amoxicillin+Cloxacillin |
| Amoxicillin+Dicloxacillin |
| Amoxicillin+Flucloxacillin |
| Amoxicillin+Metronidazole |
| Amoxicillin+Pivsulbactam |
| Ampicillin+Cloxacillin |
| Ampicillin+Dicloxacillin |
| Ampicillin+Flucloxacillin |
| Azithromycin+Cefixime |
| Azithromycin+Cefpodoxime-proxetil |
| Bromhexine+Sulfamethoxazole+Trimethoprim |
| Cefepime+Tazobactam |
| Cefixime+Dicloxacillin |
| Cefixime+Ofloxacin |
| Cefoperazone+Sulbactam |
| Cefoperazone+Tazobactam |
| Cefpodoxime-proxetil+Clavulanic acid |
| Cefpodoxime-proxetil+Sulbactam |
| Ceftazidime+Sulbactam |
| Ceftazidime+Tazobactam |
| Ceftriaxone+Sulbactam |
| Ceftriaxone+Tazobactam |
| Ciprofloxacin+Ornidazole |
| Ciprofloxacin+Tinidazole |
| Metronidazole+Norfloxacin |
| Metronidazole+Spiramycin |
| Ofloxacin+Ornidazole |
| Piperacillin+Sulbactam |

Notes: These antibiotics are not recommended by the EML book; EML Essential Medicine List. Authors' analysis of IQVIA MIDAS Quarterly Sales data for the year 2019, reflecting estimates of real-world activity. Copyright IQVIA. All Rights Reserved.

**Table A8.** Volume-weighted ex-manufacturer prices (Int$) per country by AWaRe category, data from the IQVIA MIDAS Quarterly sales data for the year 2019

| Country ISO-3 code | Access | | | | Reserve | | | | Watch | | | |
|---|---|---|---|---|---|---|---|---|---|---|---|---|
| | Median | p25 | p75 | IQR | Median | p25 | p75 | IQR | Median | p25 | p75 | IQR |
| ARE | 2.70 | 1.48 | 8.83 | 7.35 | 319.36 | 212.04 | 542.64 | 330.60 | 4.57 | 3.23 | 56.41 | 53.18 |
| ARG | 1.71 | 1.54 | 17.62 | 16.09 | 65.12 | 88.46 | 345.28 | 256.83 | 2.29 | 2.30 | 48.34 | 46.04 |
| AUS | 0.49 | 0.45 | 3.78 | 3.33 | 97.21 | 120.12 | 313.01 | 192.89 | 1.48 | 0.81 | 13.14 | 12.32 |
| AUT | 2.73 | 0.89 | 8.92 | 8.03 | 119.24 | 116.67 | 439.44 | 322.76 | 9.19 | 2.23 | 47.97 | 45.74 |
| BEL | 1.01 | 1.06 | 7.37 | 6.31 | 62.45 | 52.12 | 145.09 | 92.97 | 2.48 | 0.96 | 31.79 | 30.83 |
| BGD | 0.68 | 0.61 | 1.98 | 1.37 | 5.44 | 15.22 | 64.26 | 49.04 | 1.42 | 1.01 | 12.86 | 11.86 |
| BGR | 2.19 | 1.98 | 5.50 | 3.52 | 87.15 | 84.63 | 396.82 | 312.19 | 3.11 | 2.28 | 15.20 | 12.93 |
| BIH | 1.54 | 1.23 | 5.33 | 4.10 | 180.64 | 166.30 | 218.42 | 52.12 | 2.20 | 1.87 | 15.70 | 13.83 |
| BLR | 1.53 | 0.72 | 2.62 | 1.90 | 106.01 | 133.86 | 184.87 | 51.01 | 2.76 | 1.71 | 7.22 | 5.51 |
| BRA | 2.10 | 1.51 | 5.26 | 3.75 | 156.82 | 123.87 | 516.08 | 392.21 | 2.27 | 3.06 | 54.09 | 51.04 |
| CAN | 1.09 | 0.96 | 9.19 | 8.23 | 78.20 | 49.65 | 125.94 | 76.29 | 2.45 | 1.44 | 31.92 | 30.48 |
| CHE | 1.91 | 1.02 | 12.17 | 11.16 | 102.87 | 138.78 | 316.11 | 177.33 | 5.13 | 2.37 | 48.05 | 45.68 |
| CHL | 1.26 | 0.81 | 2.72 | 1.91 | 194.43 | 194.43 | 194.43 | 0.00 | 1.26 | 2.05 | 93.12 | 91.07 |
| CHN | 1.87 | 0.37 | 8.59 | 8.22 | 86.27 | 23.18 | 791.02 | 767.84 | 4.97 | 1.73 | 24.15 | 22.41 |
| COL | 1.14 | 1.20 | 6.90 | 5.71 | 50.75 | 62.58 | 170.44 | 107.87 | 1.45 | 3.71 | 15.37 | 11.67 |
| CZE | 1.37 | 0.94 | 6.52 | 5.59 | 115.64 | 128.62 | 1073.18 | 944.56 | 2.45 | 2.15 | 23.43 | 21.28 |
| DEU | 1.34 | 0.59 | 3.29 | 2.69 | 48.54 | 51.56 | 438.89 | 387.33 | 2.24 | 1.07 | 16.13 | 15.06 |
| DOM | 4.84 | 1.75 | 10.39 | 8.64 | 200.91 | 146.63 | 624.45 | 477.82 | 4.78 | 4.35 | 54.05 | 49.71 |
| DZA | 1.55 | 1.99 | 9.33 | 7.33 | | | | | 3.27 | 1.74 | 5.98 | 4.25 |
| ECU | 1.57 | 1.01 | 4.42 | 3.41 | 125.87 | 118.69 | 153.56 | 34.88 | 0.45 | 1.99 | 15.14 | 13.15 |
| EGY | 1.90 | 0.65 | 3.77 | 3.12 | 6.04 | 4.87 | 103.00 | 98.13 | 3.48 | 1.21 | 15.18 | 13.97 |
| ESP | 0.61 | 0.53 | 3.95 | 3.43 | 112.66 | 90.38 | 519.49 | 429.11 | 2.72 | 1.28 | 12.62 | 11.34 |
| EST | 1.07 | 1.13 | 4.59 | 3.47 | 48.49 | 48.49 | 48.49 | 0.00 | 1.18 | 1.56 | 29.61 | 28.05 |
| FIN | 1.24 | 0.84 | 10.79 | 9.94 | 77.34 | 83.43 | 501.07 | 417.64 | 4.04 | 2.39 | 30.60 | 28.21 |
| FRA | 0.91 | 1.22 | 8.58 | 7.36 | 119.40 | 99.35 | 429.91 | 330.55 | 5.72 | 2.10 | 48.89 | 46.79 |
| GBR | 1.07 | 1.01 | 8.47 | 7.46 | 60.82 | 79.44 | 393.30 | 313.86 | 3.09 | 2.15 | 27.06 | 24.91 |
| GRC | 0.79 | 0.92 | 11.52 | 10.59 | 100.80 | 92.82 | 110.32 | 17.50 | 1.19 | 1.29 | 13.14 | 11.84 |
| HKG | 0.59 | 0.30 | 2.25 | 1.95 | 162.61 | 129.80 | 325.79 | 195.98 | 2.09 | 0.78 | 15.75 | 14.97 |
| HRV | 11.49 | 11.75 | 114.85 | 103.10 | 1479.44 | 1202.90 | 5274.65 | 4071.75 | 30.36 | 15.96 | 332.79 | 316.84 |
| HUN | 1.69 | 1.16 | 10.12 | 8.96 | 141.42 | 188.92 | 695.72 | 506.79 | 2.70 | 1.98 | 36.71 | 34.73 |
| IDN | 1.57 | 0.62 | 6.92 | 6.30 | 133.52 | 142.03 | 240.91 | 98.89 | 5.11 | 3.05 | 58.65 | 55.59 |
| IND | 1.25 | 0.63 | 4.64 | 4.01 | 7.50 | 19.62 | 165.78 | 146.16 | 1.11 | 0.88 | 23.45 | 22.57 |
| IRL | 1.04 | 0.70 | 6.33 | 5.63 | 70.07 | 70.90 | 425.41 | 354.51 | 3.72 | 1.81 | 23.39 | 21.58 |
| ITA | 1.30 | 1.29 | 9.56 | 8.27 | 150.17 | 143.21 | 503.76 | 360.54 | 3.64 | 1.99 | 17.30 | 15.31 |
| JOR | 1.84 | 0.93 | 6.44 | 5.51 | 229.08 | 192.23 | 223.12 | 30.89 | 3.82 | 2.71 | 13.35 | 10.64 |
| JPN | 2.98 | 1.58 | 11.57 | 10.00 | 16.42 | 42.49 | 175.86 | 133.36 | 2.60 | 1.84 | 21.31 | 19.47 |
| KAZ | 1.63 | 0.97 | 3.87 | 2.90 | 7.04 | 84.67 | 240.13 | 155.47 | 2.98 | 1.76 | 11.10 | 9.34 |
| KOR | 1.33 | 0.88 | 5.53 | 4.65 | 72.73 | 68.85 | 113.06 | 44.21 | 3.49 | 1.68 | 24.68 | 23.00 |
| KWT | 2.44 | 1.36 | 16.20 | 14.84 | 1257.85 | 1257.85 | 1257.85 | 0.00 | 4.38 | 2.53 | 32.22 | 29.69 |
| LBN | 1.21 | 0.61 | 2.98 | 2.37 | 46.92 | 205.06 | 691.12 | 486.06 | 3.41 | 3.04 | 21.93 | 18.89 |
| LKA | 1.15 | 0.45 | 2.46 | 2.01 | 21.59 | 21.59 | 21.59 | 0.00 | 1.00 | 0.74 | 22.24 | 21.51 |
| LTU | 1.21 | 1.07 | 3.53 | 2.47 | 60.29 | 43.40 | 226.78 | 183.38 | 2.13 | 1.80 | 9.03 | 7.23 |

| | | | | | | | | | | | | |
|---|---|---|---|---|---|---|---|---|---|---|---|---|
| LUX | 0.61 | 0.57 | 2.22 | 1.65 | | | | | 0.86 | 0.64 | 1.78 | 1.14 |
| LVA | 1.17 | 0.85 | 4.30 | 3.45 | 190.87 | 161.27 | 372.86 | 211.59 | 2.45 | 1.83 | 10.01 | 8.18 |
| MAR | 1.64 | 1.52 | 7.32 | 5.80 | 61.32 | 61.32 | 61.32 | 0.00 | 2.93 | 2.19 | 10.73 | 8.54 |
| MEX | 3.10 | 2.11 | 12.24 | 10.12 | 144.94 | 169.91 | 402.61 | 232.71 | 5.36 | 3.80 | 24.49 | 20.69 |
| MYS | 1.56 | 0.69 | 6.55 | 5.86 | 169.38 | 159.48 | 235.45 | 75.97 | 4.05 | 1.90 | 23.10 | 21.20 |
| NLD | 1.02 | 0.72 | 11.49 | 10.77 | 62.19 | 90.03 | 145.01 | 54.98 | 2.97 | 1.39 | 41.08 | 39.69 |
| NOR | 1.03 | 0.89 | 10.14 | 9.25 | 56.71 | 64.15 | 150.15 | 85.99 | 2.89 | 1.05 | 18.45 | 17.39 |
| NZL | 0.38 | 0.33 | 4.40 | 4.06 | 76.06 | 83.64 | 163.87 | 80.23 | 0.87 | 0.73 | 18.60 | 17.86 |
| PAK | 0.78 | 0.43 | 2.18 | 1.75 | 5.35 | 74.05 | 182.03 | 107.99 | 1.87 | 1.14 | 24.58 | 23.44 |
| PER | 1.22 | 1.11 | 3.68 | 2.57 | 117.82 | 112.73 | 197.86 | 85.13 | 1.21 | 1.74 | 8.45 | 6.71 |
| PHL | 3.97 | 2.60 | 17.09 | 14.49 | 390.19 | 347.23 | 418.31 | 71.08 | 6.86 | 3.46 | 133.94 | 130.47 |
| POL | 1.34 | 1.20 | 7.48 | 6.27 | 88.04 | 220.29 | 1050.39 | 830.10 | 2.10 | 2.11 | 32.11 | 29.99 |
| PRI | 2.72 | 2.66 | 12.30 | 9.64 | 207.91 | 101.84 | 462.29 | 360.45 | 6.47 | 4.93 | 40.60 | 35.67 |
| PRT | 1.08 | 1.75 | 5.97 | 4.22 | 92.48 | 74.54 | 182.54 | 108.00 | 1.95 | 2.01 | 14.39 | 12.38 |
| ROU | 1.26 | 0.78 | 4.31 | 3.53 | 118.07 | 129.56 | 985.82 | 856.26 | 2.69 | 1.85 | 13.83 | 11.98 |
| RUS | 1.29 | 0.48 | 2.50 | 2.01 | 93.83 | 131.39 | 264.79 | 133.40 | 1.97 | 1.53 | 14.91 | 13.39 |
| SAU | 3.15 | 2.94 | 12.63 | 9.70 | 212.40 | 196.18 | 592.33 | 396.15 | 5.98 | 3.52 | 58.37 | 54.85 |
| SGP | 0.96 | 0.23 | 2.45 | 2.22 | 107.47 | 97.32 | 184.48 | 87.16 | 2.61 | 1.02 | 12.23 | 11.22 |
| SRB | 1.32 | 1.32 | 5.26 | 3.94 | 167.17 | 133.81 | 199.49 | 65.68 | 2.02 | 1.66 | 12.60 | 10.94 |
| SVK | 1.43 | 1.32 | 8.46 | 7.14 | 74.22 | 112.78 | 1052.31 | 939.52 | 1.68 | 2.07 | 37.76 | 35.69 |
| SVN | 2.37 | 1.86 | 12.36 | 10.50 | 169.46 | 116.19 | 480.35 | 364.16 | 4.81 | 3.21 | 45.80 | 42.59 |
| SWE | 0.96 | 0.72 | 5.44 | 4.72 | 71.29 | 83.11 | 330.37 | 247.26 | 4.68 | 1.76 | 49.82 | 48.05 |
| THA | 0.97 | 0.37 | 3.95 | 3.58 | 91.11 | 137.84 | 629.82 | 491.98 | 4.95 | 2.24 | 43.59 | 41.35 |
| TUN | 1.23 | 0.80 | 4.04 | 3.25 | 225.01 | 222.61 | 378.48 | 155.86 | 2.83 | 1.80 | 9.71 | 7.91 |
| TUR | 0.98 | 0.79 | 6.39 | 5.60 | 25.23 | 23.22 | 133.19 | 109.97 | 1.61 | 1.36 | 15.66 | 14.31 |
| TWN | 1.28 | 0.57 | 6.44 | 5.86 | 133.87 | 100.98 | 206.81 | 105.84 | 12.77 | 1.28 | 45.10 | 43.82 |
| URY | 1.23 | 0.99 | 3.16 | 2.17 | 43.48 | 44.04 | 67.50 | 23.46 | 1.49 | 1.58 | 22.82 | 21.24 |
| USA | 0.95 | 0.92 | 8.27 | 7.34 | 94.02 | 112.68 | 811.47 | 698.79 | 5.19 | 3.38 | 37.94 | 34.56 |
| VEN | 0.90 | 0.75 | 3.56 | 2.81 | 21.86 | 21.86 | 21.86 | 0.00 | 0.68 | 0.53 | 4.23 | 3.70 |
| VNM | 1.27 | 0.43 | 8.59 | 8.17 | 96.21 | 35.70 | 194.50 | 158.79 | 3.04 | 1.42 | 23.97 | 22.55 |
| ZAF | 0.81 | 0.65 | 3.96 | 3.30 | 45.87 | 29.00 | 184.04 | 155.04 | 2.94 | 1.92 | 23.73 | 21.81 |

Notes: IQR= Interquartile range (percentile $75^{th}$ – percentile $25^{th}$). p25= percentile $25^{th}$, p75= percentile $75^{th}$. Int$= International dollars, 2019. Table A1 indicates the abbreviations of country-ISO3-codes. Authors' analysis of IQVIA MIDAS Quarterly Sales data for the year 2019, reflecting estimates of real-world activity. Copyright IQVIA. All Rights Reserved.

**Figure A6.** Average volume-weighted ex-manufacturer price per DDD by AWaRe category and across WHO regions and income groups, data from the IQVIA MIDAS Quarterly sales data for the year 2019

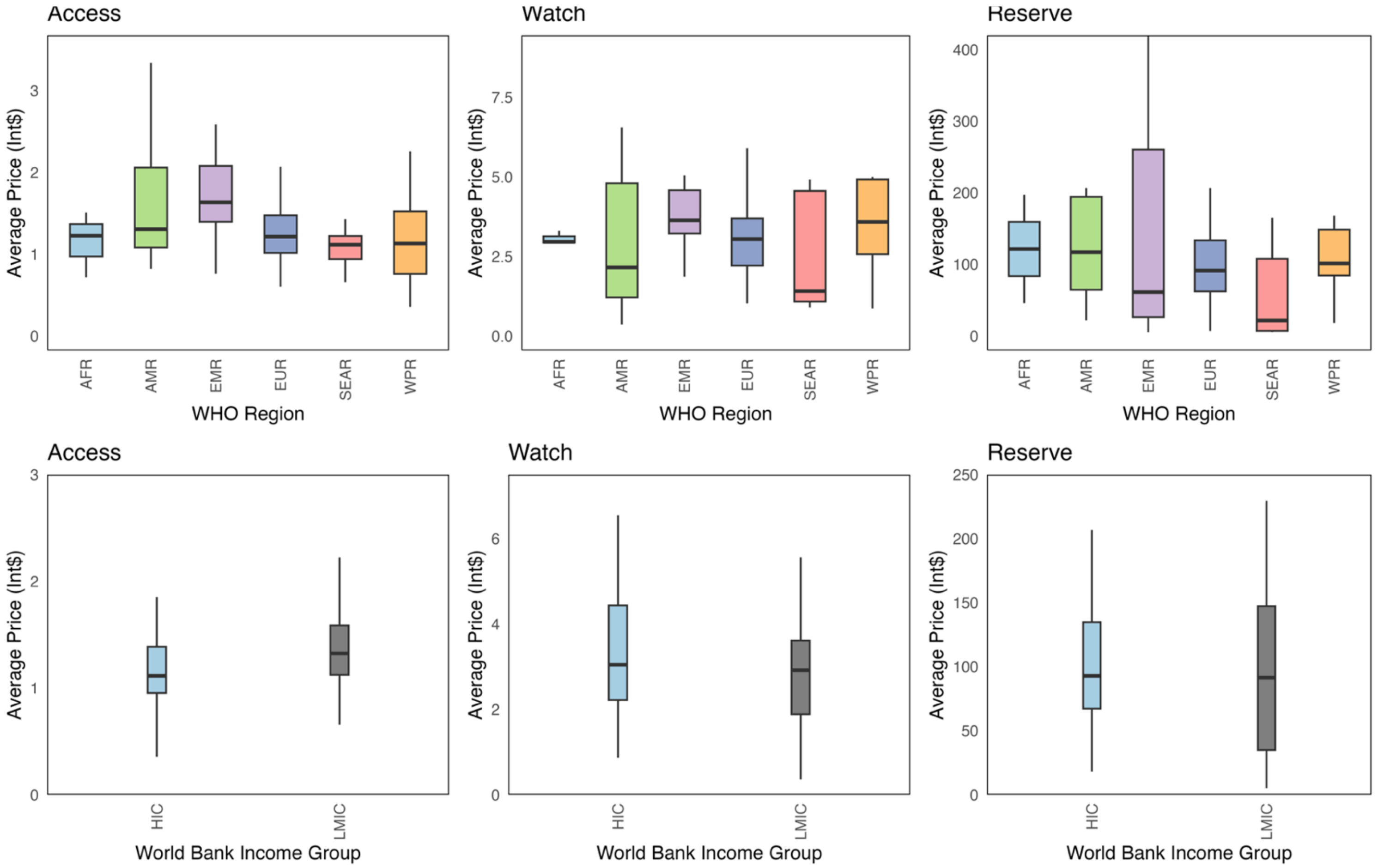

Notes: LMIC= Low-and-middle income countries (here it is just middle-income countries as there is no low-income countries). HIC= High-income countries. WHO= World Health Organization. AFRO= African region, AMRO= Americas region, EMRO= East Mediterranean region, EURO= Europe, SEARO= Southeast Asia region, WPRO= Western-pacific region. Authors' analysis of IQVIA MIDAS Quarterly Sales data for the year 2019, reflecting estimates of real-world activity. Copyright IQVIA. All Rights Reserved.

**Text A8.** Contribution of ex-manufacturer volume and antibiotic type to price variation

To quantify the contribution of ex-manufacturer volume and antibiotic composition to variation in antibiotic prices, we conducted a two-level decomposition using random forest models with impurity-based variable importance, implemented via the ranger package (used on R). At the first level (within AWaRe categories and by looking at antibiotics within countries), we modelled price per DDD across country–molecule combinations to assess the relative contribution of molecule identity (e.g., the name of the antibiotic per se) and volume purchased (log DDDs for each antibiotic) in explaining price variation for all antibiotics. At the second level (across AWaRe categories), we analysed country-level average price per AWaRe class to estimate the contributions of total ex-manufacturer volume per class and AWaRe category membership.

Variable importance was measured using the Mean Decrease in Impurity (MDI), calculated as the cumulative reduction in residual sum of squares (RSS) across all trees and decision nodes where a given variable was used for splitting. Higher scores indicate stronger predictive contributions to explaining price variation. Importance values were normalised to express each variable's percentage contribution to total model impurity reduction. All models were run using default hyperparameters.

We found that within AWaRe categories antibiotic type (mix) and volume (see figure below).

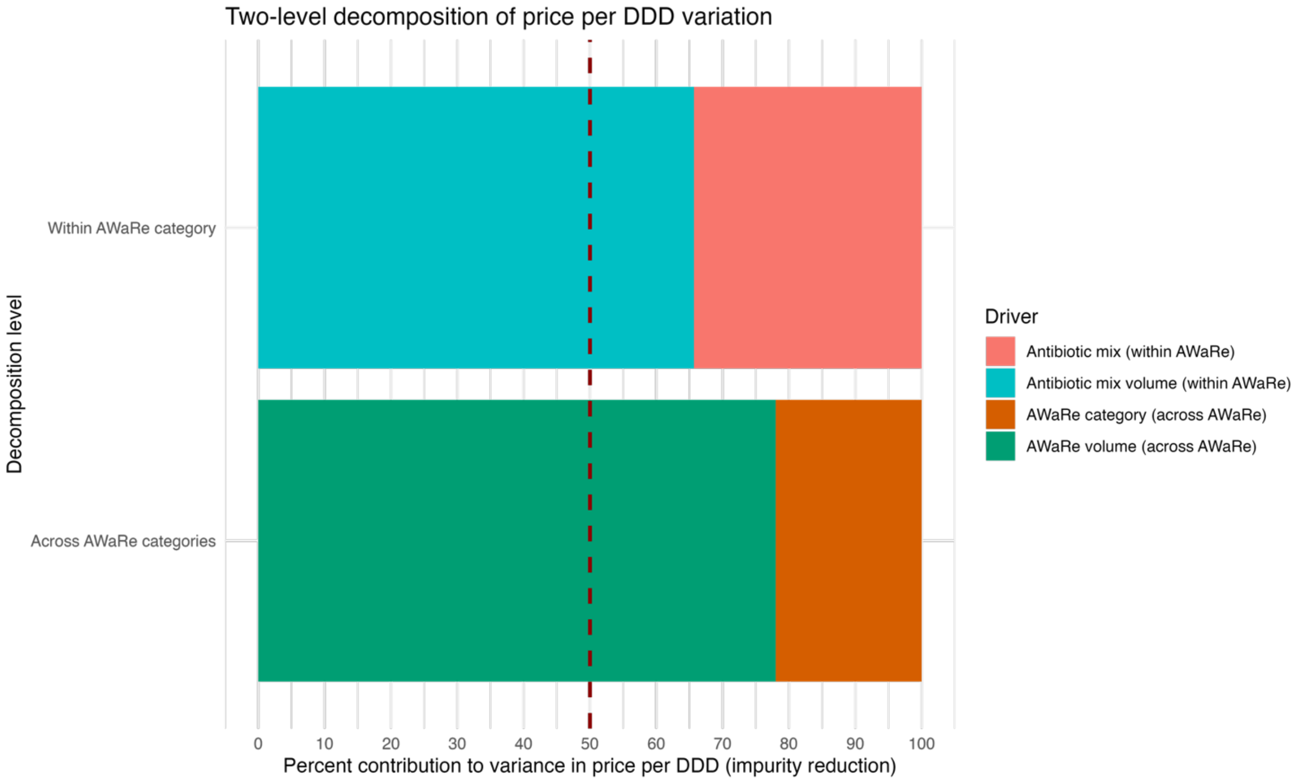

Notes: AWaRe (Access, Watch and Reserve). Authors' analysis of IQVIA MIDAS Quarterly Sales data for the year 2019, reflecting estimates of real-world activity. Copyright IQVIA. All Rights Reserved.

**Figure A7.** Access-to-Watch Price Ratio among countries, by country, data from the IQVIA MIDAS Quarterly sales data for the year 2019

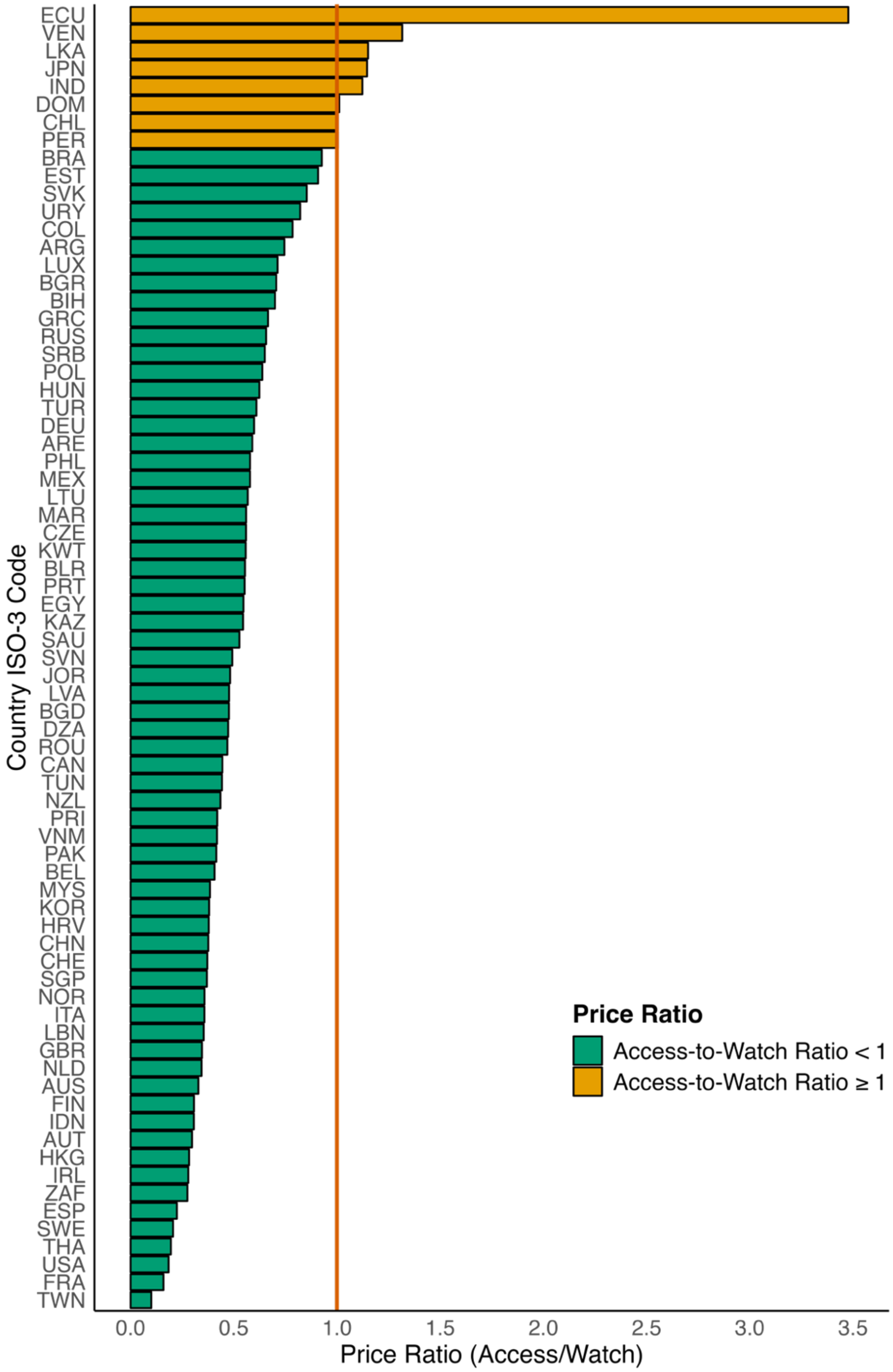

Notes: Full name for countries abbreviations are shown in Supplementary material. Table A1 indicates the abbreviations of country-ISO3-codes. Authors' analysis of IQVIA MIDAS Quarterly Sales data for the year 2019, reflecting estimates of real-world activity. Copyright IQVIA. All Rights Reserved.

**Table A9.** Access-to-Watch Price ratio among all antibiotics, by WHO-region and income level, data from the IQVIA MIDAS Quarterly sales data for the year 2019

| WHO region | Mean ratio | p25 | p75 | IQR |
|---|---|---|---|---|
| AFR | 0.397 | 0.359 | 0.458 | 0.099 |
| AMR | 0.978 | 0.578 | 1.005 | 0.426 |
| EMR | 0.501 | 0.449 | 0.558 | 0.109 |
| EUR | 0.503 | 0.358 | 0.635 | 0.277 |
| SEAR | 0.650 | 0.306 | 1.123 | 0.817 |
| WPR | 0.437 | 0.349 | 0.427 | 0.078 |
| **Income levels** | **Mean ratio** | **p25** | **p75** | **IQR** |
| HIC | 0.477 | 0.340 | 0.592 | 0.252 |
| UMC | 0.786 | 0.450 | 0.574 | 0.123 |
| LMC | 0.658 | 0.481 | 0.819 | 0.338 |

Notes: UMIC= Upper middle-income country. LMC= Lower middle income countries. HIC= High-income countries. WHO= World Health Organization. AFRO= African region, AMRO= Americas region, EMRO= East Mediterranean region, EURO= Europe, SEARO= Southeast Asia region, WPRO= Western-pacific region. p25= percentile 25$^{th}$ , p75= percentile 75$^{th}$ . IQR= Interquartile range. AMU= antibiotic usage. Authors' analysis of IQVIA MIDAS Quarterly Sales data for the year 2019, reflecting estimates of real-world activity. Copyright IQVIA. All Rights Reserved.

**Table A10.** Country volume-weighted ex-manufacturer prices (int$) per antibiotic across all sampled countries, data from the IQVIA MIDAS Quarterly sales data for the year 2019

| Antibiotic name | AWaRe category | Median | Percentile 25th | Percentile 75th | IQR |
|---|---|---|---|---|---|
| Amikacin | Access | 9.35 | 4.53 | 21.47 | 16.94 |
| Amoxicillin | Access | 0.67 | 0.49 | 0.90 | 0.41 |
| Amoxicillin+Clavulanic acid | Access | 2.07 | 1.33 | 3.11 | 1.78 |
| Ampicillin | Access | 4.24 | 1.21 | 9.83 | 8.62 |
| Ampicillin+Sulbactam | Access | 29.47 | 16.03 | 61.52 | 45.48 |
| Bacampicillin | Access | 0.60 | 0.51 | 0.60 | 0.08 |
| Benzylpenicillin | Access | 10.12 | 4.34 | 22.09 | 17.76 |
| Cefadroxil | Access | 2.22 | 1.51 | 3.35 | 1.84 |
| Cefalexin | Access | 1.66 | 1.19 | 2.25 | 1.06 |
| Cefalotin | Access | 10.92 | 5.69 | 16.98 | 11.28 |
| Cefapirin | Access | 11.27 | 11.27 | 11.27 | 0.00 |
| Cefatrizine | Access | 2.64 | 2.05 | 3.24 | 1.19 |
| Cefazedone | Access | 60.54 | 39.02 | 82.06 | 43.04 |
| Cefazolin | Access | 7.90 | 4.38 | 12.40 | 8.02 |
| Cefradine | Access | 1.67 | 1.18 | 2.83 | 1.65 |
| Cefroxadine | Access | 5.34 | 5.16 | 5.52 | 0.35 |
| Ceftezole | Access | 10.26 | 8.44 | 12.37 | 3.93 |
| Chloramphenicol | Access | 2.77 | 1.16 | 9.10 | 7.94 |
| Clindamycin | Access | 3.58 | 2.35 | 5.33 | 2.98 |
| Cloxacillin | Access | 1.88 | 0.88 | 3.91 | 3.03 |
| Dicloxacillin | Access | 0.89 | 0.85 | 3.02 | 2.17 |
| Doxycycline | Access | 0.33 | 0.18 | 0.49 | 0.31 |
| Flucloxacillin | Access | 2.54 | 1.58 | 8.37 | 6.79 |
| Furazidin | Access | 2.40 | 1.61 | 2.57 | 0.96 |
| Gentamicin | Access | 2.81 | 1.46 | 8.23 | 6.77 |
| Mecillinam | Access | 27.61 | 23.60 | 31.62 | 8.02 |
| Metronidazole | Access | 5.95 | 2.77 | 11.61 | 8.83 |
| Nafcillin | Access | 24.11 | 11.89 | 35.71 | 23.82 |
| Nifurtoinol | Access | 1.40 | 0.80 | 2.00 | 1.20 |
| Nitrofurantoin | Access | 0.60 | 0.44 | 1.16 | 0.71 |
| Ornidazole | Access | 3.22 | 1.77 | 4.91 | 3.14 |
| Oxacillin | Access | 3.68 | 1.48 | 6.98 | 5.49 |
| Phenoxymethylpenicillin | Access | 1.02 | 0.58 | 1.57 | 0.99 |
| Pivampicillin | Access | 3.76 | 3.76 | 3.76 | 0.00 |
| Pivmecillinam | Access | 1.61 | 1.42 | 1.81 | 0.39 |
| Secnidazole | Access | 5.15 | 3.61 | 9.29 | 5.68 |
| Spectinomycin | Access | 14.56 | 9.55 | 22.12 | 12.57 |
| Sulbactam | Access | 11.33 | 8.40 | 12.08 | 3.68 |
| Sulfadiazine | Access | 0.49 | 0.11 | 2.76 | 2.65 |
| Sulfadiazine+Trimethoprim | Access | 0.42 | 0.42 | 0.42 | 0.00 |
| Sulfadimethoxine | Access | 0.10 | 0.10 | 0.10 | 0.00 |
| Sulfadimidine | Access | 0.32 | 0.17 | 0.60 | 0.43 |

| | | | | | |
|---|---|---|---|---|---|
| Sulfalene | Access | 0.05 | 0.05 | 0.05 | 0.00 |
| Sulfamethizole | Access | 6.01 | 4.75 | 7.26 | 2.51 |
| Sulfamethoxazole | Access | 0.36 | 0.36 | 0.36 | 0.00 |
| Sulfamethoxazole+Trimethoprim | Access | 0.75 | 0.46 | 1.05 | 0.59 |
| Sulfamethoxypyridazine | Access | 0.16 | 0.16 | 0.16 | 0.00 |
| Sulfametrole+Trimethoprim | Access | 0.65 | 0.60 | 0.69 | 0.09 |
| Sulfapyridine | Access | 0.05 | 0.05 | 0.05 | 0.00 |
| Tetracycline | Access | 0.47 | 0.28 | 1.07 | 0.79 |
| Thiamphenicol | Access | 3.10 | 1.27 | 7.11 | 5.84 |
| Tinidazole | Access | 3.60 | 1.57 | 9.21 | 7.63 |
| Trimethoprim | Access | 0.70 | 0.40 | 1.41 | 1.01 |
| Arbekacin | Watch | 46.14 | 38.95 | 74.86 | 35.91 |
| Azithromycin | Watch | 1.53 | 0.99 | 2.05 | 1.06 |
| Azlocillin | Watch | 18.99 | 18.99 | 18.99 | 0.00 |
| Biapenem | Watch | 79.09 | 63.56 | 141.44 | 77.88 |
| Carbenicillin | Watch | 48.01 | 48.01 | 48.01 | 0.00 |
| Cefaclor | Watch | 2.49 | 1.80 | 3.61 | 1.81 |
| Cefamandole | Watch | 48.79 | 26.20 | 52.82 | 26.62 |
| Cefbuperazone | Watch | 16.61 | 16.61 | 16.61 | 0.00 |
| Cefcapene-pivoxil | Watch | 1.53 | 1.49 | 1.57 | 0.08 |
| Cefdinir | Watch | 6.18 | 2.91 | 9.45 | 6.54 |
| Cefditoren-pivoxil | Watch | 3.40 | 2.84 | 6.45 | 3.61 |
| Cefepime | Watch | 41.56 | 25.16 | 65.77 | 40.61 |
| Cefetamet-pivoxil | Watch | 2.68 | 2.48 | 18.66 | 16.18 |
| Cefixime | Watch | 3.79 | 2.20 | 5.15 | 2.95 |
| Cefmenoxime | Watch | 18.72 | 18.15 | 19.30 | 1.15 |
| Cefmetazole | Watch | 27.42 | 25.31 | 29.24 | 3.93 |
| Cefminox | Watch | 36.36 | 19.90 | 50.11 | 30.20 |
| Cefodizime | Watch | 16.65 | 16.21 | 16.97 | 0.76 |
| Cefonicid | Watch | 9.15 | 6.73 | 11.56 | 4.83 |
| Cefoperazone | Watch | 22.98 | 14.34 | 35.39 | 21.05 |
| Ceforanide | Watch | 31.64 | 31.64 | 31.64 | 0.00 |
| Cefotaxime | Watch | 15.58 | 8.55 | 30.98 | 22.43 |
| Cefotetan | Watch | 38.45 | 31.09 | 40.41 | 9.32 |
| Cefotiam | Watch | 28.85 | 27.36 | 29.53 | 2.17 |
| Cefoxitin | Watch | 33.42 | 27.18 | 71.77 | 44.59 |
| Cefozopran | Watch | 42.88 | 42.88 | 42.88 | 0.00 |
| Cefpiramide | Watch | 14.74 | 14.61 | 14.88 | 0.28 |
| Cefpirome | Watch | 66.88 | 34.91 | 130.66 | 95.75 |
| Cefpodoxime-proxetil | Watch | 4.26 | 2.74 | 6.15 | 3.41 |
| Cefprozil | Watch | 3.60 | 2.73 | 5.61 | 2.88 |
| Cefsulodin | Watch | 59.24 | 59.24 | 59.24 | 0.00 |
| Ceftazidime | Watch | 23.07 | 14.78 | 39.19 | 24.41 |
| Cefteram-pivoxil | Watch | 4.87 | 3.22 | 6.52 | 3.30 |
| Ceftibuten | Watch | 4.21 | 3.10 | 5.00 | 1.89 |
| Ceftizoxime | Watch | 49.82 | 25.39 | 63.99 | 38.60 |

| | | | | | |
|---|---|---|---|---|---|
| Ceftriaxone | Watch | 11.48 | 4.29 | 21.90 | 17.61 |
| Cefuroxime | Watch | 10.87 | 7.32 | 18.09 | 10.77 |
| Cefuroxime axetil | Watch | 1.46 | 0.92 | 2.40 | 1.48 |
| Chlortetracycline | Watch | 1.46 | 1.46 | 1.46 | 0.00 |
| Cinoxacin | Watch | 1.30 | 1.30 | 1.30 | 0.00 |
| Ciprofloxacin | Watch | 1.44 | 0.91 | 2.77 | 1.86 |
| Clarithromycin | Watch | 1.32 | 0.90 | 1.90 | 1.00 |
| Delafloxacin | Watch | 135.91 | 135.91 | 135.91 | 0.00 |
| Demeclocycline | Watch | 7.80 | 1.70 | 26.44 | 24.74 |
| Dibekacin | Watch | 16.42 | 12.01 | 20.83 | 8.82 |
| Dirithromycin | Watch | 1.57 | 1.48 | 1.65 | 0.16 |
| Doripenem | Watch | 177.84 | 81.12 | 242.54 | 161.42 |
| Enoxacin | Watch | 2.06 | 1.99 | 2.13 | 0.15 |
| Ertapenem | Watch | 72.30 | 55.16 | 97.87 | 42.71 |
| Erythromycin | Watch | 1.08 | 0.66 | 2.07 | 1.41 |
| Fidaxomicin | Watch | 198.86 | 171.20 | 286.06 | 114.87 |
| Fleroxacin | Watch | 0.86 | 0.86 | 0.86 | 0.00 |
| Flomoxef | Watch | 36.35 | 26.70 | 50.18 | 23.48 |
| Flumequine | Watch | 1.17 | 1.17 | 1.17 | 0.00 |
| Fosfomycin | Watch | 16.00 | 7.17 | 25.34 | 18.17 |
| Fusidic acid | Watch | 6.82 | 5.70 | 12.64 | 6.95 |
| Garenoxacin | Watch | 3.85 | 3.56 | 4.13 | 0.57 |
| Gatifloxacin | Watch | 1.73 | 0.27 | 6.01 | 5.75 |
| Gemifloxacin | Watch | 5.99 | 3.89 | 7.21 | 3.32 |
| Isepamicin | Watch | 15.50 | 7.07 | 15.59 | 8.52 |
| Josamycin | Watch | 5.51 | 2.91 | 6.19 | 3.28 |
| Kanamycin | Watch | 1.85 | 0.76 | 3.06 | 2.30 |
| Latamoxef | Watch | 56.83 | 46.82 | 66.85 | 20.03 |
| Levofloxacin | Watch | 2.36 | 1.56 | 3.48 | 1.92 |
| Lincomycin | Watch | 4.26 | 1.96 | 7.47 | 5.51 |
| Lomefloxacin | Watch | 1.70 | 0.67 | 2.77 | 2.09 |
| Lymecycline | Watch | 1.31 | 0.65 | 2.98 | 2.33 |
| Meropenem | Watch | 65.81 | 43.11 | 121.71 | 78.59 |
| Metacycline | Watch | 1.20 | 1.03 | 1.38 | 0.35 |
| Mezlocillin | Watch | 23.20 | 18.87 | 27.54 | 8.67 |
| Midecamycin | Watch | 2.22 | 1.62 | 2.41 | 0.79 |
| Minocycline | Watch | 1.42 | 0.88 | 2.36 | 1.49 |
| Moxifloxacin | Watch | 5.09 | 2.97 | 7.35 | 4.38 |
| Neomycin | Watch | 10.61 | 5.24 | 16.14 | 10.90 |
| Netilmicin | Watch | 18.18 | 9.82 | 46.11 | 36.29 |
| Norfloxacin | Watch | 0.86 | 0.56 | 1.29 | 0.73 |
| Ofloxacin | Watch | 1.30 | 0.85 | 2.26 | 1.41 |
| Oxytetracycline | Watch | 0.28 | 0.16 | 0.67 | 0.52 |
| Pazufloxacin | Watch | 22.15 | 20.81 | 22.64 | 1.83 |
| Pefloxacin | Watch | 2.25 | 1.19 | 3.31 | 2.12 |
| Pheneticillin | Watch | 3.02 | 3.02 | 3.02 | 0.00 |

| | | | | | |
|---|---|---|---|---|---|
| Pipemidic acid | Watch | 1.43 | 0.65 | 2.02 | 1.36 |
| Piperacillin | Watch | 31.24 | 20.48 | 68.53 | 48.05 |
| Piperacillin+Tazobactam | Watch | 51.40 | 30.37 | 89.83 | 59.46 |
| Pristinamycin | Watch | 6.51 | 5.43 | 8.61 | 3.18 |
| Prulifloxacin | Watch | 3.86 | 3.10 | 8.61 | 5.51 |
| Ribostamycin | Watch | 10.32 | 5.71 | 14.94 | 9.23 |
| Rifamycin | Watch | 10.56 | 8.64 | 29.78 | 21.14 |
| Rifaximin | Watch | 4.72 | 2.55 | 6.86 | 4.32 |
| Roxithromycin | Watch | 1.22 | 0.57 | 1.86 | 1.28 |
| Rufloxacin | Watch | 6.99 | 4.54 | 9.43 | 4.89 |
| Sarecycline | Watch | 25.47 | 25.47 | 25.47 | 0.00 |
| Sitafloxacin | Watch | 8.29 | 5.16 | 11.42 | 6.27 |
| Sparfloxacin | Watch | 0.64 | 0.45 | 1.02 | 0.57 |
| Spiramycin | Watch | 3.47 | 2.25 | 5.10 | 2.85 |
| Streptomycin | Watch | 1.75 | 0.65 | 5.60 | 4.95 |
| Sulbenicillin | Watch | 66.40 | 66.40 | 66.40 | 0.00 |
| Tebipenem | Watch | 25.48 | 25.48 | 25.48 | 0.00 |
| Teicoplanin | Watch | 66.67 | 42.67 | 93.09 | 50.42 |
| Telithromycin | Watch | 6.97 | 5.92 | 7.30 | 1.37 |
| Temocillin | Watch | 158.88 | 120.74 | 215.56 | 94.82 |
| Ticarcillin | Watch | 35.82 | 33.96 | 37.67 | 3.70 |
| Tobramycin | Watch | 12.20 | 8.52 | 25.65 | 17.13 |
| Tosufloxacin | Watch | 2.42 | 2.28 | 3.06 | 0.79 |
| Vancomycin | Watch | 29.18 | 17.68 | 51.80 | 34.11 |
| Aztreonam | Reserve | 89.99 | 48.12 | 137.26 | 89.14 |
| Ceftaroline-fosamil | Reserve | 183.52 | 149.64 | 227.29 | 77.65 |
| Ceftazidime+Avibactam | Reserve | 775.14 | 536.07 | 944.05 | 407.98 |
| Ceftobiprole-medocaril | Reserve | 244.37 | 176.98 | 264.44 | 87.47 |
| Ceftolozane+Tazobactam | Reserve | 1264.14 | 918.52 | 1462.10 | 543.59 |
| Colistin | Reserve | 88.91 | 48.49 | 149.85 | 101.36 |
| Dalbavancin | Reserve | 3181.89 | 2764.65 | 3695.16 | 930.52 |
| Daptomycin | Reserve | 95.28 | 77.18 | 133.85 | 56.67 |
| Eravacycline | Reserve | 148.58 | 148.58 | 148.58 | 0.00 |
| Faropenem | Reserve | 8.89 | 5.58 | 8.91 | 3.32 |
| Fosfomycin | Reserve | 75.36 | 58.47 | 129.96 | 71.49 |
| Lefamulin | Reserve | 263.90 | 263.90 | 263.90 | 0.00 |
| Linezolid | Reserve | 83.45 | 32.00 | 142.39 | 110.39 |
| Meropenem+Vaborbactam | Reserve | 423.57 | 401.90 | 534.77 | 132.87 |
| Minocycline | Reserve | 3.87 | 3.87 | 3.87 | 0.00 |
| Omadacycline | Reserve | 342.69 | 342.69 | 342.69 | 0.00 |
| Oritavancin | Reserve | 2373.14 | 2373.14 | 2373.14 | 0.00 |
| Plazomicin | Reserve | 783.62 | 783.62 | 783.62 | 0.00 |
| Polymyxin B | Reserve | 112.54 | 25.18 | 254.14 | 228.97 |
| Tedizolid | Reserve | 230.19 | 186.89 | 326.45 | 139.56 |
| Telavancin | Reserve | 364.18 | 347.69 | 380.66 | 32.97 |
| Tigecycline | Reserve | 164.77 | 129.99 | 207.00 | 77.00 |

Notes: AWaRe =Access, Watch and Reserve. IQR= Interquartile range (percentile 75th – percentile 25th) . int$= International prices, 2019. Authors' analysis of IQVIA MIDAS Quarterly Sales data for the year 2019, reflecting estimates of real-world activity. Copyright IQVIA. All Rights Reserved.

**Table A11.** Ex-manufacturer prices (int$) per antibiotic DDD across AWaRe categories and route of administration, by WHO region and income-group, data from the IQVIA MIDAS Quarterly sales data for the year 2019

| Group | Route of administration | Aware category | Volume-weighted | p25 | Median | p75 |
|---|---|---|---|---|---|---|
| **WHO region** | | | | | | |
| AFR | Oral | Access | 0.031 | 0.016 | 0.033 | 0.093 |
| AFR | Oral | Watch | 0.068 | 0.033 | 0.082 | 0.190 |
| AFR | Oral | Reserve | 1.102 | 1.525 | 6.180 | 8.916 |
| AFR | Parenteral | Access | 0.261 | 0.089 | 0.184 | 0.633 |
| AFR | Parenteral | Watch | 1.181 | 0.451 | 1.357 | 2.997 |
| AFR | Parenteral | Reserve | 9.970 | 4.195 | 7.595 | 11.336 |
| AMR | Oral | Access | 0.562 | 0.054 | 0.357 | 1.207 |
| AMR | Oral | Watch | 1.979 | 0.081 | 0.477 | 1.885 |
| AMR | Oral | Reserve | 26.052 | 5.675 | 13.254 | 148.586 |
| AMR | Parenteral | Access | 9.014 | 0.758 | 6.866 | 18.631 |
| AMR | Parenteral | Watch | 12.808 | 2.891 | 9.469 | 26.546 |
| AMR | Parenteral | Reserve | 112.862 | 24.957 | 73.449 | 203.778 |
| EMR | Oral | Access | 0.070 | 0.012 | 0.028 | 0.173 |
| EMR | Oral | Watch | 0.127 | 0.010 | 0.018 | 0.107 |
| EMR | Oral | Reserve | 0.105 | 0.035 | 0.041 | 0.054 |
| EMR | Parenteral | Access | 0.223 | 0.057 | 0.135 | 0.361 |
| EMR | Parenteral | Watch | 0.647 | 0.143 | 0.236 | 0.648 |
| EMR | Parenteral | Reserve | 5.954 | 0.479 | 1.277 | 25.238 |
| EUR | Oral | Access | 0.541 | 0.166 | 0.586 | 1.417 |
| EUR | Oral | Watch | 0.896 | 0.272 | 0.921 | 1.851 |
| EUR | Oral | Reserve | 60.099 | 3.977 | 40.394 | 95.734 |
| EUR | Parenteral | Access | 5.124 | 0.611 | 5.423 | 13.292 |
| EUR | Parenteral | Watch | 11.518 | 1.253 | 11.101 | 36.982 |
| EUR | Parenteral | Reserve | 81.307 | 10.723 | 62.865 | 132.319 |
| SEAR | Oral | Access | 0.065 | 0.001 | 0.022 | 0.060 |
| SEAR | Oral | Watch | 0.031 | 0.006 | 0.014 | 0.030 |
| SEAR | Oral | Reserve | 0.062 | 0.031 | 0.088 | 0.117 |
| SEAR | Parenteral | Access | 0.361 | 0.012 | 0.099 | 0.440 |
| SEAR | Parenteral | Watch | 1.046 | 0.021 | 0.252 | 0.858 |
| SEAR | Parenteral | Reserve | 7.501 | 1.144 | 2.896 | 3.704 |
| WPR | Oral | Access | 0.084 | 0.009 | 0.047 | 0.119 |
| WPR | Oral | Watch | 0.130 | 0.008 | 0.040 | 0.137 |
| WPR | Oral | Reserve | 1.984 | 0.066 | 0.985 | 15.110 |
| WPR | Parenteral | Access | 1.092 | 0.084 | 0.296 | 1.423 |
| WPR | Parenteral | Watch | 2.733 | 0.166 | 0.743 | 3.073 |
| WPR | Parenteral | Reserve | 11.108 | 0.858 | 3.729 | 19.937 |
| **Income level** | | | | | | |
| HIC | Oral | Access | 0.634 | 0.196 | 0.637 | 1.587 |
| HIC | Oral | Watch | 1.631 | 0.073 | 0.824 | 1.991 |
| HIC | Oral | Reserve | 27.720 | 5.662 | 46.454 | 107.649 |
| HIC | Parenteral | Access | 7.332 | 2.639 | 8.986 | 19.591 |

| | | | | | | |
|---|---|---|---|---|---|---|
| HIC | Parenteral | Watch | 15.161 | 2.051 | 14.397 | 37.731 |
| HIC | Parenteral | Reserve | 97.271 | 28.087 | 83.941 | 158.564 |
| LMC | Oral | Access | 0.018 | 0.004 | 0.018 | 0.035 |
| LMC | Oral | Watch | 0.016 | 0.007 | 0.016 | 0.033 |
| LMC | Oral | Reserve | 0.053 | 0.032 | 0.070 | 0.113 |
| LMC | Parenteral | Access | 0.079 | 0.044 | 0.115 | 0.318 |
| LMC | Parenteral | Watch | 0.148 | 0.082 | 0.239 | 0.678 |
| LMC | Parenteral | Reserve | 1.548 | 1.005 | 2.694 | 3.719 |
| UMC | Oral | Access | 0.163 | 0.036 | 0.097 | 0.316 |
| UMC | Oral | Watch | 0.206 | 0.048 | 0.147 | 0.357 |
| UMC | Oral | Reserve | 4.123 | 1.028 | 1.687 | 12.158 |
| UMC | Parenteral | Access | 1.259 | 0.115 | 0.510 | 1.795 |
| UMC | Parenteral | Watch | 2.659 | 0.359 | 1.601 | 4.281 |
| UMC | Parenteral | Reserve | 11.993 | 2.197 | 7.334 | 29.355 |

Notes: WHO= World Health Organization. AFRO= African region, AMRO= Americas region, EMRO= East Mediterranean region, EURO= Europe, SEARO= Southeast Asia region, WPRO= Western-pacific region. HIC= High-income country, UMC= Upper middle-income country. LMC= Lower middle-income country. p25= percentile 25th , p75= percentile 75th. Authors' analysis of IQVIA MIDAS Quarterly Sales data for the year 2019, reflecting estimates of real-world activity. Copyright IQVIA. All Rights Reserved.

- Oral and parenteral ABU distributions varied by country (Figure A9), with parenteral Access antibiotics being much pricier in HICs (10·6x), UMCs (6·7x), and LMCs (3·4x), compared to oral (Table A12). Watch parenteral antibiotics were 8·2-11·9 times higher, with Reserve antibiotics in LMCs having a 28·2 times higher price compared to 2·5-1·9 times in HICs and UMCs.

**Table A12.** Ex Overall price per DDD by AWaRe category and route of administration, data from the IQVIA MIDAS Quarterly sales data for the year 2019

| Antibiotic route of administration | Aware category | Median | p25 | p75 | IQR |
|---|---|---|---|---|---|
| Oral | Access | 1.02 | 0.75 | 1.37 | 0.62 |
| Oral | Reserve | 68.10 | 26.43 | 142.19 | 115.76 |
| Oral | Watch | 1.48 | 1.17 | 2.07 | 0.90 |
| Parenteral | Access | 8.36 | 5.48 | 12.42 | 6.94 |
| Parenteral | Reserve | 121.43 | 83.16 | 167.83 | 84.67 |
| Parenteral | Watch | 21.71 | 11.35 | 33.01 | 21.66 |

Notes: IQR= Interquartile range. Authors' analysis of IQVIA MIDAS Quarterly Sales data for the year 2019, reflecting estimates of real-world activity. Copyright IQVIA. All Rights Reserved.

**Figure A8.** Proportion of total antibiotic usage by administration route (oral or parenteral), by country, data from the IQVIA MIDAS Quarterly sales data for the year 2019

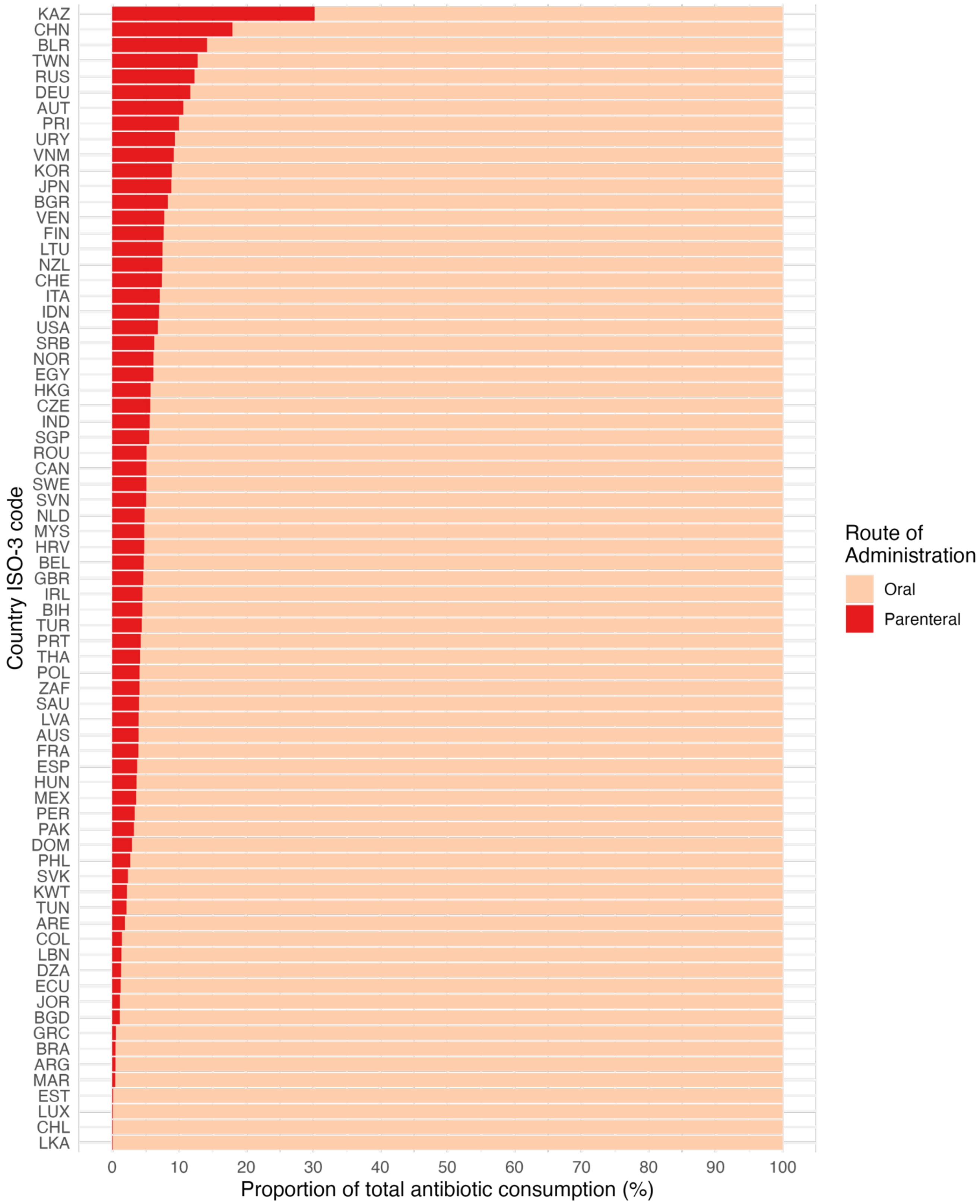

Notes: Table A1 indicates the abbreviations of country-ISO3-codes. Authors' analysis of IQVIA MIDAS Quarterly Sales data for the year 2019, reflecting estimates of real-world activity. Copyright IQVIA. All Rights Reserved.

**Figure A9**. (A) Antibiotic expenditure per capita (ex-manufacturer volume) and (B) number of unique antibiotics (chemical compounds) sold by AWaRe categories, by country (n=73 countries), data from the IQVIA MIDAS Quarterly sales data for the year 2019

A.

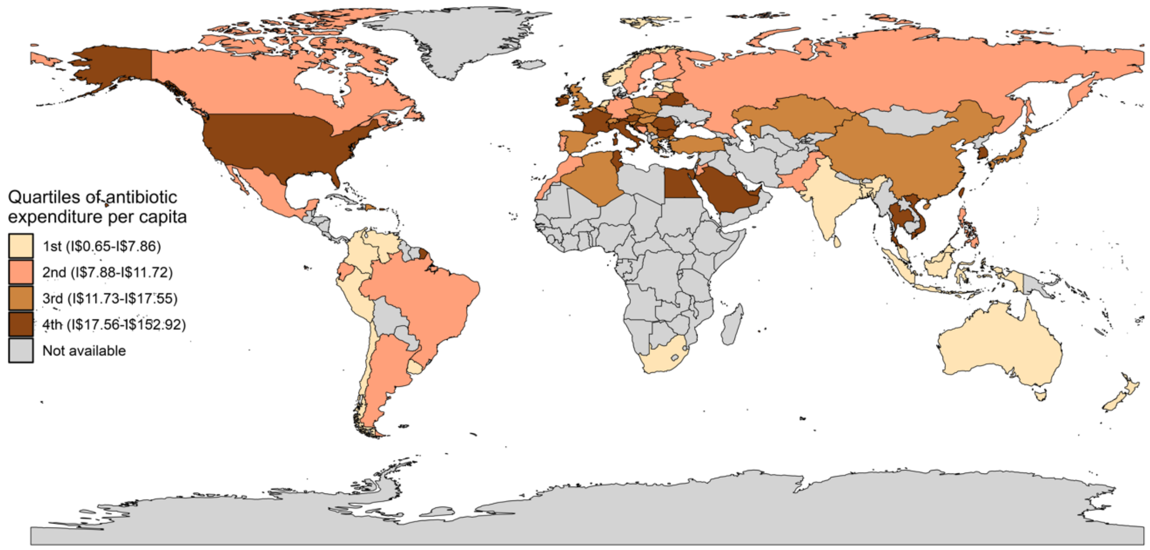

B.

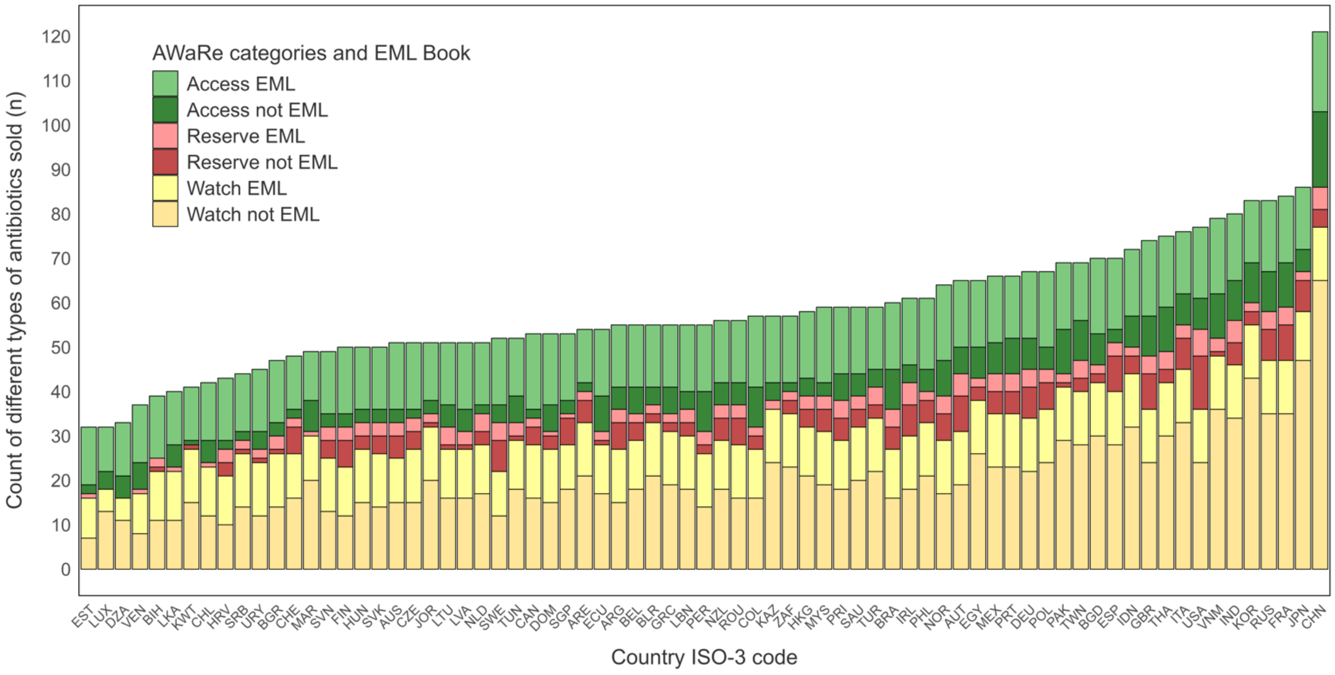

Notes: AWaRe= Access, Watch and Reserve. EML= Essential Medicine list. I$= International dollars, 2019. Country ISO-3 code come from the International Standard for country codes, see Supplementary Table A1 for full list. Authors' analysis of IQVIA MIDAS Quarterly Sales data for the year 2019, reflecting estimates of real-world activity. Copyright IQVIA. All Rights Reserved.

**Table A13.** Net potential country savings per capita if Access consumption threshold is increased to 60%, 65%, 70%, 75% or 80% considering 2019 ABU volume by country, according to ex-manufacturer prices (n=65 countries)*, data from the IQVIA MIDAS Quarterly sales data for the year 2019

| Country/ access % | Net potential antibiotic expenditure savings ex-manufacturer per capita, (Int$) | | | | |
|---|---|---|---|---|---|
| | 80% | 75% | 70% | 65% | 60% |
| ECU | | | | | |
| JPN | | | | | |
| IND | | | | | |
| LKA | | | | | |
| DOM | | | | | |
| VEN | | | | | |
| PER | | | | | |
| CHL | | | | | |
| NZL | 0.069 | | | | |
| EST | 0.092 | 0.071 | 0.051 | 0.031 | 0.011 |
| BRA | 0.112 | 0.080 | 0.048 | 0.017 | |
| COL | 0.135 | 0.085 | 0.034 | | |
| URY | 0.237 | 0.169 | 0.101 | 0.032 | |
| DZA | 0.294 | | | | |
| AUS | 0.363 | | | | |
| LUX | 0.365 | 0.275 | 0.185 | 0.095 | 0.005 |
| ARG | 0.391 | 0.260 | 0.128 | | |
| MAR | 0.442 | 0.151 | | | |
| LTU | 0.480 | 0.205 | | | |
| LVA | 0.527 | 0.236 | | | |
| NLD | 0.547 | 0.214 | | | |
| ZAF | 0.642 | 0.060 | | | |
| BIH | 0.657 | 0.450 | 0.242 | 0.035 | |
| SVN | 0.736 | 0.161 | | | |
| SVK | 0.736 | 0.644 | 0.552 | 0.459 | 0.367 |
| IDN | 0.954 | 0.747 | 0.539 | 0.331 | 0.123 |
| CAN | 1.006 | 0.620 | 0.233 | | |
| DEU | 1.032 | 0.803 | 0.574 | 0.345 | 0.116 |
| PRT | 1.198 | 0.870 | 0.543 | 0.215 | |
| RUS | 1.228 | 1.035 | 0.843 | 0.651 | 0.459 |
| CZE | 1.231 | 0.893 | 0.555 | 0.216 | |
| SGP | 1.243 | 0.920 | 0.598 | 0.276 | |
| MYS | 1.296 | 0.987 | 0.679 | 0.370 | 0.062 |
| POL | 1.299 | 0.992 | 0.685 | 0.379 | 0.072 |
| FIN | 1.408 | 0.685 | | | |

| | | | | | |
|---|---|---|---|---|---|
| MEX | 1.432 | 1.183 | 0.935 | 0.687 | 0.439 |
| TUN | 1.477 | 0.553 | | | |
| SRB | 1.594 | 1.260 | 0.926 | 0.592 | 0.257 |
| HUN | 1.606 | 1.334 | 1.062 | 0.789 | 0.517 |
| PHL | 1.610 | 1.363 | 1.117 | 0.870 | 0.624 |
| JOR | 1.655 | 1.348 | 1.040 | 0.733 | 0.426 |
| BGD | 1.770 | 1.568 | 1.366 | 1.164 | 0.962 |
| NOR | 1.791 | 1.230 | 0.669 | 0.108 | |
| SWE | 1.798 | 0.958 | 0.119 | | |
| GRC | 1.825 | 1.577 | 1.328 | 1.079 | 0.831 |
| KAZ | 1.988 | 1.610 | 1.232 | 0.854 | 0.476 |
| GBR | 1.999 | 1.289 | 0.579 | | |
| KWT | 2.007 | 1.736 | 1.466 | 1.196 | 0.926 |
| TUR | 2.044 | 1.669 | 1.294 | 0.920 | 0.545 |
| BLR | 2.063 | 1.585 | 1.106 | 0.627 | 0.149 |
| HKG | 2.235 | 1.804 | 1.374 | 0.943 | 0.512 |
| CHE | 2.353 | 1.738 | 1.122 | 0.506 | |
| BEL | 2.399 | 1.728 | 1.058 | 0.388 | |
| THA | 2.525 | 0.870 | | | |
| BGR | 2.548 | 2.183 | 1.818 | 1.453 | 1.088 |
| PAK | 2.588 | 2.189 | 1.790 | 1.391 | 0.992 |
| IRL | 2.886 | 1.742 | 0.599 | | |
| ESP | 3.502 | 2.490 | 1.478 | 0.465 | |
| FRA | 3.551 | 1.435 | | | |
| ROU | 3.748 | 3.053 | 2.359 | 1.664 | 0.969 |
| USA | 4.141 | 2.314 | 0.487 | | |
| LBN | 4.180 | 3.185 | 2.190 | 1.194 | 0.199 |
| CHN | 4.409 | 3.962 | 3.516 | 3.070 | 2.623 |
| VNM | 5.000 | 4.164 | 3.327 | 2.491 | 1.654 |
| ARE | 5.853 | 5.056 | 4.260 | 3.464 | 2.667 |
| EGY | 5.859 | 4.930 | 4.000 | 3.071 | 2.142 |
| AUT | 6.126 | 4.407 | 2.689 | 0.970 | |
| SAU | 6.633 | 5.579 | 4.526 | 3.472 | 2.419 |
| ITA | 6.744 | 5.708 | 4.672 | 3.636 | 2.600 |
| KOR | 6.744 | 5.800 | 4.855 | 3.911 | 2.967 |
| PRI | 7.599 | 6.321 | 5.042 | 3.764 | 2.486 |
| TWN | 8.032 | 4.806 | 1.581 | | |
| HRV | 24.435 | 17.276 | 10.118 | 2.960 | |

Notes: *Omitted countries presented (n=8) relatively higher volume-weighted prices (ex-manufacturer) for access antibiotics compared to watch, reason why net savings might not be expected. ($) are presented in international dollars PPP-adjusted, 2019. Blank spaces refer to those countries having either higher Access prices compared to Watch or those which respective Access % (threshold) was fulfilled already; hence no cost savings were experienced. Int$: International dollars, 2019. Table A1 indicates the abbreviations of country-ISO3-codes. Authors' analysis of IQVIA MIDAS Quarterly Sales data for the year 2019, reflecting estimates of real-world activity. Copyright IQVIA. All Rights Reserved.

**Table A14.** Net potential country antibiotic expenditure savings if Access consumption threshold is increased to 60%, 65%, 70%, 75% or 80% considering 2019 ABU volume by country, according to ex-manufacturer prices (n=65 countries)*, data from the IQVIA MIDAS Quarterly sales data for the year 2019

| Country/ access % | Net potential savings ex-manufacturer, (Int$) | | | | |
|---|---|---|---|---|---|
| | 80% | 75% | 70% | 65% | 60% |
| ECU | | | | | |
| JPN | | | | | |
| IND | | | | | |
| LKA | | | | | |
| DOM | | | | | |
| VEN | | | | | |
| PER | | | | | |
| CHL | | | | | |
| NZL | 349,027 | - | - | - | - |
| EST | 121,673 | 94,907 | 68,141 | 41,375 | 14,609 |
| BRA | 23,900,969 | 17,115,254 | 10,329,540 | 3,543,825 | - |
| COL | 6,892,267 | 4,315,582 | 1,738,897 | - | - |
| URY | 813,082 | 579,089 | 345,096 | 111,103 | - |
| DZA | 12,762,428 | - | - | - | - |
| AUS | 9,299,650 | - | - | - | - |
| LUX | 230,202 | 173,475 | 116,748 | 60,020 | 3,293 |
| ARG | 17,750,807 | 11,777,821 | 5,804,834 | - | - |
| MAR | 16,198,284 | 5,525,378 | - | - | - |
| LTU | 1,341,317 | 572,024 | - | - | - |
| LVA | 1,000,736 | 448,729 | - | - | - |
| NLD | 9,535,424 | 3,730,390 | - | - | - |
| ZAF | 37,740,453 | 3,540,106 | - | - | - |
| BIH | 2,180,316 | 1,492,431 | 804,546 | 116,662 | - |
| SVN | 1,546,371 | 337,915 | - | - | - |
| SVK | 4,017,804 | 3,514,259 | 3,010,715 | 2,507,170 | 2,003,626 |
| IDN | 259,485,278 | 202,984,900 | 146,484,522 | 89,984,143 | 33,483,765 |
| CAN | 38,253,121 | 23,563,644 | 8,874,167 | - | - |
| DEU | 85,861,051 | 66,812,001 | 47,762,950 | 28,713,900 | 9,664,849 |
| PRT | 12,336,173 | 8,961,715 | 5,587,257 | 2,212,799 | - |
| RUS | 178,291,759 | 150,373,404 | 122,455,049 | 94,536,695 | 66,618,340 |

| | | | | | |
|---|---|---|---|---|---|
| CZE | 13,171,235 | 9,551,744 | 5,932,252 | 2,312,761 | - |
| SGP | 7,066,447 | 5,233,544 | 3,400,641 | 1,567,739 | - |
| MYS | 43,014,622 | 32,772,641 | 22,530,659 | 12,288,678 | 2,046,697 |
| POL | 49,217,184 | 37,595,609 | 25,974,035 | 14,352,461 | 2,730,886 |
| FIN | 7,787,289 | 3,787,857 | - | - | - |
| MEX | 180,384,112 | 149,111,738 | 117,839,365 | 86,566,992 | 55,294,618 |
| TUN | 17,959,197 | 6,730,867 | - | - | - |
| SRB | 11,000,604 | 8,694,379 | 6,388,154 | 4,081,928 | 1,775,703 |
| HUN | 15,655,957 | 13,003,042 | 10,350,127 | 7,697,212 | 5,044,297 |
| PHL | 180,581,887 | 152,937,661 | 125,293,435 | 97,649,208 | 70,004,982 |
| JOR | 18,089,850 | 14,730,095 | 11,370,341 | 8,010,586 | 4,650,831 |
| BGD | 296,360,149 | 262,531,855 | 228,703,562 | 194,875,268 | 161,046,974 |
| NOR | 9,632,215 | 6,615,378 | 3,598,542 | 581,705 | - |
| SWE | 18,619,413 | 9,923,722 | 1,228,032 | - | - |
| GRC | 19,527,349 | 16,867,815 | 14,208,282 | 11,548,749 | 8,889,215 |
| KAZ | 37,285,110 | 30,194,436 | 23,103,762 | 16,013,088 | 8,922,414 |
| GBR | 134,091,577 | 86,471,939 | 38,852,301 | - | - |
| KWT | 8,749,585 | 7,571,130 | 6,392,675 | 5,214,220 | 4,035,764 |
| TUR | 170,428,592 | 139,176,478 | 107,924,363 | 76,672,249 | 45,420,135 |
| BLR | 19,351,485 | 14,862,699 | 10,373,913 | 5,885,126 | 1,396,340 |
| HKG | 16,718,881 | 13,497,211 | 10,275,542 | 7,053,872 | 3,832,202 |
| CHE | 20,328,465 | 15,009,191 | 9,689,917 | 4,370,644 | - |
| BEL | 27,679,220 | 19,943,325 | 12,207,430 | 4,471,536 | - |
| THA | 180,480,760 | 62,206,234 | - | - | - |
| BGR | 17,665,207 | 15,134,368 | 12,603,529 | 10,072,689 | 7,541,850 |
| PAK | 587,902,034 | 497,245,256 | 406,588,477 | 315,931,699 | 225,274,920 |
| IRL | 14,387,740 | 8,686,172 | 2,984,603 | - | - |
| ESP | 165,888,684 | 117,941,702 | 69,994,720 | 22,047,739 | - |
| FRA | 239,951,328 | 96,951,899 | - | - | - |
| ROU | 72,204,399 | 58,820,760 | 45,437,121 | 32,053,482 | 18,669,843 |
| USA | 1,372,833,322 | 767,094,734 | 161,356,147 | - | - |
| LBN | 23,671,285 | 18,035,309 | 12,399,333 | 6,763,357 | 1,127,380 |
| CHN | 6,221,199,234 | 5,591,303,035 | 4,961,406,836 | 4,331,510,638 | 3,701,614,439 |

| | | | | | |
|---|---|---|---|---|---|
| VNM | 483,279,507 | 402,428,981 | 321,578,454 | 240,727,928 | 159,877,401 |
| ARE | 53,914,496 | 46,579,011 | 39,243,526 | 31,908,041 | 24,572,556 |
| EGY | 629,660,106 | 529,781,680 | 429,903,254 | 330,024,828 | 230,146,402 |
| AUT | 54,622,401 | 39,297,864 | 23,973,328 | 8,648,791 | - |
| SAU | 238,769,163 | 200,845,819 | 162,922,476 | 124,999,133 | 87,075,789 |
| ITA | 400,837,704 | 339,256,632 | 277,675,560 | 216,094,489 | 154,513,417 |
| KOR | 349,598,792 | 300,642,503 | 251,686,214 | 202,729,924 | 153,773,635 |
| PRI | 24,936,095 | 20,741,294 | 16,546,492 | 12,351,691 | 8,156,890 |
| TWN | 189,547,239 | 113,430,436 | 37,313,633 | - | - |
| HRV | 98,903,239 | 69,929,587 | 40,955,934 | 11,982,281 | - |

Notes: *Omitted countries presented (n=8) relatively higher volume-weighted prices (ex-manufacturer) for access antibiotics compared to watch, reason why net savings might not be expected. (Int$) are presented in international dollars PPP-adjusted, 2019. Blank spaces refer to those countries having either higher Access prices compared to Watch or those which respective Access % (threshold) was fulfilled already; hence no cost savings were experienced. Int$: International dollars, 2019. Table A1 indicates the abbreviations of country-ISO3-codes. Authors' analysis of IQVIA MIDAS Quarterly Sales data for the year 2019, reflecting estimates of real-world activity. Copyright IQVIA. All Rights Reserved.

**Figure A10.** Percentage of total pharmaceutical spending, including other drugs than antibiotics, that could be saved if 70% Access antibiotic consumption target is met (only among those countries with available information), data from the IQVIA MIDAS Quarterly sales data for the year 2019

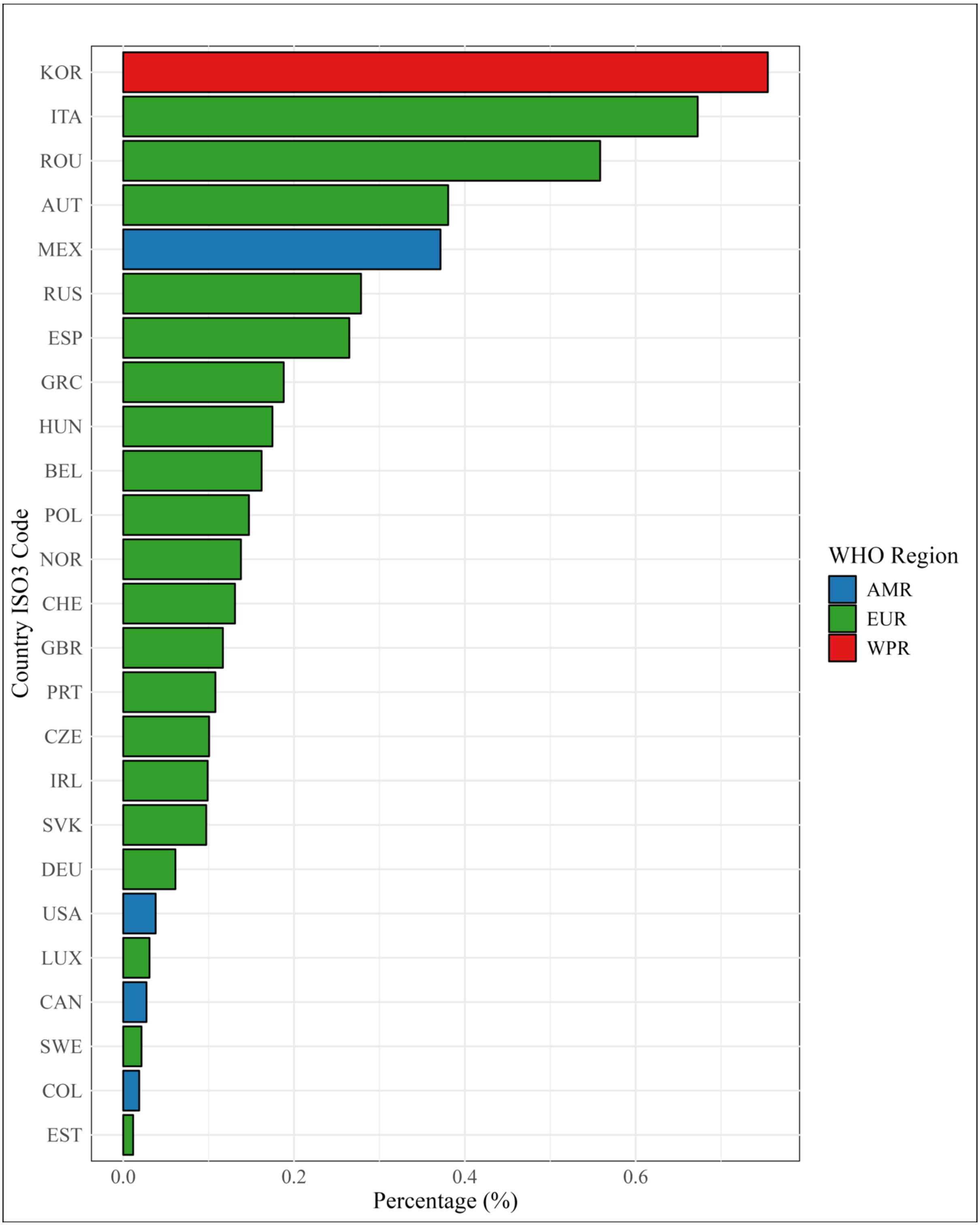

Notes: AMR= Americas, EUR= Europe, WPR= Western Pacific region. *Sample of countries is reduced due to data availability on health expenditure per country. Table A1 indicates the abbreviations of country-ISO3-codes. Authors' analysis of IQVIA MIDAS Quarterly Sales data for the year 2019, reflecting estimates of real-world activity. Copyright IQVIA. All Rights Reserved.

**Table A15.** Pharmaceutical spending per capita by country and the proportion of cost savings (if 70% access antibiotics are met) regarding the pharmaceutical spending by country*, data from the IQVIA MIDAS Quarterly sales data for the year 2019

| Country ISO3 code | Pharmaceutical spending per capita in 2019 USD[10] | Percentage of net savings if 70% Access is met, regarding pharmaceutical spending |
|---|---:|---:|
| AUT | 705.6 | 0.380 |
| BEL | 653.9 | 0.162 |
| CAN | 854.5 | 0.027 |
| CHE | 857.9 | 0.131 |
| COL | 183.0 | 0.019 |
| CZE | 550.4 | 0.101 |
| DEU | 937.4 | 0.061 |
| ESP | 557.9 | 0.265 |
| EST | 433.9 | 0.012 |
| GBR | 495.7 | 0.117 |
| GRC | 703.8 | 0.188 |
| HUN | 611.2 | 0.175 |
| IRL | 604.7 | 0.099 |
| ITA | 694.2 | 0.673 |
| KOR | 643.4 | 0.755 |
| LUX | 595.9 | 0.031 |
| MEX | 251.5 | 0.371 |
| NOR | 483.2 | 0.138 |
| POL | 466.0 | 0.147 |
| PRT | 503.6 | 0.108 |
| ROU | 422.0 | 0.558 |
| RUS | 300.4 | 0.278 |
| SVK | 557.8 | 0.097 |
| SWE | 552.8 | 0.021 |
| USA | 1280.8 | 0.038 |

Notes: USD= United States Dollars. *Sample of countries is reduced due to data availability on health expenditure per country. Table A1 indicates the abbreviations of country-ISO3-codes. Authors' analysis of IQVIA MIDAS Quarterly Sales data for the year 2019, reflecting estimates of real-world activity. Copyright IQVIA. All Rights Reserved.

**Table A16.** Volume-weighted ex-manufacturer prices per country, AWaRe category and whether antibiotics pertained to the EML book, data from the IQVIA MIDAS Quarterly sales data for the year 2019

| **Country ISO-3 code** | **Not included in EML** | | | **Included in EML** | | | **Percentage change (non EML vs EML, %)** | | |
|---|---|---|---|---|---|---|---|---|---|
| | *Access* | *Watch* | *Reserve* | *Access* | *Watch* | *Reserve* | *Access* | *Watch* | *Reserve* |
| DZA | 2.41 | 3.83 | | 1.54 | 2.61 | | 56% | 47% | |
| ARG | 5.05 | 2.18 | 171.26 | 1.66 | 2.34 | 54.06 | 204% | -7% | 217% |
| AUS | 1.05 | 1.21 | 138.85 | 0.46 | 1.78 | 37.98 | 128% | -32% | 266% |
| AUT | 12.18 | 6.28 | 144.12 | 1.69 | 10.38 | 107.93 | 622% | -39% | 34% |
| BGD | 1.27 | 1.26 | 23.87 | 0.50 | 1.44 | 5.35 | 156% | -13% | 346% |
| BLR | 2.63 | 2.89 | 227.98 | 1.44 | 2.71 | 101.94 | 82% | 6% | 124% |
| BEL | 4.01 | 2.31 | 47.70 | 0.92 | 2.55 | 68.39 | 337% | -9% | -30% |
| BIH | | 2.42 | 256.02 | 1.54 | 2.17 | 178.35 | -100% | 12% | 44% |
| BRA | 1.83 | 2.63 | 125.96 | 2.13 | 2.15 | 185.14 | -14% | 22% | -32% |
| BGR | 11.15 | 3.80 | 130.14 | 1.96 | 2.90 | 82.54 | 468% | 31% | 58% |
| CAN | 0.99 | 3.14 | 91.04 | 1.09 | 2.19 | 41.81 | -9% | 44% | 118% |
| CHL | 1.45 | 2.55 | | 1.24 | 0.91 | 194.43 | 16% | 181% | -100% |
| CHN | 4.91 | 4.85 | 78.30 | 1.51 | 5.12 | 96.45 | 225% | -5% | -19% |
| COL | 2.05 | 2.66 | 43.89 | 1.04 | 1.19 | 63.55 | 98% | 124% | -31% |
| HRV | 93.34 | 29.99 | 2854.26 | 11.31 | 30.43 | 1434.84 | 725% | -1% | 99% |
| CZE | 5.66 | 3.75 | 181.12 | 1.32 | 2.29 | 89.86 | 330% | 64% | 102% |
| DOM | 6.00 | 5.70 | 237.52 | 4.76 | 4.34 | 174.71 | 26% | 32% | 36% |
| ECU | 2.04 | 2.91 | 184.46 | 1.52 | 0.35 | 115.24 | 34% | 726% | 60% |
| EGY | 4.94 | 2.44 | 32.40 | 1.43 | 4.15 | 4.91 | 245% | -41% | 561% |
| EST | 2.96 | 1.82 | | 1.02 | 1.14 | 48.49 | 191% | 59% | -100% |
| FIN | 1.15 | 1.79 | 126.20 | 1.25 | 5.92 | 44.95 | -8% | -70% | 181% |
| FRA | 1.91 | 4.38 | 117.97 | 0.90 | 7.46 | 120.96 | 113% | -41% | -2% |
| DEU | 5.98 | 2.42 | 58.17 | 0.96 | 2.18 | 45.49 | 520% | 11% | 28% |
| GRC | 1.56 | 2.08 | 102.10 | 0.79 | 0.94 | 87.50 | 97% | 120% | 17% |
| HKG | 1.28 | 1.91 | 150.52 | 0.58 | 2.16 | 171.48 | 121% | -11% | -12% |
| HUN | 6.17 | 2.65 | 265.68 | 1.68 | 2.73 | 102.00 | 268% | -3% | 160% |
| IND | 0.55 | 0.93 | 14.02 | 1.33 | 1.22 | 5.90 | -58% | -23% | 138% |
| IDN | 2.69 | 3.25 | 119.61 | 1.17 | 5.95 | 150.61 | 130% | -45% | -21% |
| IRL | 2.43 | 2.46 | 79.97 | 0.89 | 4.24 | 58.26 | 173% | -42% | 37% |
| ITA | 3.76 | 3.04 | 134.67 | 1.28 | 4.03 | 167.21 | 194% | -24% | -19% |
| JPN | 13.28 | 2.31 | 9.23 | 1.55 | 3.09 | 103.78 | 757% | -25% | -91% |
| JOR | 4.51 | 4.26 | 214.11 | 1.83 | 3.65 | 231.14 | 146% | 17% | -7% |
| KAZ | 1.62 | 2.92 | | 1.63 | 3.01 | 7.04 | 0% | -3% | -100% |
| KOR | 4.83 | 2.78 | 59.76 | 1.01 | 4.68 | 86.07 | 378% | -41% | -31% |
| KWT | 4.16 | 4.16 | 1257.85 | 2.44 | 4.46 | | 71% | -7% | |
| LVA | 2.61 | 1.92 | 198.77 | 1.05 | 2.50 | 190.81 | 148% | -23% | 4% |
| LBN | 1.79 | 4.21 | 281.58 | 1.19 | 2.93 | 22.34 | 50% | 43% | 1160% |
| LTU | 4.02 | 2.17 | 209.83 | 1.13 | 2.12 | 52.26 | 257% | 2% | 302% |
| LUX | 2.24 | 1.36 | | 0.59 | 0.76 | | 279% | 79% | |

| | | | | | | | | | |
|---|---|---|---|---|---|---|---|---|---|
| MYS | 19.86 | 3.78 | 225.32 | 1.26 | 4.10 | 150.66 | 1470% | -8% | 50% |
| MEX | 4.06 | 5.17 | 209.66 | 3.05 | 5.49 | 136.18 | 33% | -6% | 54% |
| MAR | 2.33 | 2.75 | | 1.59 | 3.03 | 61.32 | 46% | -9% | -100% |
| NLD | 2.48 | 2.98 | 127.60 | 0.88 | 2.97 | 55.40 | 183% | 0% | 130% |
| NZL | 0.71 | 0.81 | 120.10 | 0.34 | 0.94 | 66.56 | 110% | -14% | 80% |
| NOR | 0.81 | 1.01 | 119.20 | 1.11 | 6.09 | 41.62 | -27% | -83% | 186% |
| PAK | 2.05 | 1.12 | 143.25 | 0.67 | 2.28 | 5.01 | 205% | -51% | 2758% |
| PER | 0.84 | 1.91 | 65.94 | 1.27 | 1.01 | 127.16 | -34% | 89% | -48% |
| PHL | 82.35 | 7.81 | 432.40 | 3.18 | 6.69 | 362.20 | 2486% | 17% | 19% |
| POL | 1.12 | 2.37 | 273.01 | 1.43 | 2.04 | 69.01 | -22% | 16% | 296% |
| PRT | 1.97 | 2.50 | 184.86 | 1.03 | 1.82 | 66.50 | 91% | 38% | 178% |
| PRI | 6.12 | 8.49 | 298.96 | 2.60 | 5.89 | 136.36 | 136% | 44% | 119% |
| ROU | 4.97 | 2.31 | 145.28 | 1.08 | 2.84 | 112.78 | 359% | -19% | 29% |
| RUS | 2.37 | 2.23 | 157.51 | 1.20 | 1.85 | 82.82 | 98% | 21% | 90% |
| SAU | 7.00 | 5.62 | 207.26 | 3.12 | 6.06 | 215.07 | 125% | -7% | -4% |
| SRB | 2.01 | 2.46 | 241.84 | 1.31 | 1.86 | 141.77 | 54% | 32% | 71% |
| SGP | 1.87 | 3.38 | 110.71 | 0.95 | 2.43 | 70.27 | 96% | 39% | 58% |
| SVK | 10.08 | 3.42 | 197.22 | 1.18 | 1.45 | 59.51 | 753% | 136% | 231% |
| SVN | 6.98 | 5.13 | 212.50 | 2.23 | 4.73 | 145.72 | 212% | 8% | 46% |
| ZAF | 0.65 | 5.51 | 189.62 | 0.82 | 2.15 | 21.76 | -20% | 157% | 771% |
| ESP | 1.26 | 3.64 | 119.84 | 0.61 | 2.33 | 108.14 | 106% | 56% | 11% |
| LKA | 1.73 | 0.99 | | 1.12 | 1.00 | 21.59 | 55% | -1% | -100% |
| SWE | 1.22 | 1.52 | 112.18 | 0.87 | 9.63 | 35.57 | 41% | -84% | 215% |
| CHE | 15.37 | 3.48 | 90.56 | 1.79 | 5.67 | 163.60 | 759% | -39% | -45% |
| TWN | 4.42 | 12.94 | 175.25 | 0.90 | 12.60 | 93.92 | 393% | 3% | 87% |
| THA | 0.61 | 1.70 | 363.28 | 1.07 | 10.78 | 60.29 | -43% | -84% | 503% |
| TUN | 1.10 | 3.53 | 312.86 | 1.24 | 2.39 | 170.79 | -11% | 48% | 83% |
| TUR | 3.20 | 2.25 | 27.90 | 0.90 | 1.36 | 22.58 | 257% | 65% | 24% |
| ARE | 2.03 | 5.21 | 390.29 | 2.70 | 4.32 | 292.97 | -25% | 21% | 33% |
| GBR | 1.59 | 1.60 | 76.44 | 0.98 | 4.73 | 53.49 | 63% | -66% | 43% |
| URY | 2.92 | 1.88 | 88.83 | 1.05 | 1.43 | 42.70 | 178% | 32% | 108% |
| USA | 7.98 | 9.45 | 121.53 | 0.89 | 2.58 | 43.72 | 799% | 267% | 178% |
| VEN | 0.89 | 0.63 | | 0.90 | 0.70 | 21.86 | -1% | -11% | -100% |
| VNM | 3.11 | 4.79 | 191.51 | 0.98 | 2.20 | 95.21 | 217% | 117% | 101% |

Notes: EML= Essential Medicines List. Empty cells mean no data available for the country. AWaRe= Access, Watch and Reserve. Table A1 indicates the abbreviations of country-ISO3-codes. Authors' analysis of IQVIA MIDAS Quarterly Sales data for the year 2019, reflecting estimates of real-world activity. Copyright IQVIA. All Rights Reserved.

**Figure A11.** Access-to-Watch Price Ratio among antibiotics included on the EML book only, by country, data from the IQVIA MIDAS Quarterly sales data for the year 2019

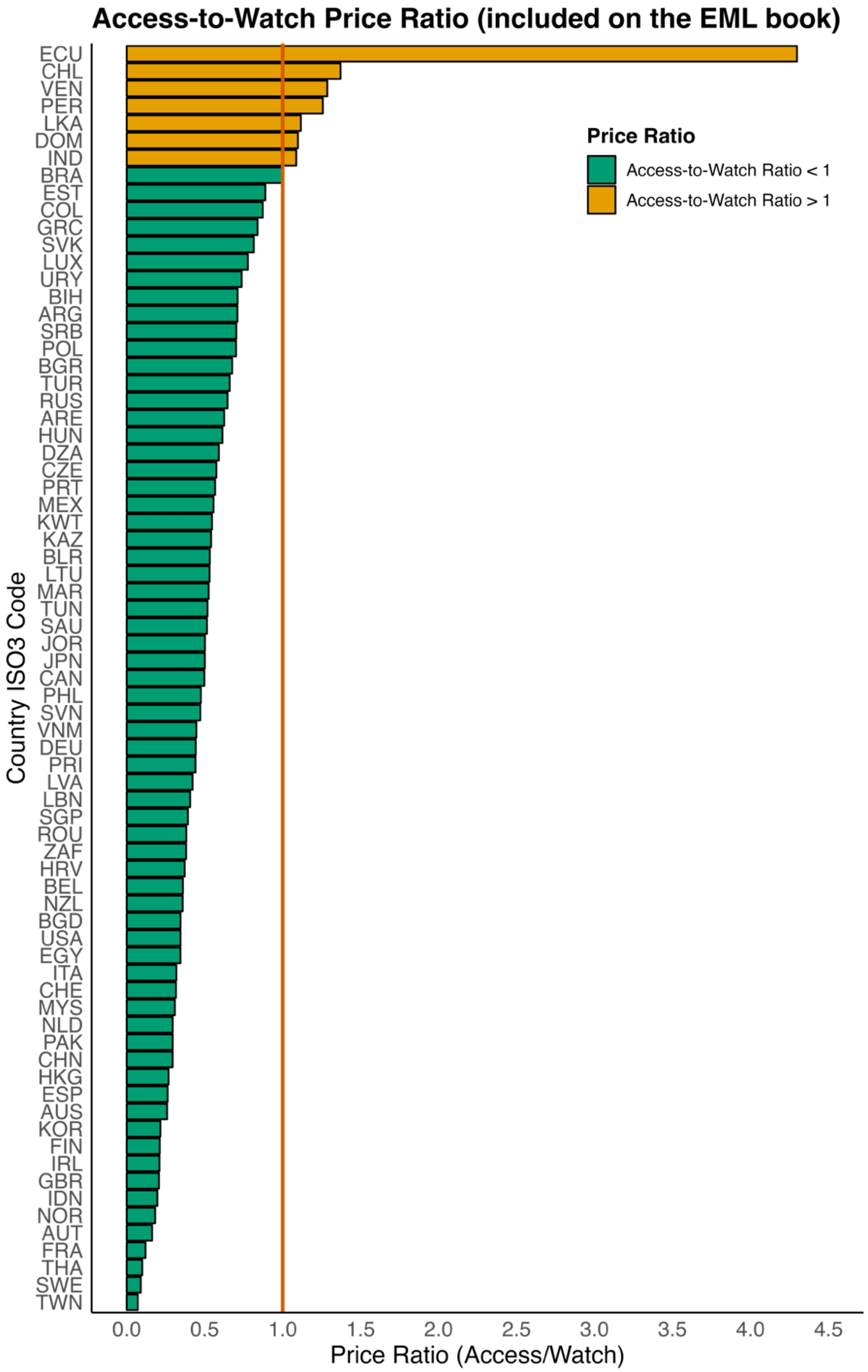

Notes: EML= Essential Medicine List. Table A1 indicates the abbreviations of country-ISO3-codes. Authors' analysis of IQVIA MIDAS Quarterly Sales data for the year 2019, reflecting estimates of real-world activity. Copyright IQVIA. All Rights Reserved.

**Table A17.** Access-to-Watch Price ratio for those antibiotics included on the EML book, by WHO-region and income level, data from the IQVIA MIDAS Quarterly sales data for the year 2019

| WHO region | Mean ratio | p25 | p75 | IQR |
|---|---|---|---|---|
| AFR | 0.499 | 0.454 | 0.559 | 0.105 |
| AMR | 1.113 | 0.556 | 1.257 | 0.701 |
| EMR | 0.463 | 0.376 | 0.536 | 0.160 |
| EUR | 0.474 | 0.300 | 0.656 | 0.356 |
| SEAR | 0.569 | 0.196 | 1.087 | 0.891 |
| WPR | 0.326 | 0.263 | 0.419 | 0.156 |
| **Income levels** | **Mean ratio** | **p25** | **p75** | **IQR** |
| HIC | 0.441 | 0.261 | 0.569 | 0.308 |
| UMC | 0.806 | 0.370 | 0.576 | 0.206 |
| LMC | 0.675 | 0.439 | 0.844 | 0.404 |

Notes: UMIC= Upper middle-income country. LMC= Lower middle income countries. HIC= High-income countries. WHO= World Health Organization. AFRO= African region, AMRO= Americas region, EMRO= East Mediterranean region, EURO= Europe, SEARO= Southeast Asia region, WPRO= Western-pacific region. p25= percentile 25$^{th}$ , p75= percentile 75$^{th}$ . IQR= Interquartile range. ABU= antibiotic usage. Authors' analysis of IQVIA MIDAS Quarterly Sales data for the year 2019, reflecting estimates of real-world activity. Copyright IQVIA. All Rights Reserved.

**Text A9.** Overall ABU shares by AWaRe groups

Access antibiotics had a median percentage of total ABU of 50% (IQR 26%) (Figure 3, panel A, and Figure Text A9.1 below), highest in Africa (74%) and lowest in Southeast Asia (36%) (Table Text A9.1, below). Europe and the EMRO presented 48% and 59% of Access antibiotics, respect to total ABU. Watch antibiotics had a median percentage of total ABU of 46% (IQR 17%), peaking in the West Pacific (54%) and lowest in Africa (23%). HICs had higher Access usage (62%) than LMCs (48%) and UMCs (48%), while Watch usage was evenly distributed. Japan, Ecuador, India, and China had the lowest Access ABU (18%, 22%, 22%, 31%), while New Zealand, Algeria, and Australia had the highest (78%, 78%, 76%).

The overall Access-to-Watch ABU ratio was 1·70, highest in Africa (mean 3·46, IQR 0·75) and Europe (mean 1·76, IQR 0·90), and lowest in Southeast Asia (mean 1·25, IQR 0·65) (Figure Text A9.2, Table Text A9.2, below). HICs had a ratio of 1·85, followed by LMCs (1·54) and UMCs (1·50).

**Figure Text A9.1.** Percentage of AWaRe antibiotics, including not recommended and unclassified, respect to total ABU and gap until 60%, 70% and 80% Access target set by the 2024 UN-GA, by country, data from the IQVIA MIDAS Quarterly sales data for the year 2019

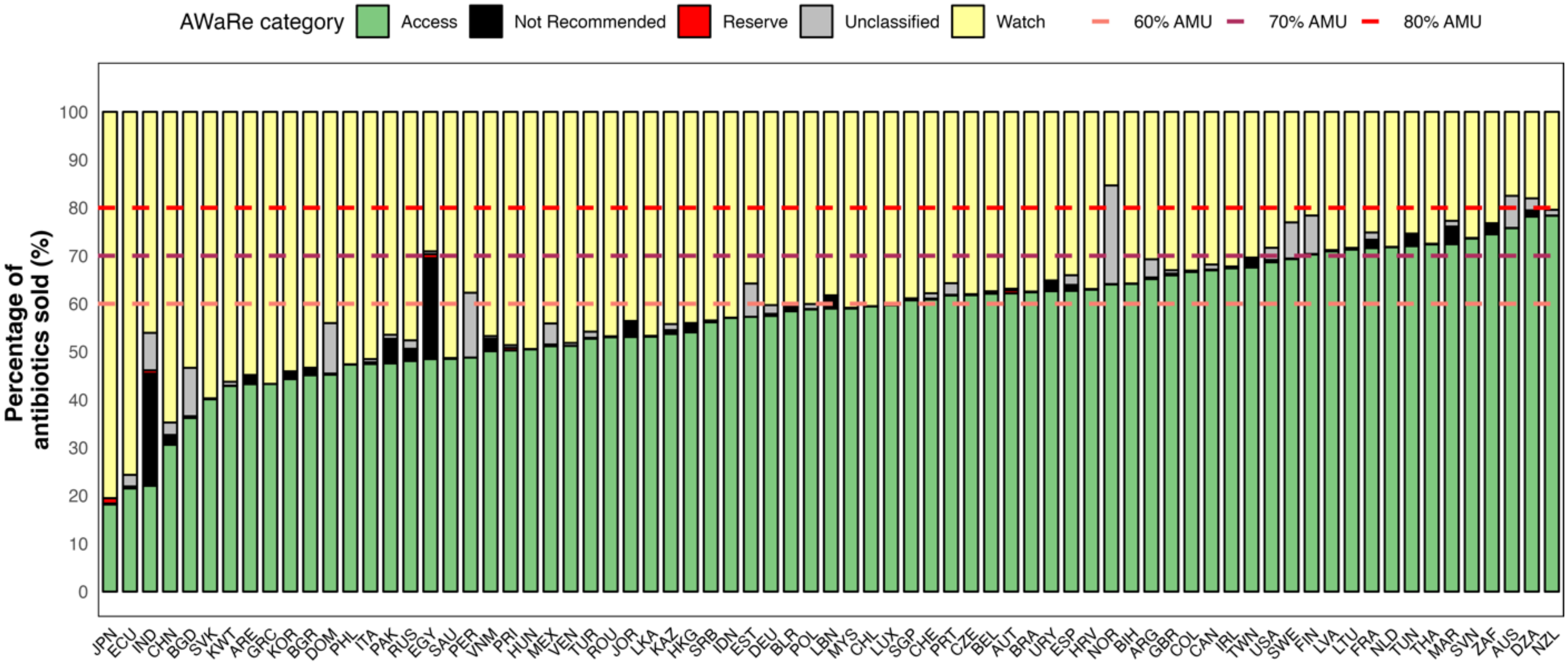

Notes: UN-GA= United Nations General Assembly zero-draft. ABU= Antibiotic usage. Table A1 indicates the abbreviations of country-ISO3-codes. Some countries reported hospital and retail data, which impacted oral and parenteral formulations and volumes; however, it was inconsistent. Authors' analysis of IQVIA MIDAS Quarterly Sales data for the year 2019, reflecting estimates of real-world activity. Copyright IQVIA. All Rights Reserved.

**Figure Text A9.2.** Percentage of Access antibiotics respect to total ABU and gap until 70% target set by the 2024 UN-GA, by country, data from the IQVIA MIDAS Quarterly sales data for the year 2019

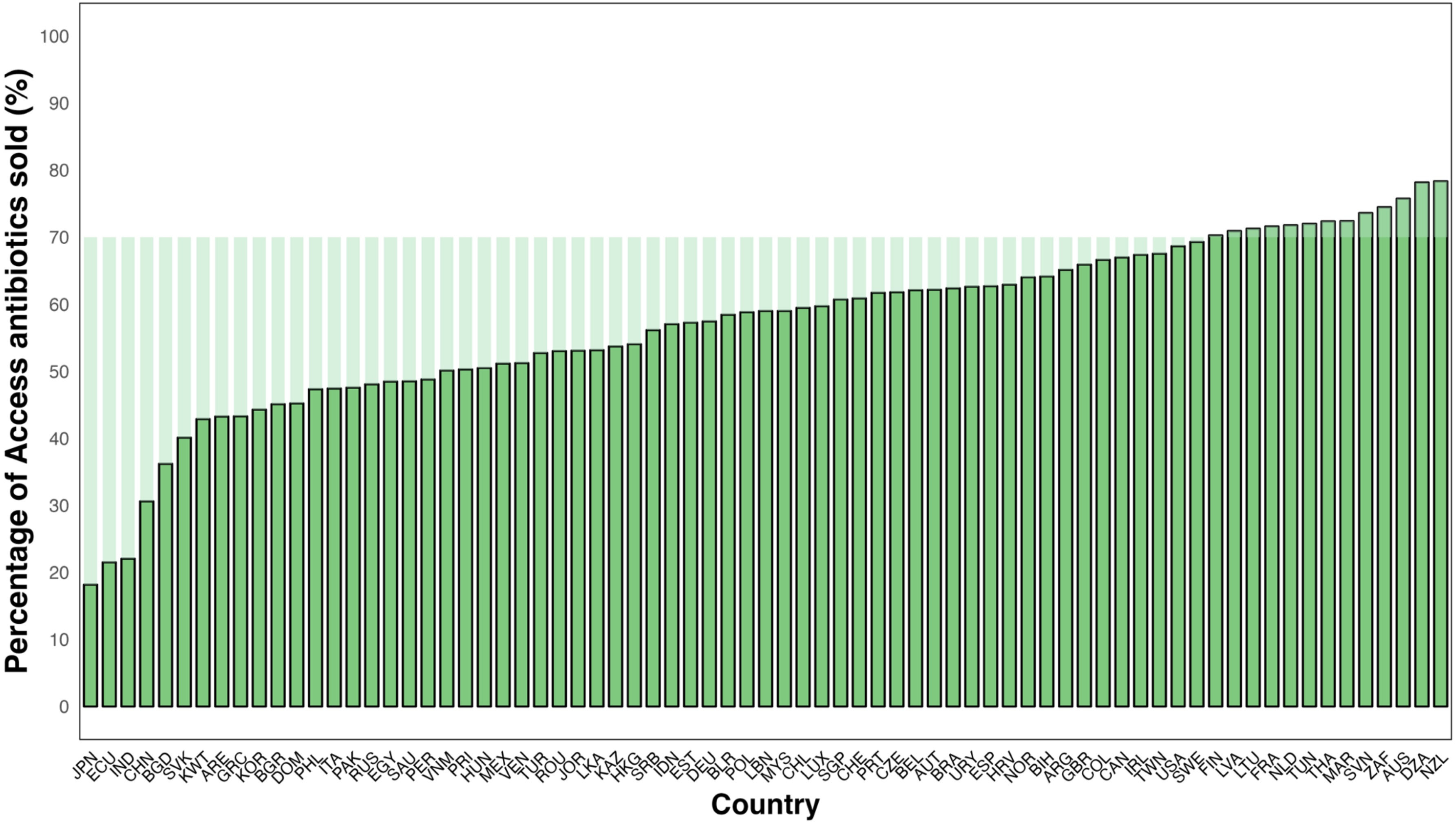

Notes: UN-GA= United Nations General Assembly zero-draft. ABU= Antibiotic usage. Table A1 indicates the abbreviations of country-ISO3-codes. Authors' analysis of IQVIA MIDAS Quarterly Sales data for the year 2019, reflecting estimates of real-world activity. Copyright IQVIA. All Rights Reserved.

**Figure Text A9.3.** Access-to-Watch ABU ratio utilising antibiotic usage in DDDs, by country, data from the IQVIA MIDAS Quarterly sales data for the year 2019

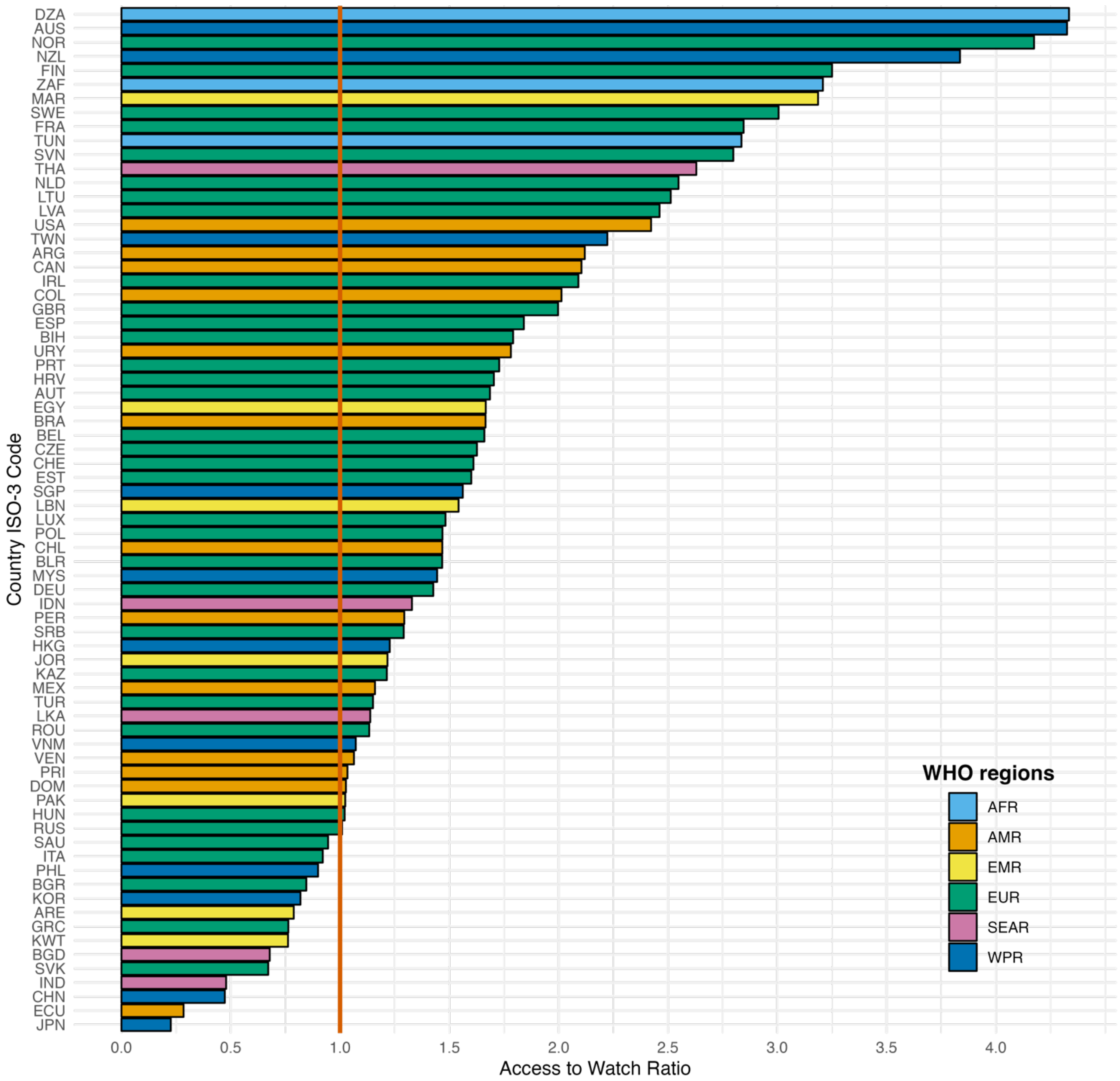

Notes: WHO= World Health Organization. AFRO= African region, AMRO= Americas region, EMRO= East Mediterranean region, EURO= Europe, SEARO= Southeast Asia region, WPRO= Western-pacific region. ABU= antibiotic usage. Table A1 indicates the abbreviations of country-ISO3-codes. Authors' analysis of IQVIA MIDAS Quarterly Sales data for the year 2019, reflecting estimates of real-world activity. Copyright IQVIA. All Rights Reserved.

**Table Text A9.1.** Percentage of access ABU across countries per WHO-region and income group (n=73 countries), data from the IQVIA MIDAS Quarterly sales data for the year 2019

| | A. Access antibiotics | | | | B. Watch antibiotics | | | | C. Reserve antibiotics | | | |
|---|---|---|---|---|---|---|---|---|---|---|---|---|
| **Group** | Median, % | p25th, % | p75th, % | IQR, % | Median, % | p25th, % | p75th, % | IQR, % | Median, % | p25th, % | p75th, % | IQR, % |
| All | 50·11 | 36·2 | 62·63 | 26·43 | 46·07 | 34·06 | 51·53 | 17·47 | 0·10 | 0·03 | 0·24 | 0·21 |
| **WHO region** | | | | | | | | | | | | |
| AFR, n=2 | 74·48 | 72·00 | 74·48 | 2·48 | 23·21 | 23·21 | 25·39 | 2·18 | 0·15 | 0·09 | 0·20 | 0·11 |
| AMR, n=13 | 62·39 | 50·28 | 68·67 | 18·39 | 37·46 | 28·34 | 44·11 | 15·77 | 0·01 | 0·00 | 0·06 | 0·06 |
| EMR, n=9 | 47·58 | 47·58 | 48·48 | 0·90 | 46·47 | 29·09 | 46·47 | 17·38 | 0·02 | 0·00 | 0·17 | 0·17 |
| EUR, n=33 | 58·83 | 50·49 | 65·92 | 15·43 | 39·83 | 33·01 | 47·63 | 14·63 | 0·14 | 0·09 | 0·21 | 0·11 |
| SEAR, n=5 | 36·20 | 22·06 | 57·04 | 34·98 | 46·07 | 42·92 | 46·07 | 3·15 | 0·08 | 0·04 | 0·33 | 0·30 |
| WPR, n=11 | 44·29 | 30·62 | 50·11 | 19·50 | 54·10 | 46·75 | 64·75 | 18·00 | 0·13 | 0·06 | 0·31 | 0·25 |
| **Income group** | | | | | | | | | | | | |
| HIC, n=40 | 62·11 | 50·28 | 68·67 | 18·39 | 36·88 | 28·34 | 48·63 | 20·29 | 0·12 | 0·06 | 0·24 | 0·18 |
| LMC, n=20 | 47·58 | 22·06 | 50·11 | 28·05 | 46·07 | 46·07 | 46·75 | 0·68 | 0·04 | 0·02 | 0·33 | 0·32 |
| UMC, n=13 | 48·07 | 30·62 | 53·71 | 23·09 | 47·63 | 38·26 | 64·75 | 26·49 | 0·04 | 0·02 | 0·15 | 0·13 |

Notes: WHO= World Health Organization. AFRO= African region, AMRO= Americas region, EMRO= East Mediterranean region, EURO= Europe, SEARO= Southeast Asia region, WPRO= Western-pacific region. HIC= High-income country, UMC= Upper middle-income country. LMC= Lower middle-income country. p25th= percentile 25th , p75th= percentile 75th. ABU= Antibiotic usage. Authors' analysis of IQVIA MIDAS Quarterly Sales data for the year 2019, reflecting estimates of real-world activity. Copyright IQVIA. All Rights Reserved.

**Table Text A9.2.** Access-to-Watch ABU ratio utilising antibiotic usage in DDDs, by WHO-region and income level, data from the IQVIA MIDAS Quarterly sales data for the year 2019

| WHO region | Mean ratio | p25 | p75 | IQR |
|---|---|---|---|---|
| AFR, n=2 | 3.460 | 3.022 | 3.772 | 0.749 |
| AMR, n=13 | 1.495 | 1.064 | 2.013 | 0.949 |
| EMR, n=9 | 1.455 | 0.906 | 1.604 | 0.698 |
| EUR, n=33 | 1.757 | 1.167 | 2.067 | 0.900 |
| SEAR, n=5 | 1.251 | 0.678 | 1.329 | 0.651 |
| WPR, n=11 | 1.646 | 0.859 | 1.892 | 1.033 |
| **Income levels** | **Mean ratio** | **p25** | **p75** | **IQR** |
| HIC, n=40 | 1.852 | 1.275 | 2.432 | 1.157 |
| LMC, n=20 | 1.495 | 1.045 | 1.902 | 0.857 |
| UMC, n=13 | 1.544 | 0.930 | 1.617 | 0.686 |

Notes: UMIC= Upper middle-income country. LMC= Lower middle-income countries. HIC= High-income countries. WHO= World Health Organization. AFRO= African region, AMRO= Americas region, EMRO= East Mediterranean region, EURO= Europe, SEARO= Southeast Asia region, WPRO= Western-pacific region. DDD= Daily defined doses. p25= percentile 25$^{th}$ , p75= percentile 75$^{th}$ . IQR= Interquartile range. ABU= antibiotic usage. Authors' analysis of IQVIA MIDAS Quarterly Sales data for the year 2019, reflecting estimates of real-world activity. Copyright IQVIA. All Rights Reserved.

**Text A10.** ABU volume and EML-listed antibiotics, by AWaRe group

Regarding ABU volume, we found that 93% of Access, 70% of Watch, and 70% of Reserve ABU pertained to the EML (Tables Text A10.1-2, Figure Text A10.1). The Southeast region had the lowest percentage of ABU for EML-listed Access antibiotics (83%), with Indonesia (74%) and Bangladesh (77%) being the lowest (Table Text A7.3, Figure Text A7.2). HICs had lower percentages of EML-listed Watch and Reserve antibiotics (69% and 63%) compared to LMCs (71% and 85%) and UMCs (72% and 76%). Access-to-Watch ABU ratio is presented in Table Text A10.4.

The median price variation (percentage change across countries) for non-EML-listed antibiotics compared to their EML-listed counterparts was 123% (IQR 216%) for Access, 0·33% (IQR 53%) for Watch, and 54% (IQR 142%) for Reserve antibiotics (Table A15), with the greatest variations in MICs (e.g., Malaysia, the Philippines, Pakistan).

**Figure Text A10.1.** Percentage of the total antibiotic usage (in DDDs) per AWaRe category and EML classification, by country, data from the IQVIA MIDAS Quarterly sales data for the year 2019

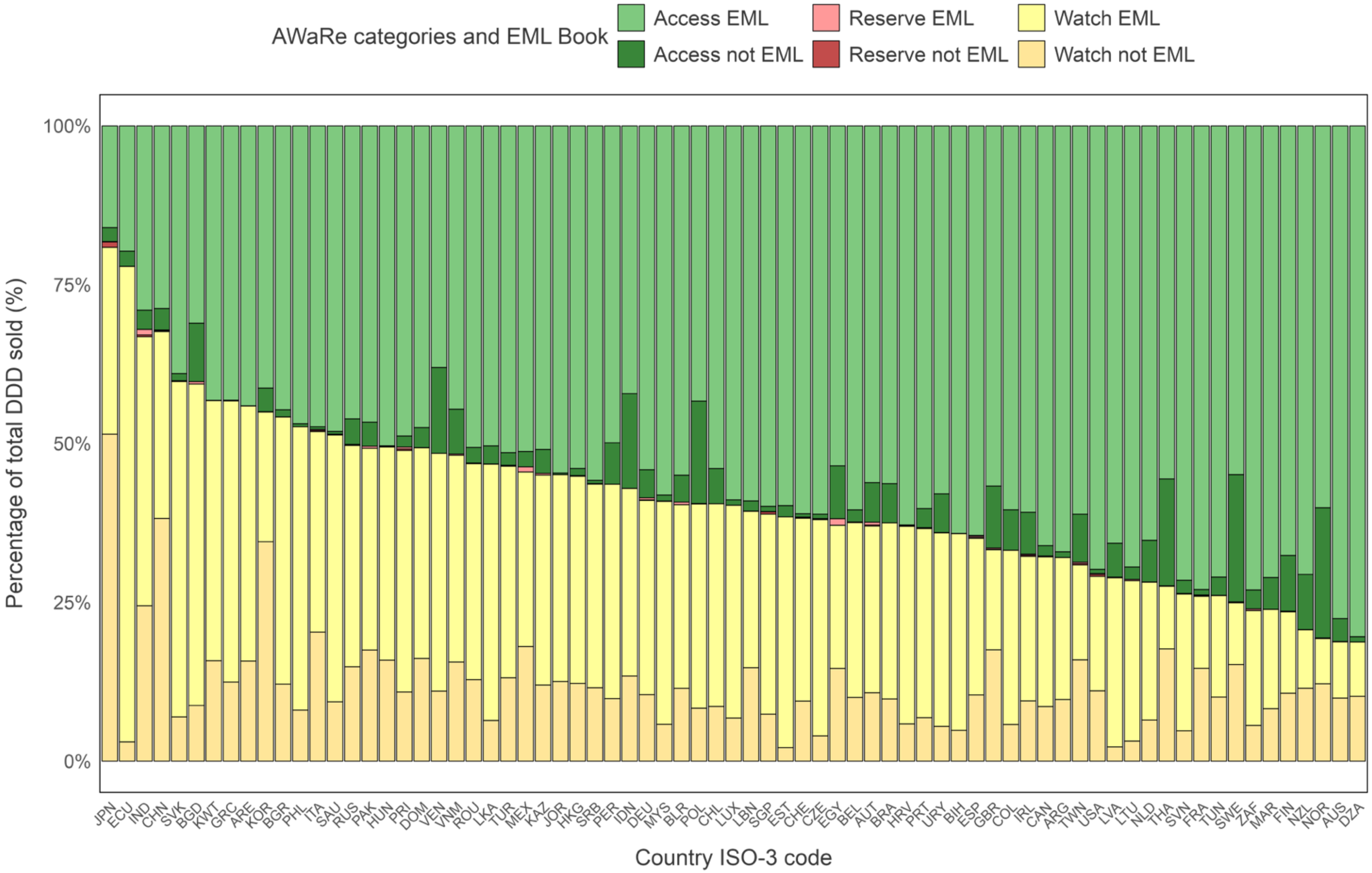

Notes: EML= Essential Medicine list. Table A1 indicates the abbreviations of country-ISO3-codes. Authors' analysis of IQVIA MIDAS Quarterly Sales data for the year 2019, reflecting estimates of real-world activity. Copyright IQVIA. All Rights Reserved.

**Table Text A10.1.** Total DDDs consumed, ABU, per country, AWaRe category and whether antibiotics pertained to the EML book, data from the IQVIA MIDAS Quarterly sales data for the year 2019

| Country ISO-3 code | Not included in EML | | | Included in EML | | |
|---|---|---|---|---|---|---|
| | *Access* | *Watch* | *Reserve* | *Access* | *Watch* | *Reserve* |
| DZA | 3,336,565 | 39,813,785 | | 313,270,605 | 33,228,871 | |
| ARG | 1,779,261 | 19,168,829 | 1,062 | 132,387,948 | 44,133,579 | 10,185 |
| AUS | 7,511,010 | 20,545,612 | 59,077 | 160,456,305 | 18,285,906 | 41,526 |
| AUT | 2,947,144 | 5,094,096 | 89,438 | 26,552,884 | 12,405,500 | 196,838 |
| BGD | 75,726,782 | 72,055,454 | 15,110 | 255,209,099 | 415,844,979 | 3,024,266 |
| BLR | 3,063,009 | 8,265,612 | 9,679 | 39,594,791 | 20,808,560 | 289,843 |
| BEL | 1,996,857 | 10,517,116 | 28,363 | 63,373,956 | 28,859,016 | 70,355 |
| BIH | | 1,010,958 | 260 | 13,372,483 | 6,452,644 | 8,565 |
| BRA | 49,221,873 | 78,073,176 | 1,611 | 448,839,685 | 220,970,763 | 1,755 |
| BGR | 614,994 | 6,568,732 | 1,530 | 24,198,290 | 22,773,465 | 14,256 |
| CAN | 3,465,400 | 18,351,623 | 222,343 | 141,224,686 | 50,406,142 | 78,446 |
| CHL | 3,868,520 | 5,999,706 | | 37,610,124 | 22,259,496 | 380 |
| CHN | 132,121,471 | 1,487,136,803 | 5,214,831 | 1,112,117,221 | 1,144,334,513 | 4,083,445 |
| COL | 10,566,288 | 9,564,831 | 1,912 | 100,190,803 | 45,458,933 | 1,025 |
| HRV | 43,126 | 1,792,691 | 1,177 | 19,282,534 | 9,551,107 | 36,267 |
| CZE | 474,543 | 2,670,413 | 31,220 | 40,951,964 | 22,803,501 | 79,269 |
| DOM | 822,193 | 4,215,040 | 3,461 | 12,391,712 | 8,653,086 | 4,836 |
| ECU | 6,054,103 | 7,596,360 | 630 | 49,447,697 | 187,719,356 | 3,469 |
| EGY | 82,624,066 | 144,732,776 | 429,518 | 530,283,977 | 223,055,194 | 9,953,938 |
| EST | 80,324 | 96,846 | | 2,706,772 | 1,644,770 | 302 |
| FIN | 2,306,063 | 2,811,063 | 12,614 | 17,766,662 | 3,363,936 | 19,026 |
| FRA | 5,062,372 | 84,285,690 | 634,110 | 420,725,474 | 65,319,984 | 587,250 |
| DEU | 18,366,010 | 43,490,455 | 409,641 | 224,882,609 | 127,034,285 | 1,295,212 |
| GRC | 156,740 | 16,555,809 | 10,663 | 57,406,225 | 58,846,182 | 1,044 |
| HKG | 468,509 | 5,164,521 | 23,985 | 22,750,049 | 13,753,864 | 32,676 |
| HUN | 69,732 | 8,352,521 | 6,898 | 26,455,696 | 17,626,380 | 21,742 |
| IND | 164,256,094 | 1,321,984,783 | 10,827,227 | 1,559,060,789 | 2,277,022,959 | 43,992,638 |
| IDN | 47,607,065 | 42,765,714 | 33,388 | 134,460,951 | 94,234,698 | 27,164 |
| IRL | 2,799,932 | 4,035,563 | 80,223 | 25,870,724 | 9,680,967 | 67,211 |
| ITA | 2,233,584 | 106,215,948 | 939,320 | 247,535,409 | 165,001,434 | 854,635 |
| JPN | 12,877,362 | 298,154,195 | 6,026,607 | 93,014,368 | 170,920,462 | 496,266 |
| JOR | 84,455 | 4,115,116 | 26 | 17,928,791 | 10,687,409 | 188 |
| KAZ | 3,886,722 | 12,364,075 | | 52,532,296 | 34,109,046 | 263,221 |
| KOR | 16,614,532 | 154,216,642 | 154,496 | 184,173,356 | 91,004,949 | 150,198 |
| KWT | 6,135 | 1,904,231 | 6 | 5,202,988 | 4,931,920 | |
| LVA | 460,098 | 193,369 | 55 | 5,658,171 | 2,292,132 | 8,488 |
| LBN | 805,266 | 7,336,770 | 1,449 | 29,423,976 | 12,265,594 | 13,828 |
| LTU | 326,365 | 526,613 | 1,690 | 11,594,692 | 4,217,657 | 31,459 |
| LUX | 38,594 | 308,231 | | 2,671,139 | 1,519,730 | |

| | | | | | | |
|---|---|---|---|---|---|---|
| MYS | 767,014 | 4,764,315 | 14,316 | 47,769,965 | 28,846,603 | 42,774 |
| MEX | 6,387,587 | 49,607,836 | 14,537 | 135,193,768 | 72,470,941 | 107,387 |
| MAR | 7,905,788 | 12,998,962 | | 111,914,530 | 24,602,739 | 4,795 |
| NLD | 3,910,305 | 3,857,230 | 2,248 | 38,830,779 | 12,915,321 | 21,656 |
| NZL | 3,712,668 | 4,906,295 | 3,293 | 30,144,676 | 3,920,241 | 15,280 |
| NOR | 5,284,507 | 3,140,314 | 4,935 | 15,488,167 | 1,835,504 | 20,430 |
| PAK | 59,455,297 | 274,560,278 | 12,442 | 731,926,740 | 498,500,666 | 5,155,827 |
| PER | 7,245,717 | 10,939,154 | 111 | 55,368,210 | 37,455,312 | 615 |
| PHL | 901,143 | 15,368,773 | 13,036 | 89,661,395 | 85,324,777 | 19,657 |
| POL | 48,807,052 | 25,228,999 | 22,978 | 131,098,218 | 97,292,072 | 223,402 |
| PRT | 2,292,271 | 5,184,359 | 24,491 | 45,587,214 | 22,523,313 | 87,113 |
| PRI | 388,533 | 2,421,995 | 52,577 | 10,859,660 | 8,458,358 | 66,911 |
| ROU | 4,595,443 | 23,999,177 | 34,217 | 94,658,980 | 63,570,952 | 176,040 |
| RUS | 31,819,910 | 116,984,398 | 178,566 | 362,888,789 | 274,155,836 | 1,033,028 |
| SAU | 1,171,759 | 25,035,836 | 125,610 | 128,865,107 | 112,526,512 | 241,312 |
| SRB | 336,096 | 7,603,159 | 15,285 | 36,662,476 | 21,051,802 | 44,930 |
| SGP | 188,247 | 1,644,406 | 79,202 | 13,302,659 | 6,995,584 | 6,903 |
| SVK | 458,626 | 2,806,640 | 5,589 | 15,697,012 | 21,258,154 | 46,708 |
| SVN | 206,477 | 473,177 | 3,578 | 7,084,066 | 2,131,594 | 6,484 |
| ZAF | 9,448,661 | 17,675,876 | 119,411 | 229,736,829 | 56,865,355 | 712,022 |
| ESP | 534,976 | 45,977,918 | 570,398 | 284,423,803 | 108,821,950 | 907,028 |
| LKA | 1,829,167 | 4,127,712 | | 32,401,619 | 25,947,633 | 22,685 |
| SWE | 8,668,551 | 6,575,212 | 30,138 | 23,727,049 | 4,201,634 | 34,494 |
| CHE | 172,320 | 3,090,535 | 51,309 | 19,945,666 | 9,403,148 | 10,403 |
| TWN | 9,847,705 | 20,782,503 | 277,065 | 79,649,389 | 19,482,123 | 286,952 |
| THA | 99,926,246 | 105,025,592 | 46,410 | 330,202,310 | 58,537,753 | 409,926 |
| TUN | 3,947,186 | 13,798,675 | 17,720 | 97,111,619 | 21,833,317 | 28,715 |
| TUR | 19,466,799 | 128,581,562 | 757,294 | 503,716,119 | 326,096,462 | 762,061 |
| ARE | 468 | 12,135,986 | 4,810 | 33,931,757 | 30,914,393 | 12,927 |
| GBR | 45,630,943 | 81,949,491 | 425,766 | 265,172,339 | 73,672,436 | 907,192 |
| URY | 1,070,639 | 964,646 | 182 | 10,201,706 | 5,362,891 | 10,590 |
| USA | 18,325,201 | 308,040,773 | 8,247,094 | 1,943,699,345 | 501,781,749 | 4,511,275 |
| VEN | 3,064,329 | 2,498,849 | | 8,619,578 | 8,485,735 | 570 |
| VNM | 62,685,215 | 138,576,704 | 14,575 | 395,127,733 | 288,534,437 | 1,389,564 |

Notes: EML= Essential Medicines List. Empty cells mean no data available for the country. AWaRe= Access, Watch and Reserve. ABU= Antibiotic usage. Table A1 indicates the abbreviations of country-ISO3-codes. Authors' analysis of IQVIA MIDAS Quarterly Sales data for the year 2019, reflecting estimates of real-world activity. Copyright IQVIA. All Rights Reserved.

**Table Text A10.2.** Percentage of antibiotics consumed by AWaRe category included and not included in the EML book, by country, data from the IQVIA MIDAS Quarterly sales data for the year 2019

| Country ISO-3 code | Not included in EML | | | Included in EML | | |
|---|---|---|---|---|---|---|
| | *Access* | *Watch* | *Reserve* | *Access* | *Watch* | *Reserve* |
| DZA | 1.05% | 54.51% | | 98.95% | 45.49% | |
| ARG | 1.33% | 30.28% | 9.44% | 98.67% | 69.72% | 90.56% |
| AUS | 4.47% | 52.91% | 58.72% | 95.53% | 47.09% | 41.28% |
| AUT | 9.99% | 29.11% | 31.24% | 90.01% | 70.89% | 68.76% |
| BGD | 22.88% | 14.77% | 0.50% | 77.12% | 85.23% | 99.50% |
| BLR | 7.18% | 28.43% | 3.23% | 92.82% | 71.57% | 96.77% |
| BEL | 3.05% | 26.71% | 28.73% | 96.95% | 73.29% | 71.27% |
| BIH | 0.00% | 13.55% | 2.95% | 100.00% | 86.45% | 97.05% |
| BRA | 9.88% | 26.11% | 47.85% | 90.12% | 73.89% | 52.15% |
| BGR | 2.48% | 22.39% | 9.69% | 97.52% | 77.61% | 90.31% |
| CAN | 2.40% | 26.69% | 73.92% | 97.60% | 73.31% | 26.08% |
| CHL | 9.33% | 21.23% | 0.00% | 90.67% | 78.77% | 100.00% |
| CHN | 10.62% | 56.51% | 56.08% | 89.38% | 43.49% | 43.92% |
| COL | 9.54% | 17.38% | 65.11% | 90.46% | 82.62% | 34.89% |
| HRV | 0.22% | 15.80% | 3.14% | 99.78% | 84.20% | 96.86% |
| CZE | 1.15% | 10.48% | 28.26% | 98.85% | 89.52% | 71.74% |
| DOM | 6.22% | 32.76% | 41.71% | 93.78% | 67.24% | 58.29% |
| ECU | 10.91% | 3.89% | 15.37% | 89.09% | 96.11% | 84.63% |
| EGY | 13.48% | 39.35% | 4.14% | 86.52% | 60.65% | 95.86% |
| EST | 2.88% | 5.56% | 0.00% | 97.12% | 94.44% | 100.00% |
| FIN | 11.49% | 45.52% | 39.87% | 88.51% | 54.48% | 60.13% |
| FRA | 1.19% | 56.34% | 51.92% | 98.81% | 43.66% | 48.08% |
| DEU | 7.55% | 25.50% | 24.03% | 92.45% | 74.50% | 75.97% |
| GRC | 0.27% | 21.96% | 91.08% | 99.73% | 78.04% | 8.92% |
| HKG | 2.02% | 27.30% | 42.33% | 97.98% | 72.70% | 57.67% |
| HUN | 0.26% | 32.15% | 24.09% | 99.74% | 67.85% | 75.91% |
| IND | 9.53% | 36.73% | 19.75% | 90.47% | 63.27% | 80.25% |
| IDN | 26.15% | 31.22% | 55.14% | 73.85% | 68.78% | 44.86% |
| IRL | 9.77% | 29.42% | 54.41% | 90.23% | 70.58% | 45.59% |
| ITA | 0.89% | 39.16% | 52.36% | 99.11% | 60.84% | 47.64% |
| JPN | 12.16% | 63.56% | 92.39% | 87.84% | 36.44% | 7.61% |
| JOR | 0.47% | 27.80% | 12.12% | 99.53% | 72.20% | 87.88% |
| KAZ | 6.89% | 26.60% | 0.00% | 93.11% | 73.40% | 100.00% |
| KOR | 8.27% | 62.89% | 50.71% | 91.73% | 37.11% | 49.29% |
| KWT | 0.12% | 27.86% | 100.00% | 99.88% | 72.14% | 0.00% |
| LVA | 7.52% | 7.78% | 0.64% | 92.48% | 92.22% | 99.36% |
| LBN | 2.66% | 37.43% | 9.48% | 97.34% | 62.57% | 90.52% |
| LTU | 2.74% | 11.10% | 5.10% | 97.26% | 88.90% | 94.90% |
| LUX | 1.42% | 16.86% | | 98.58% | 83.14% | |
| MYS | 1.58% | 14.17% | 25.08% | 98.42% | 85.83% | 74.92% |
| MEX | 4.51% | 40.64% | 11.92% | 95.49% | 59.36% | 88.08% |

| | | | | | | |
|---|---|---|---|---|---|---|
| MAR | 6.60% | 34.57% | 0.00% | 93.40% | 65.43% | 100.00% |
| NLD | 9.15% | 23.00% | 9.40% | 90.85% | 77.00% | 90.60% |
| NZL | 10.97% | 55.59% | 17.73% | 89.03% | 44.41% | 82.27% |
| NOR | 25.44% | 63.11% | 19.46% | 74.56% | 36.89% | 80.54% |
| PAK | 7.51% | 35.52% | 0.24% | 92.49% | 64.48% | 99.76% |
| PER | 11.57% | 22.60% | 15.25% | 88.43% | 77.40% | 84.75% |
| PHL | 1.00% | 15.26% | 39.87% | 99.00% | 84.74% | 60.13% |
| POL | 27.13% | 20.59% | 9.33% | 72.87% | 79.41% | 90.67% |
| PRT | 4.79% | 18.71% | 21.94% | 95.21% | 81.29% | 78.06% |
| PRI | 3.45% | 22.26% | 44.00% | 96.55% | 77.74% | 56.00% |
| ROU | 4.63% | 27.41% | 16.27% | 95.37% | 72.59% | 83.73% |
| RUS | 8.06% | 29.91% | 14.74% | 91.94% | 70.09% | 85.26% |
| SAU | 0.90% | 18.20% | 34.23% | 99.10% | 81.80% | 65.77% |
| SRB | 0.91% | 26.53% | 25.38% | 99.09% | 73.47% | 74.62% |
| SGP | 1.40% | 19.03% | 91.98% | 98.60% | 80.97% | 8.02% |
| SVK | 2.84% | 11.66% | 10.69% | 97.16% | 88.34% | 89.31% |
| SVN | 2.83% | 18.17% | 35.56% | 97.17% | 81.83% | 64.44% |
| ZAF | 3.95% | 23.71% | 14.36% | 96.05% | 76.29% | 85.64% |
| ESP | 0.19% | 29.70% | 38.61% | 99.81% | 70.30% | 61.39% |
| LKA | 5.34% | 13.72% | 0.00% | 94.66% | 86.28% | 100.00% |
| SWE | 26.76% | 61.01% | 46.63% | 73.24% | 38.99% | 53.37% |
| CHE | 0.86% | 24.74% | 83.14% | 99.14% | 75.26% | 16.86% |
| TWN | 11.00% | 51.61% | 49.12% | 89.00% | 48.39% | 50.88% |
| THA | 23.23% | 64.21% | 10.17% | 76.77% | 35.79% | 89.83% |
| TUN | 3.91% | 38.73% | 38.16% | 96.09% | 61.27% | 61.84% |
| TUR | 3.72% | 28.28% | 49.84% | 96.28% | 71.72% | 50.16% |
| ARE | 0.00% | 28.19% | 27.12% | 100.00% | 71.81% | 72.88% |
| GBR | 14.68% | 52.66% | 31.94% | 85.32% | 47.34% | 68.06% |
| URY | 9.50% | 15.25% | 1.69% | 90.50% | 84.75% | 98.31% |
| USA | 0.93% | 38.04% | 64.64% | 99.07% | 61.96% | 35.36% |
| VEN | 26.23% | 22.75% | 0.00% | 73.77% | 77.25% | 100.00% |
| VNM | 13.69% | 32.45% | 1.04% | 86.31% | 67.55% | 98.96% |

Notes: EML= Essential Medicines List. Empty cells mean no data available for the country. AWaRe= Access, Watch and Reserve. Table A1 indicates the abbreviations of country-ISO3-codes. Authors' analysis of IQVIA MIDAS Quarterly Sales data for the year 2019, reflecting estimates of real-world activity. Copyright IQVIA. All Rights Reserved.

**Table Text A10.3.** Average percentage of ABU included on the EML book across AWaRe categories, by WHO region and income level, data from the IQVIA MIDAS Quarterly sales data for the year 2019

| Group | Access | Watch | Reserve |
|---|---|---|---|
| **WHO region** | | | |
| AFR, n=2 | 97.03% | 61.02% | 73.74% |
| AMR, n=13 | 91.86% | 75.39% | 69.93% |
| EMR, n=9 | 95.59% | 67.04% | 78.13% |
| EUR, n=33 | 93.83% | 73.00% | 72.79% |
| SEAR, n=5 | 82.57% | 67.87% | 82.89% |
| WPR, n=11 | 92.98% | 58.97% | 52.27% |
| **Income level** | | | |
| HIC, n=40 | 93.47% | 68.73% | 62.68% |
| LMC, n=20 | 90.74% | 70.93% | 85.42% |
| UMC, n=13 | 93.21% | 71.59% | 75.79% |

Notes: Notes: WHO= World Health Organization. AFRO= African region, AMRO= Americas region, EMRO= East Mediterranean region, EURO= Europe, SEARO= Southeast Asia region, WPRO= Western-pacific region. HIC= High-income country, UMC= Upper middle-income country. LMC= Lower middle-income country. EML=Essential Medicine List. ABU= Antibiotic usage. n= number of countries per subgroup. Authors' analysis of IQVIA MIDAS Quarterly Sales data for the year 2019, reflecting estimates of real-world activity. Copyright IQVIA. All Rights Reserved.

**Table Text A10.4.** Access-to-Watch ratio ABU utilising antibiotic usage in DDDs for antibiotics included in the EML book and not included, by WHO-region and income level, data from the IQVIA MIDAS Quarterly sales data for the year 2019

| Included in the EML book | | | | |
|---|---|---|---|---|
| **WHO region** | **Mean ratio** | **p25** | **p75** | **IQR** |
| AFR, n=2 | 0.301 | 0.185 | 0.410 | 0.225 |
| AMR, n=13 | 0.539 | 0.160 | 0.797 | 0.637 |
| EMR, n=9 | 0.218 | 0.012 | 0.394 | 0.382 |
| EUR, n=33 | 0.472 | 0.057 | 0.609 | 0.553 |
| SEAR, n=5 | 0.737 | 0.443 | 1.051 | 0.608 |
| WPR, n=11 | 0.247 | 0.090 | 0.409 | 0.319 |
| **Income levels** | **Mean ratio** | **p25** | **p75** | **IQR** |
| HIC, n=40 | 0.471 | 0.054 | 0.710 | 0.656 |
| LMC, n=20 | 0.362 | 0.119 | 0.504 | 0.385 |
| UMC, n=13 | 0.443 | 0.147 | 0.599 | 0.452 |
| **Not in the EML book** | | | | |
| **WHO region** | **Mean ratio** | **p25** | **p75** | **IQR** |
| AFR, n=2 | 5.972 | 4.244 | 6.938 | 2.694 |
| AMR, n=13 | 1.911 | 1.432 | 2.204 | 0.772 |
| EMR, n=9 | 2.089 | 1.283 | 2.388 | 1.105 |
| EUR, n=33 | 2.459 | 1.511 | 2.658 | 1.147 |
| SEAR, n=5 | 1.923 | 0.685 | 1.427 | 0.742 |
| WPR, n=11 | 2.884 | 1.210 | 3.056 | 1.846 |
| **Income levels** | **Mean ratio** | **p25** | **p75** | **IQR** |
| HIC, n=40 | 2.805 | 1.652 | 2.853 | 1.201 |
| LMC, n=20 | 1.980 | 1.378 | 2.301 | 0.924 |
| UMC, n=13 | 2.322 | 1.100 | 2.259 | 1.158 |

Notes: UMIC= Upper middle-income country. LMIC= Lower middle-income countries. HIC= High-income countries. WHO= World Health Organization. AFRO= African region, AMRO= Americas region, EMRO= East Mediterranean region, EURO= Europe, SEARO= Southeast Asia region, WPRO= Western-pacific region. DDD= Daily defined doses. p25= percentile 25$^{th}$ , p75= percentile 75$^{th}$ . IQR= Interquartile range.
'n'= number of countries per subgroup. Authors' analysis of IQVIA MIDAS Quarterly Sales data for the year 2019, reflecting estimates of real-world activity. Copyright IQVIA. All Rights Reserved.

**Figure Text A10.2.** Access-to-Watch ratio utilising antibiotic usage in DDDs for antibiotics included in the EML book and not included, by country, data from the IQVIA MIDAS Quarterly sales data for the year 2019

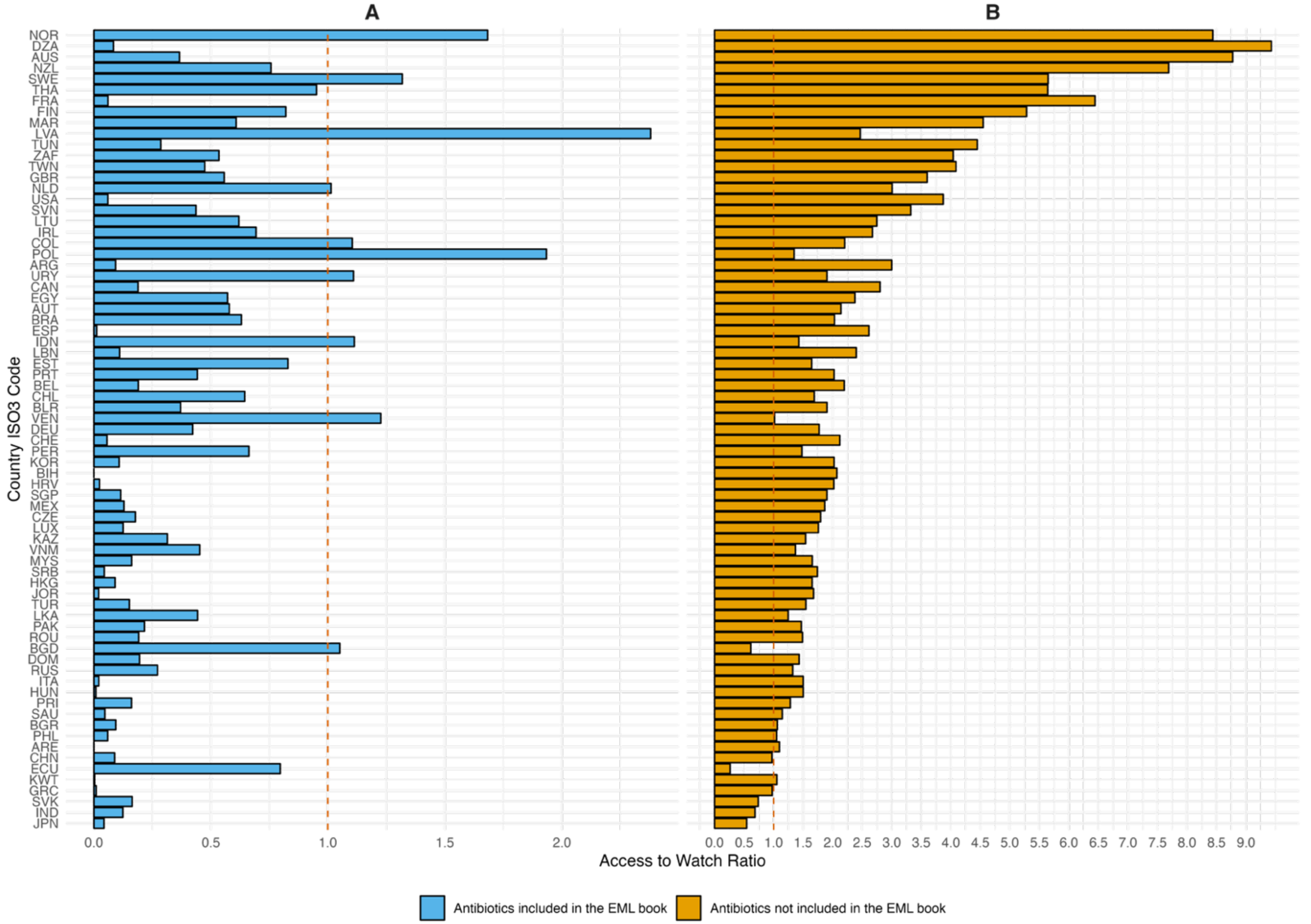

Notes: EML= Essential Medicines List. Table A1 indicates the abbreviations of country-ISO3-codes. Authors' analysis of IQVIA MIDAS Quarterly Sales data for the year 2019, reflecting estimates of real-world activity. Copyright IQVIA. All Rights Reserved.

**Table A18.** Net potential country antibiotic expenditure savings if Access consumption threshold is increased to 70% considering 2019 ABU volume by country, according to ex-manufacturer prices (n=73 countries) using EML-listed prices, data from the IQVIA MIDAS Quarterly sales data for the year 2019

| | **Ex-manufacturer prices** | |
|---|---|---|
| **Country** | **Per capita** | **Net savings** |
| Ecuador | | |
| Dominican Republic | | |
| India | | |
| Peru | | |
| Chile | | |
| Sri Lanka | | |
| Venezuela | | |
| Brazil | 0.01 | 1215240 |
| Colombia | 0.02 | 841402 |
| New Zealand | | |
| Estonia | 0.06 | 74336 |
| Algeria | | |
| Luxembourg | 0.13 | 79388 |
| Uruguay | 0.15 | 504371 |
| South Africa | | |
| Argentina | 0.15 | 6,805,667 |
| Australia | | |
| Morocco | | |
| Lithuania | | |
| Netherlands | | |
| Latvia | | |
| Bosnia | 0.23 | 767976 |
| Greece | 0.49 | 5328106 |
| Slovenia | | |
| Slovakia | 0.59 | 3251572 |
| Canada | 0.19 | 7177635 |
| Poland | 0.55 | 20847581 |
| Tunisia | | |
| Portugal | 0.49 | 5073486 |
| Czech Republic | 0.50 | 5328041 |
| Singapore | 0.54 | 3050272 |
| Russia | 0.81 | 117052621 |
| Serbia | 0.73 | 5019263 |
| Indonesia | 0.73 | 197,795,484 |
| Germany | 0.78 | 64745332 |
| Malaysia | 0.77 | 25697620 |

| | | |
|---|---|---|
| turkey | 0.95 | 78801916 |
| Jordan | 0.96 | 10451525 |
| Mexico | 1.01 | 127,224,801 |
| US | 0.19 | 64314125 |
| Hungary | 1.10 | 10760033 |
| Philippines | 1.36 | 152172995 |
| Kazakhstan | 1.26 | 23617179 |
| Kuwait | 1.53 | 6656290 |
| Belarus | 1.14 | 10711276 |
| Bangladesh | 1.74 | 290515335 |
| Finland | | |
| Hong Kong | 1.45 | 10823570 |
| Bulgaria | 1.86 | 12877519 |
| Belgium | 1.17 | 13536130 |
| Switzerland | 1.35 | 11676049 |
| Spain | 1.21 | 57057308 |
| Lebanon | 1.73 | 9806745 |
| Vietnam | 2.29 | 221652946 |
| Ireland | 0.75 | 3730754 |
| United Kingdom | 1.08 | 72126796 |
| Pakistan | 2.64 | 600,557,292 |
| Sweden | 0.28 | 2891817 |
| Japan | 3.68 | 464,962,207 |
| Romania | 2.90 | 55,922,611 |
| Norway | 1.79 | 9634805 |
| France | | |
| UAE | 3.69 | 33997065 |
| China | 4.09 | 5,777,638,284 |
| Thailand | | |
| Puerto Rico | 4.42 | 14516789 |
| Saudi Arabia | 4.70 | 169255152 |
| Italy | 5.49 | 326,328,116 |
| Taiwan | 1.61 | 37995606 |
| Austria | 3.62 | 32248950 |
| Egypt | 6.89 | 740,086,614 |
| Korea | 8.25 | 427,633,521 |
| Croatia | 10.25 | 41,498,540 |

Notes: (Int$) are presented in international dollars PPP-adjusted, 2019. Blank spaces refer to those countries having either higher Access prices compared to Watch or those which respective Access % (threshold) was fulfilled already; hence no cost savings were experienced. Int$: International dollars, 2019. Table A1 indicates the abbreviations of country-ISO3-codes. Authors' analysis of IQVIA MIDAS Quarterly Sales data for the year 2019, reflecting estimates of real-world activity. Copyright IQVIA. All Rights Reserved.

**Table A19.** Net potential country savings per capita if Access consumption threshold is increased to 60%, 65%, 70%, 75% or 80% considering 2019 ABU volume by country, according to ex-manufacturer prices (n=73 countries) using external reference pricing, data from the IQVIA MIDAS Quarterly sales data for the year 2019

| Country/ access % | Net potential antibiotic expenditure savings ex-manufacturer per capita, (Int$) | | | | |
|---|---|---|---|---|---|
| | 80% | 75% | 70% | 65% | 60% |
| IDN | 0.07 | 0.06 | 0.04 | 0.03 | 0.01 |
| THA | 0.17 | 0.06 | | | |
| LKA | 0.21 | 0.17 | 0.13 | 0.09 | 0.05 |
| VEN | 0.23 | 0.19 | 0.15 | 0.11 | 0.07 |
| NZL | 0.25 | | | | |
| DZA | 0.29 | | | | |
| NLD | 0.40 | 0.16 | | | |
| SVN | 0.43 | 0.09 | | | |
| COL | 0.44 | 0.28 | 0.11 | | |
| ZAF | 0.52 | 0.05 | | | |
| MAR | 0.59 | 0.20 | | | |
| LVA | 0.59 | 0.26 | | | |
| MEX | 0.64 | 0.53 | 0.42 | 0.31 | 0.20 |
| AUS | 0.65 | | | | |
| BGD | 0.65 | 0.57 | 0.50 | 0.43 | 0.35 |
| BRA | 0.67 | 0.48 | 0.29 | 0.10 | |
| ARG | 0.68 | 0.45 | 0.22 | | |
| SWE | 0.69 | 0.37 | 0.05 | | |
| FIN | 0.72 | 0.35 | | | |
| LTU | 0.75 | 0.32 | | | |
| CAN | 0.75 | 0.46 | 0.17 | | |
| CHL | 0.75 | 0.57 | 0.38 | 0.20 | 0.02 |
| IND | 0.88 | 0.80 | 0.72 | 0.65 | 0.57 |
| MYS | 0.92 | 0.70 | 0.48 | 0.26 | 0.04 |
| URY | 0.92 | 0.66 | 0.39 | 0.13 | |
| DOM | 0.93 | 0.80 | 0.67 | 0.53 | 0.40 |
| PHL | 0.98 | 0.83 | 0.68 | 0.53 | 0.38 |
| USA | 0.99 | 0.55 | 0.12 | | |
| CHE | 1.05 | 0.77 | 0.50 | 0.23 | |
| FRA | 1.06 | 0.43 | | | |
| EST | 1.19 | 0.93 | 0.67 | 0.41 | 0.14 |
| PER | 1.22 | 1.02 | 0.83 | 0.63 | 0.44 |
| TWN | 1.23 | 0.74 | 0.24 | | |
| SGP | 1.33 | 0.98 | 0.64 | 0.29 | |
| AUT | 1.36 | 0.98 | 0.60 | 0.22 | |

| | | | | | |
|---|---|---|---|---|---|
| JOR | 1.37 | 1.12 | 0.86 | 0.61 | 0.35 |
| NOR | 1.38 | 0.95 | 0.51 | 0.08 | |
| GBR | 1.42 | 0.91 | 0.41 | | |
| BIH | 1.42 | 0.97 | 0.53 | 0.08 | |
| IRL | 1.54 | 0.93 | 0.32 | | |
| TUN | 1.58 | 0.59 | | | |
| CZE | 1.63 | 1.18 | 0.73 | 0.29 | |
| DEU | 1.64 | 1.28 | 0.91 | 0.55 | 0.19 |
| KWT | 1.70 | 1.47 | 1.24 | 1.01 | 0.78 |
| HRV | 1.85 | 1.31 | 0.77 | 0.22 | |
| PRT | 1.97 | 1.43 | 0.89 | 0.35 | |
| PRI | 2.05 | 1.70 | 1.36 | 1.01 | 0.67 |
| LUX | 2.09 | 1.57 | 1.06 | 0.55 | 0.03 |
| KAZ | 2.11 | 1.71 | 1.31 | 0.90 | 0.50 |
| HUN | 2.27 | 1.89 | 1.50 | 1.12 | 0.73 |
| BEL | 2.33 | 1.68 | 1.03 | 0.38 | |
| ESP | 2.37 | 1.69 | 1.00 | 0.32 | |
| BLR | 2.40 | 1.84 | 1.29 | 0.73 | 0.17 |
| POL | 2.44 | 1.87 | 1.29 | 0.71 | 0.14 |
| CHN | 2.50 | 2.25 | 2.00 | 1.74 | 1.49 |
| RUS | 2.58 | 2.18 | 1.77 | 1.37 | 0.97 |
| HKG | 2.62 | 2.12 | 1.61 | 1.11 | 0.60 |
| LBN | 3.12 | 2.37 | 1.63 | 0.89 | 0.15 |
| SRB | 3.26 | 2.57 | 1.89 | 1.21 | 0.53 |
| ROU | 3.75 | 3.05 | 2.36 | 1.66 | 0.97 |
| SAU | 3.84 | 3.23 | 2.62 | 2.01 | 1.40 |
| PAK | 3.89 | 3.29 | 2.69 | 2.09 | 1.49 |
| BGR | 3.96 | 3.39 | 2.83 | 2.26 | 1.69 |
| ITA | 4.12 | 3.49 | 2.86 | 2.22 | 1.59 |
| SVK | 4.21 | 3.68 | 3.16 | 2.63 | 2.10 |
| TUR | 4.64 | 3.79 | 2.94 | 2.09 | 1.24 |
| VNM | 4.97 | 4.14 | 3.31 | 2.48 | 1.65 |
| JPN | 5.02 | 4.62 | 4.21 | 3.80 | 3.40 |
| ARE | 5.13 | 4.43 | 3.74 | 3.04 | 2.34 |
| KOR | 5.50 | 4.73 | 3.96 | 3.19 | 2.42 |
| EGY | 6.08 | 5.12 | 4.15 | 3.19 | 2.22 |
| GRC | 6.53 | 5.64 | 4.75 | 3.86 | 2.97 |
| ECU | 8.67 | 7.93 | 7.19 | 6.45 | 5.71 |

Notes: ($) are presented in international dollars PPP-adjusted, 2019. Blank spaces refer to those countries having either higher Access prices compared to Watch or those which respective Access % (threshold) was fulfilled already; hence no cost savings were experienced. Int$: International dollars, 2019. Table A1 indicates the abbreviations of country-ISO3-codes. Authors' analysis of IQVIA MIDAS Quarterly Sales data for the year 2019, reflecting estimates of real-world activity. Copyright IQVIA. All Rights Reserved.

**Table A20.** Net potential country antibiotic expenditure savings if Access consumption threshold is increased to 60%, 65%, 70%, 75% or 80% considering 2019 ABU volume by country, according to ex-manufacturer prices (n=73 countries) using external reference pricing, data from the IQVIA MIDAS Quarterly sales data for the year 2019

| Country/ access % | Net potential savings ex-manufacturer, (Int$, in millions) | | | | |
|---|---|---|---|---|---|
| | 80% | 75% | 70% | 65% | 60% |
| IDN | 19.79 | 15.48 | 11.17 | 6.86 | 2.55 |
| THA | 12.24 | 4.22 | | | |
| LKA | 4.67 | 3.80 | 2.93 | 2.06 | 1.19 |
| VEN | 6.63 | 5.47 | 4.32 | 3.17 | 2.02 |
| NZL | 1.25 | | | | |
| DZA | 12.69 | | | | |
| NLD | 6.99 | 2.74 | | | |
| SVN | 0.91 | 0.20 | | | |
| COL | 22.46 | 14.06 | 5.67 | | |
| ZAF | 30.30 | 2.84 | | | |
| MAR | 21.47 | 7.32 | | | |
| LVA | 1.12 | 0.50 | | | |
| MEX | 80.61 | 66.64 | 52.66 | 38.69 | 24.71 |
| AUS | 16.53 | | | | |
| BGD | 108.13 | 95.79 | 83.45 | 71.10 | 58.76 |
| BRA | 142.00 | 101.68 | 61.37 | 21.05 | |
| ARG | 30.91 | 20.51 | 10.11 | | |
| SWE | 7.16 | 3.81 | 0.47 | | |
| FIN | 3.98 | 1.93 | | | |
| LTU | 2.08 | 0.89 | | | |
| CAN | 28.41 | 17.50 | 6.59 | | |
| CHL | 14.46 | 10.93 | 7.41 | 3.89 | 0.37 |
| IND | 1222.09 | 1116.62 | 1011.16 | 905.70 | 800.24 |
| MYS | 30.40 | 23.16 | 15.93 | 8.69 | 1.45 |
| URY | 3.16 | 2.25 | 1.34 | 0.43 | |
| DOM | 10.26 | 8.79 | 7.31 | 5.84 | 4.36 |
| PHL | 109.97 | 93.14 | 76.30 | 59.47 | 42.63 |
| USA | 327.02 | 182.73 | 38.44 | | |
| CHE | 9.03 | 6.67 | 4.30 | 1.94 | |
| FRA | 71.34 | 28.82 | | | |
| EST | 1.58 | 1.23 | 0.89 | 0.54 | 0.19 |
| PER | 40.45 | 33.97 | 27.49 | 21.01 | 14.53 |
| TWN | 29.03 | 17.37 | 5.72 | | |
| SGP | 7.54 | 5.58 | 3.63 | 1.67 | |
| AUT | 12.09 | 8.70 | 5.31 | 1.91 | |
| JOR | 14.98 | 12.20 | 9.42 | 6.64 | 3.85 |
| NOR | 7.41 | 5.09 | 2.77 | 0.45 | |

| | | | | | |
|---|---|---|---|---|---|
| GBR | 94.93 | 61.22 | 27.50 | | |
| BIH | 4.72 | 3.23 | 1.74 | 0.25 | |
| IRL | 7.68 | 4.63 | 1.59 | | |
| TUN | 19.19 | 7.19 | | | |
| CZE | 17.44 | 12.65 | 7.85 | 3.06 | |
| DEU | 136.42 | 106.16 | 75.89 | 45.62 | 15.36 |
| KWT | 7.40 | 6.40 | 5.40 | 4.41 | 3.41 |
| HRV | 7.50 | 5.30 | 3.10 | 0.91 | |
| PRT | 20.28 | 14.73 | 9.18 | 3.64 | |
| PRI | 6.72 | 5.59 | 4.46 | 3.33 | 2.20 |
| LUX | 1.32 | 0.99 | 0.67 | 0.34 | 0.02 |
| KAZ | 39.49 | 31.98 | 24.47 | 16.96 | 9.45 |
| HUN | 22.17 | 18.41 | 14.65 | 10.90 | 7.14 |
| BEL | 26.93 | 19.40 | 11.88 | 4.35 | |
| ESP | 112.43 | 79.93 | 47.44 | 14.94 | |
| BLR | 22.50 | 17.28 | 12.06 | 6.84 | 1.62 |
| POL | 92.61 | 70.74 | 48.87 | 27.01 | 5.14 |
| CHN | 3532.04 | 3174.42 | 2816.80 | 2459.18 | 2101.56 |
| RUS | 374.94 | 316.23 | 257.52 | 198.81 | 140.09 |
| HKG | 19.62 | 15.84 | 12.06 | 8.28 | 4.50 |
| LBN | 17.65 | 13.44 | 9.24 | 5.04 | 0.84 |
| SRB | 22.47 | 17.76 | 13.05 | 8.34 | 3.63 |
| ROU | 72.20 | 58.82 | 45.44 | 32.05 | 18.67 |
| SAU | 138.37 | 116.39 | 94.41 | 72.44 | 50.46 |
| PAK | 884.55 | 748.15 | 611.75 | 475.35 | 338.95 |
| BGR | 27.46 | 23.52 | 19.59 | 15.66 | 11.72 |
| ITA | 244.96 | 207.32 | 169.69 | 132.06 | 94.42 |
| SVK | 22.98 | 20.10 | 17.22 | 14.34 | 11.46 |
| TUR | 386.85 | 315.91 | 244.97 | 174.03 | 103.10 |
| VNM | 480.55 | 400.16 | 319.76 | 239.37 | 158.97 |
| JPN | 633.92 | 582.65 | 531.39 | 480.12 | 428.85 |
| ARE | 47.28 | 40.85 | 34.42 | 27.98 | 21.55 |
| KOR | 284.86 | 244.97 | 205.08 | 165.19 | 125.30 |
| EGY | 653.57 | 549.90 | 446.23 | 342.56 | 238.89 |
| GRC | 69.81 | 60.30 | 50.79 | 41.29 | 31.78 |
| ECU | 152.50 | 139.47 | 126.43 | 113.40 | 100.36 |

Notes: (Int$) are presented in international dollars PPP-adjusted, 2019. Blank spaces refer to those countries having either higher Access prices compared to Watch or those which respective Access % (threshold) was fulfilled already; hence no cost savings were experienced. Int$: International dollars, 2019. Table A1 indicates the abbreviations of country-ISO3-codes. Authors' analysis of IQVIA MIDAS Quarterly Sales data for the year 2019, reflecting estimates of real-world activity. Copyright IQVIA. All Rights Reserved.

**Table A21.** Percentage of cost-savings relative to total pharmaceutical spending from IQVIA MIDAS Quarterly sales data for the year 2019, per country, data from the IQVIA MIDAS Quarterly sales data for the year 2019

| Country ISO-3 code | Cost-saving if 70%, total int$ | % | Cost-saving if 70% using EML-listed, total int$ | % | Cost-saving if 70% using external ref pricing, total int$ | % |
|---|---|---|---|---|---|---|
| ARE | 39243526 | 12.98 | 33997065 | 11.24 | 34420000 | 11.38 |
| ARG | 5804834 | 1.42 | 6805667 | 1.67 | 10110000 | 2.48 |
| AUS | | | | | | |
| AUT | 23973328 | 9.15 | 32248950 | 12.31 | 5310000 | 2.03 |
| BEL | 12207430 | 6.94 | 13536130 | 7.69 | 11880000 | 6.75 |
| BGD | 228703562 | 20.39 | 290515335 | 25.90 | 83450000 | 7.44 |
| BGR | 12603529 | 8.05 | 12877519 | 8.23 | 19590000 | 12.52 |
| BIH | 804546 | 1.94 | 767976 | 1.85 | 1740000 | 4.20 |
| BLR | 10373913 | 5.39 | 10711276 | 5.56 | 12060000 | 6.26 |
| BRA | 10329540 | 0.58 | 1215240 | 0.07 | 61370000 | 3.44 |
| CAN | 8874167 | 2.42 | 7177635 | 1.96 | 6590000 | 1.80 |
| CHE | 9689917 | 8.48 | 11676049 | 10.22 | 4300000 | 3.76 |
| CHL | | | | | 7410000 | 7.84 |
| CHN | 4.961E+09 | 25.77 | 5.778E+09 | 30.01 | 2.817E+09 | 14.63 |
| COL | 1738897 | 0.73 | 841402 | 0.36 | 5670000 | 2.39 |
| CZE | 5932252 | 4.36 | 5328041 | 3.91 | 7850000 | 5.76 |
| DEU | 47762950 | 5.84 | 64745332 | 7.92 | 75890000 | 9.28 |
| DOM | | | | | 7310000 | 5.13 |
| DZA | | | | | | |
| ECU | | | | | 126430000 | 59.01 |
| EGY | 429903254 | 17.05 | 740086614 | 29.35 | 446230000 | 17.70 |
| ESP | 69994720 | 8.66 | 57057308 | 7.06 | 47440000 | 5.87 |
| EST | 68141 | 1.26 | 74336 | 1.38 | 890000 | 16.51 |
| FIN | | | | | | |
| FRA | | | | | | |
| GBR | 38852301 | 4.20 | 72126796 | 7.79 | 27500000 | 2.97 |
| GRC | 14208282 | 9.95 | 5328106 | 3.73 | 50790000 | 35.57 |
| HKG | 10275542 | 15.75 | 10823570 | 16.59 | 12060000 | 18.48 |
| HRV | 40955934 | 6.46 | 41498540 | 6.54 | 3100000 | 0.49 |
| HUN | 10350127 | 7.97 | 10760033 | 8.29 | 14650000 | 11.29 |
| IDN | 146484522 | 11.82 | 197795484 | 15.97 | 11170000 | 0.90 |
| IND | | | | | 1.011E+09 | 12.11 |
| IRL | 2984603 | 3.21 | 3730754 | 4.02 | 1590000 | 1.71 |
| ITA | 277675560 | 16.74 | 326328116 | 19.68 | 169690000 | 10.23 |
| JOR | 11370341 | 11.83 | 10451525 | 10.87 | 9420000 | 9.80 |
| JPN | | | 464962207 | 28.99 | 531390000 | 33.14 |
| KAZ | 23103762 | 9.09 | 23617179 | 9.29 | 24470000 | 9.62 |
| KOR | 251686214 | 21.58 | 427633521 | 36.66 | 205080000 | 17.58 |
| KWT | 6392675 | 14.35 | 6656290 | 14.94 | 5400000 | 12.12 |
| LBN | 12399333 | 10.90 | 9806745 | 8.62 | 9240000 | 8.12 |
| LKA | | | | | 2930000 | 4.11 |
| LTU | | | | | | |
| LUX | 116748 | 3.33 | 79388 | 2.27 | 670000 | 19.13 |
| LVA | | | | | | |
| MAR | | | | | | |
| MEX | 117839365 | 9.20 | 127224801 | 9.93 | 52660000 | 4.11 |

| | | | | | | |
|---|---|---|---|---|---|---|
| MYS | 22530659 | 8.35 | 25697620 | 9.52 | 15930000 | 5.90 |
| NLD | | | | | | |
| NOR | 3598542 | 8.81 | 9634805 | 23.60 | 2770000 | 6.78 |
| NZL | | | | | | |
| PAK | 406588477 | 18.37 | 600557292 | 27.13 | 611750000 | 27.64 |
| PER | | | | | 27490000 | 17.04 |
| PHL | 125293435 | 11.39 | 152172995 | 13.84 | 76300000 | 6.94 |
| POL | 25974035 | 4.71 | 20847581 | 3.78 | 48870000 | 8.87 |
| PRI | 16546492 | 12.94 | 14516789 | 11.35 | 4460000 | 3.49 |
| PRT | 5587257 | 4.54 | 5073486 | 4.12 | 9180000 | 7.46 |
| ROU | 45437121 | 11.53 | 55922611 | 14.19 | 45440000 | 11.53 |
| RUS | 122455049 | 7.53 | 117052621 | 7.20 | 257520000 | 15.84 |
| SAU | 162922476 | 13.14 | 169255152 | 13.65 | 94410000 | 7.61 |
| SGP | 3400641 | 7.53 | 3050272 | 6.76 | 3630000 | 8.04 |
| SRB | 6388154 | 5.17 | 5019263 | 4.06 | 13050000 | 10.56 |
| SVK | 3010715 | 4.15 | 3251572 | 4.48 | 17220000 | 23.75 |
| SVN | | | | | | |
| SWE | 1228032 | 1.35 | 2891817 | 3.18 | 470000 | 0.52 |
| THA | | | | | | |
| TUN | | | | | | |
| TUR | 107924363 | 7.73 | 78801916 | 5.65 | 244970000 | 17.55 |
| TWN | 37313633 | 4.93 | 37995606 | 5.02 | 5720000 | 0.76 |
| URY | 345096 | 1.48 | 504371 | 2.16 | 1340000 | 5.73 |
| USA | 161356147 | 2.20 | 64314125 | 0.88 | 38440000 | 0.52 |
| VEN | | | | | 4320000 | 21.56 |
| VNM | 321578454 | 14.68 | 221652946 | 10.12 | 319760000 | 14.60 |
| ZAF | | | | | | |
| Mean | 161992104 | 8.62 | 196119548 | 10.22 | 129645833 | 10.67 |
| Median | 15377387 | 8.01 | 14516789 | 7.92 | 13850000 | 7.94 |
| percentile 75th | 110403114 | 11.82 | 78801916 | 13.65 | 65000000 | 14.61 |
| percentile 25th | 6274178.5 | 4.32 | 5328106 | 4.02 | 5602500 | 4.11 |
| IQR | 104128935 | 7.51 | 73473810 | 9.63 | 59397500 | 10.50 |

Notes: (Int$) are presented in international dollars PPP-adjusted, 2019. Blank spaces refer to those countries having either higher Access prices compared to Watch or those which respective Access % (threshold) was fulfilled already; hence no cost savings were experienced. Int$: International dollars, 2019. Table A1 indicates the abbreviations of country-ISO3-codes. IQR= Interquartile range. Authors' analysis of IQVIA MIDAS Quarterly Sales data for the year 2019, reflecting estimates of real-world activity. Copyright IQVIA. All Rights Reserved.

**Figure A12.** Price of treatment course (in int$) for oral Amoxicillin [Access] based on AWaRe book, by country, data from the IQVIA MIDAS Quarterly sales data for the year 2019

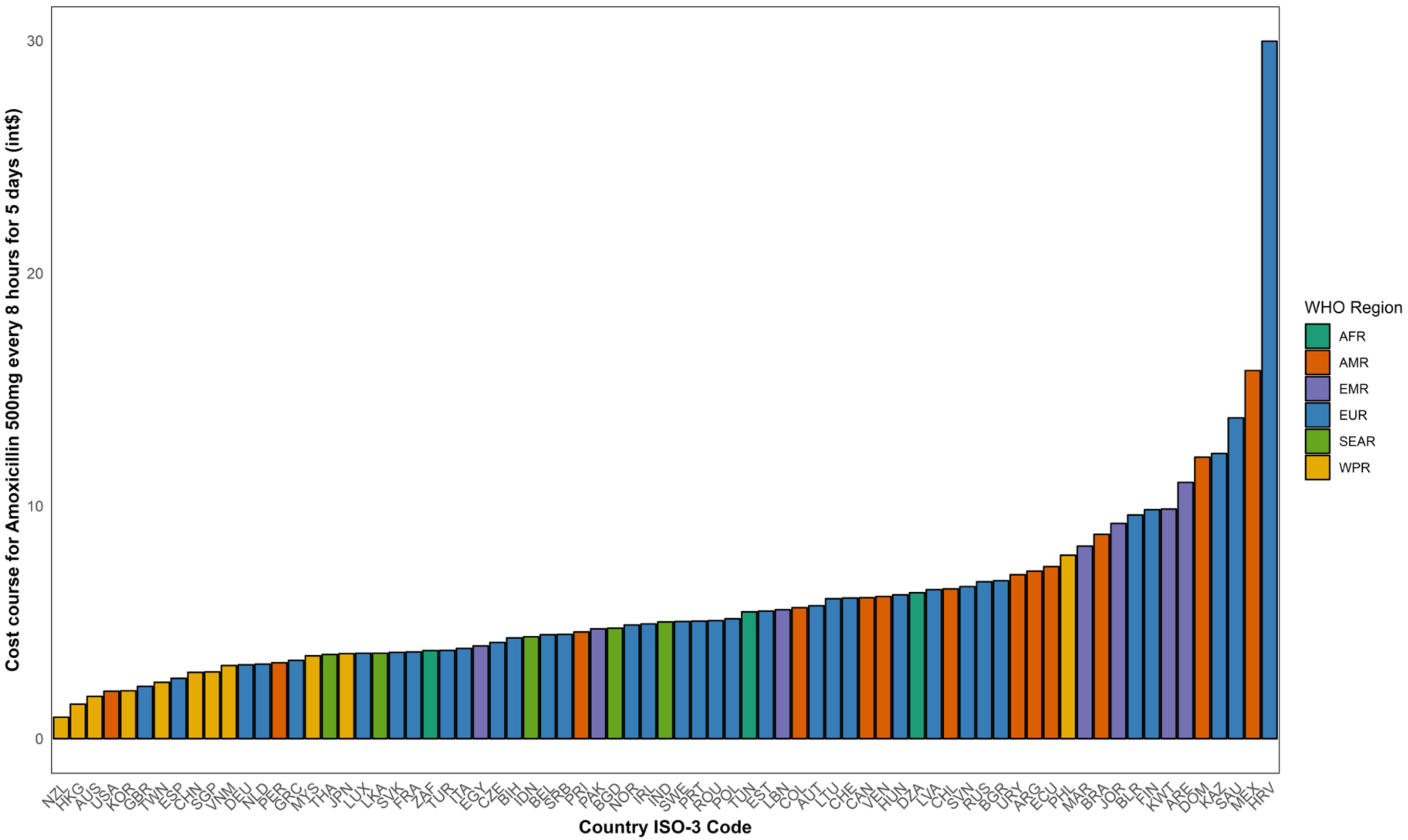

Notes: int$= International dollars. WHO= World Health Organization. AFRO= African region, AMRO= Americas region, EMRO= East Mediterranean region, EURO= Europe, SEARO= Southeast Asia region, WPRO= Western-pacific region. Table A1 indicates the abbreviations of country-ISO3-codes. Authors' analysis of IQVIA MIDAS Quarterly Sales data for the year 2019, reflecting estimates of real-world activity. Copyright IQVIA. All Rights Reserved.

**Figure A13.** Price of treatment course (in int$) for oral Amoxicillin + cluvalanic acid **[Access]** based on AWaRe book, by country, data from the IQVIA MIDAS Quarterly sales data for the year 2019

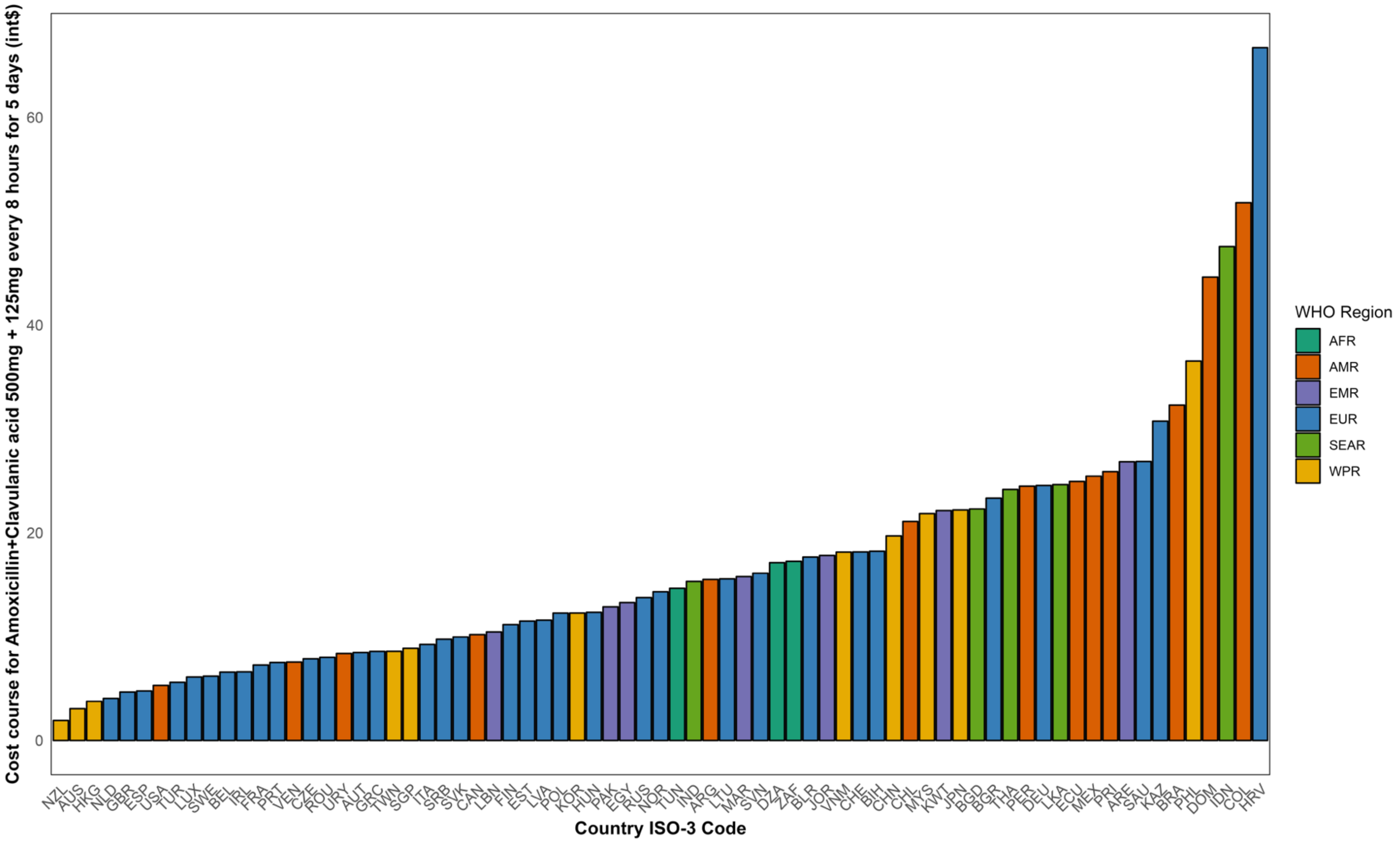

Notes: int$= International dollars. WHO= World Health Organization. AFRO= African region, AMRO= Americas region, EMRO= East Mediterranean region, EURO= Europe, SEARO= Southeast Asia region, WPRO= Western-pacific region. Table A1 indicates the abbreviations of country-ISO3-codes. Authors' analysis of IQVIA MIDAS Quarterly Sales data for the year 2019, reflecting estimates of real-world activity. Copyright IQVIA. All Rights Reserved.

**Figure A14.** Price of treatment course (in int$) for oral Cefalexin **[Access]** based on AWaRe book, by country, data from the IQVIA MIDAS Quarterly sales data for the year 2019

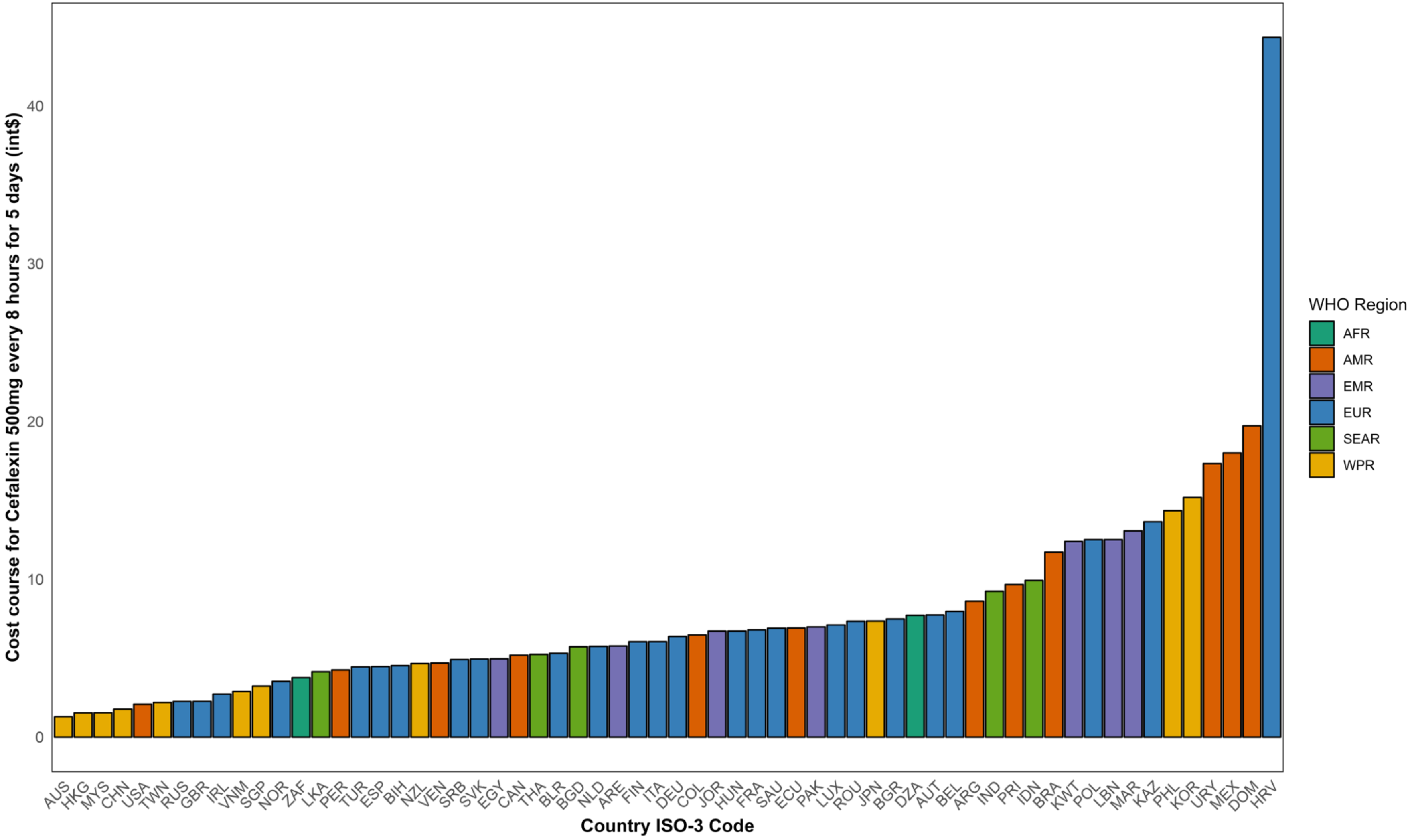

Notes: int$= International dollars. WHO= World Health Organization. AFRO= African region, AMRO= Americas region, EMRO= East Mediterranean region, EURO= Europe, SEARO= Southeast Asia region, WPRO= Western-pacific region. Table A1 indicates the abbreviations of country-ISO3-codes. Authors' analysis of IQVIA MIDAS Quarterly Sales data for the year 2019, reflecting estimates of real-world activity. Copyright IQVIA. All Rights Reserved.

**Figure A15.** Price of treatment course (in int$) for oral Doxycycline **[Access]** based on AWaRe book, by country, data from the IQVIA MIDAS Quarterly sales data for the year 2019

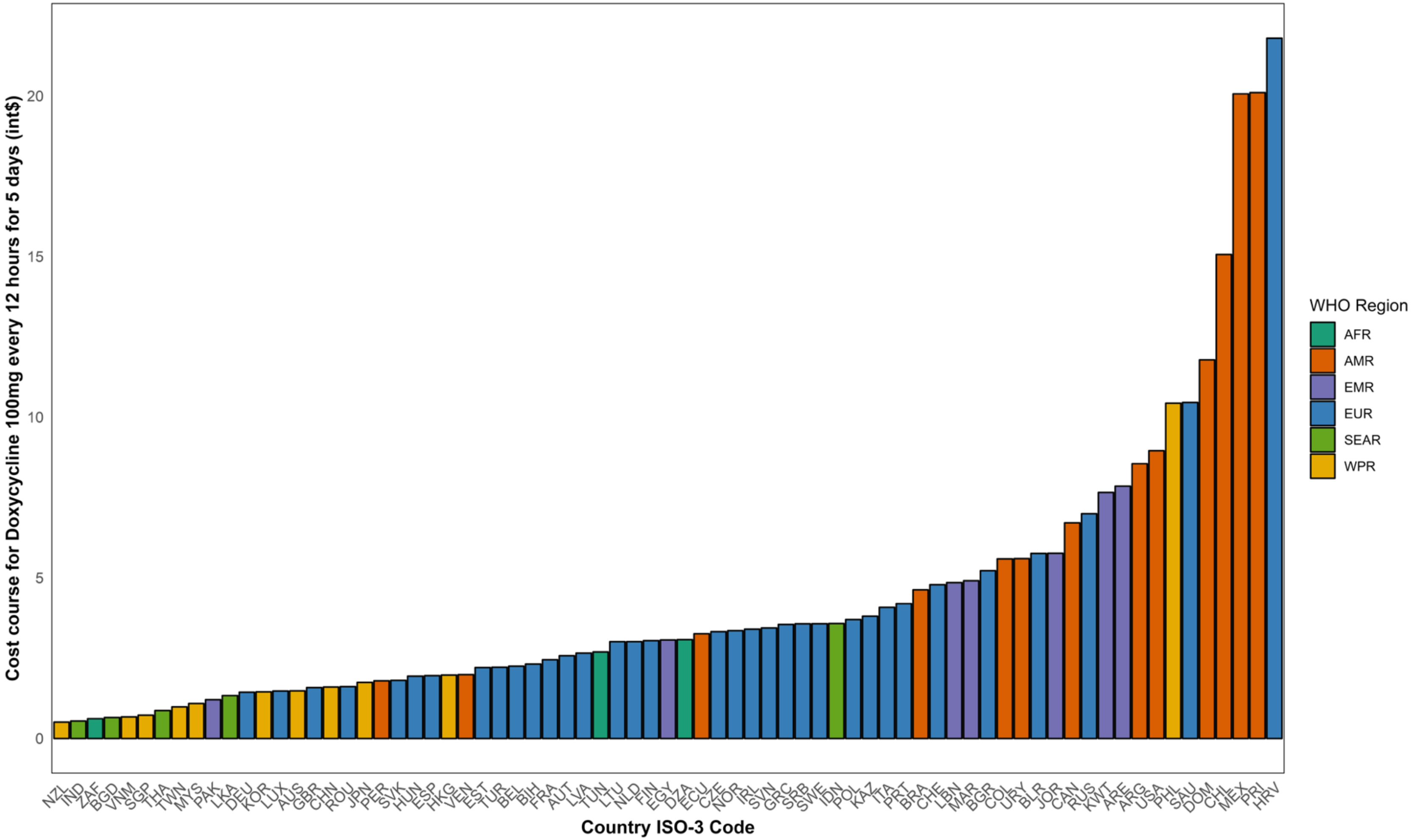

Notes: int$= International dollars. WHO= World Health Organization. AFRO= African region, AMRO= Americas region, EMRO= East Mediterranean region, EURO= Europe, SEARO= Southeast Asia region, WPRO= Western-pacific region. Table A1 indicates the abbreviations of country-ISO3-codes. Authors' analysis of IQVIA MIDAS Quarterly Sales data for the year 2019, reflecting estimates of real-world activity. Copyright IQVIA. All Rights Reserved.

**Figure A16.** Price of treatment course (in int$) for oral Nitrofurantoin **[Access]** based on AWaRe book, by country, data from the IQVIA MIDAS Quarterly sales data for the year 2019

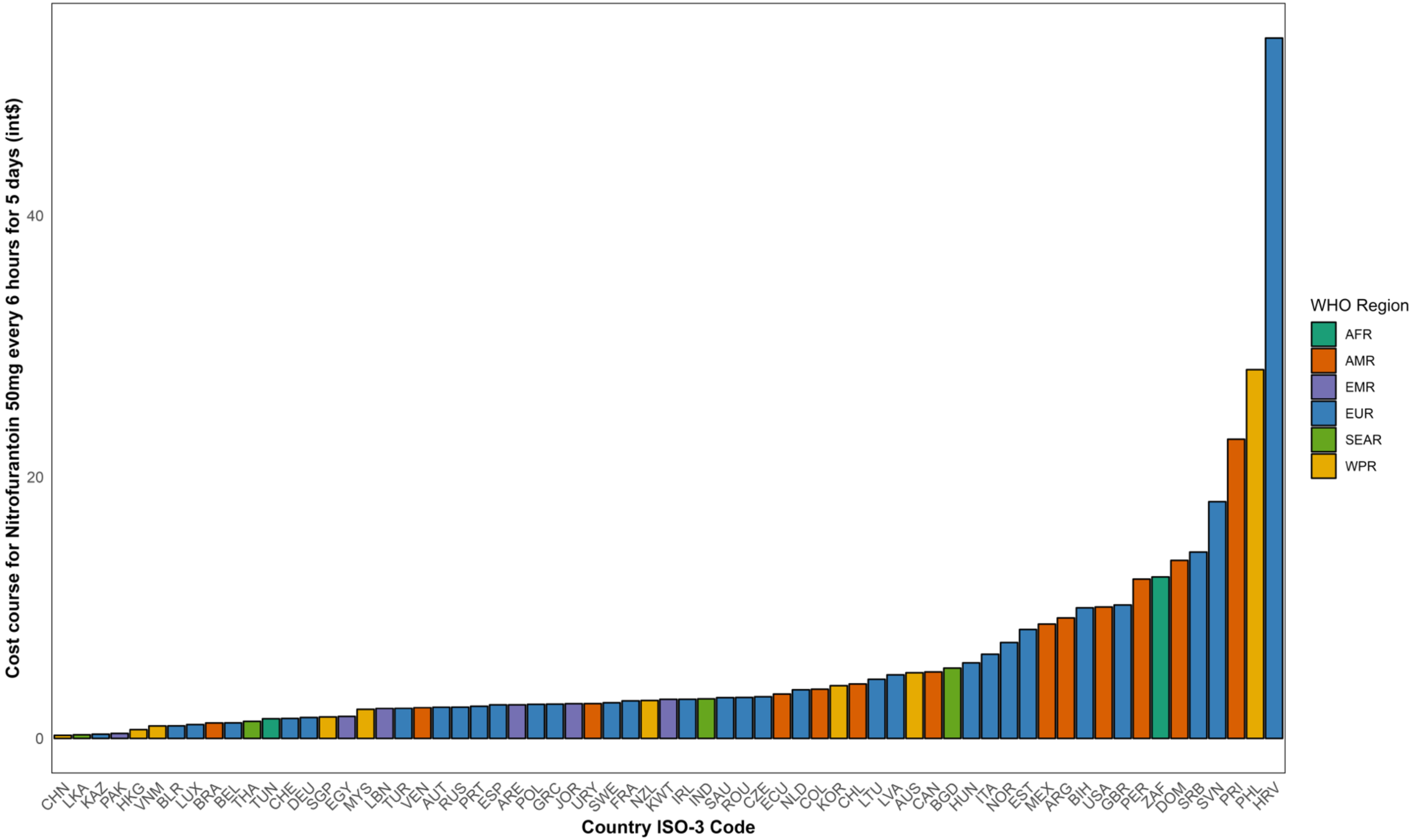

Notes: int$= International dollars. WHO= World Health Organization. AFRO= African region, AMRO= Americas region, EMRO= East Mediterranean region, EURO= Europe, SEARO= Southeast Asia region, WPRO= Western-pacific region. Table A1 indicates the abbreviations of country-ISO3-codes. Authors' analysis of IQVIA MIDAS Quarterly Sales data for the year 2019, reflecting estimates of real-world activity. Copyright IQVIA. All Rights Reserved.

**Figure A17.** Price of treatment course (in int$) for oral Phenoxymethylpenicillin **[Access]** based on AWaRe book, by country, data from the IQVIA MIDAS Quarterly sales data for the year 2019

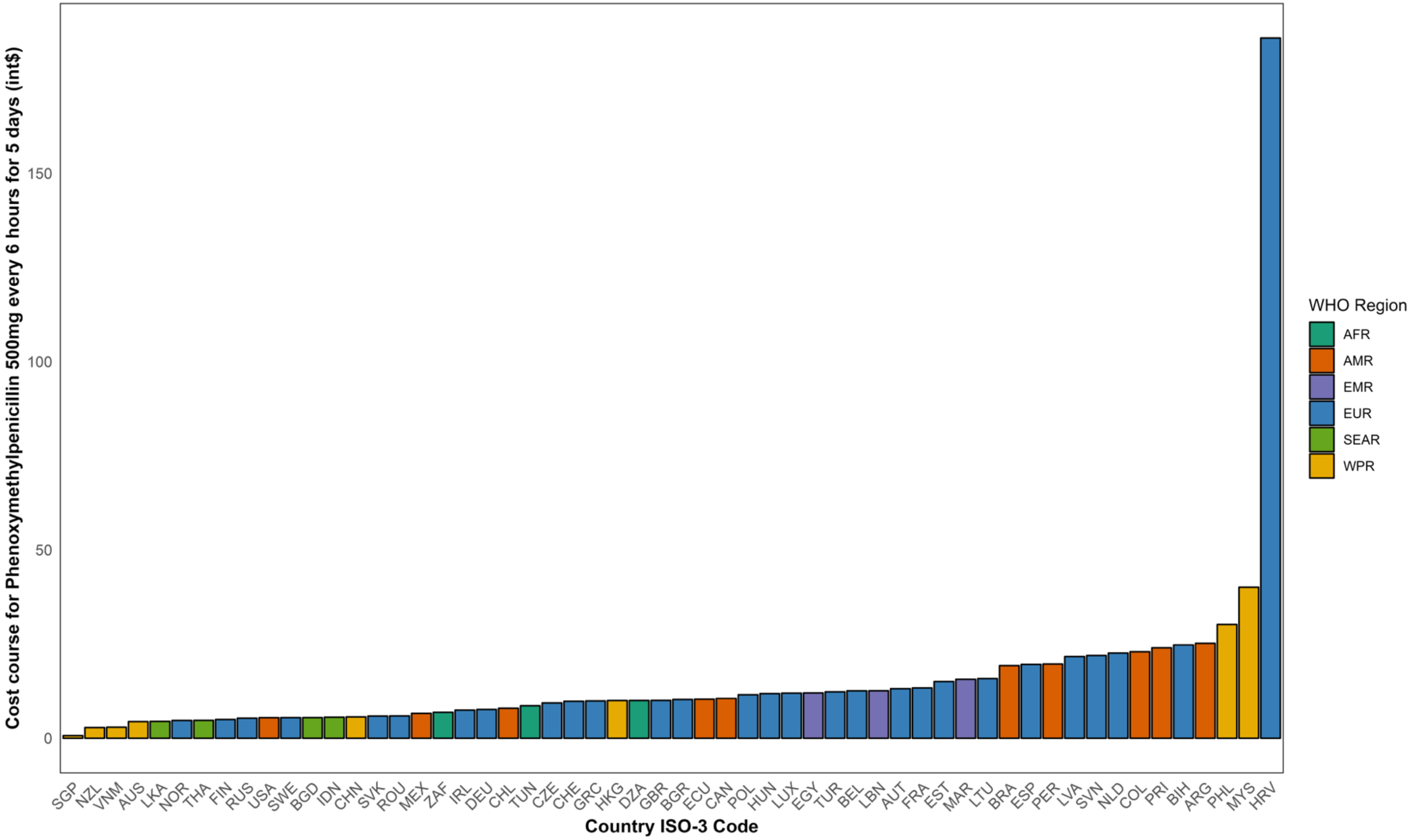

Notes: int$= International dollars. WHO= World Health Organization. AFRO= African region, AMRO= Americas region, EMRO= East Mediterranean region, EURO= Europe, SEARO= Southeast Asia region, WPRO= Western-pacific region. Table A1 indicates the abbreviations of country-ISO3-codes. Authors' analysis of IQVIA MIDAS Quarterly Sales data for the year 2019, reflecting estimates of real-world activity. Copyright IQVIA. All Rights Reserved.

**Figure A18.** Price of treatment course (in int$) for oral Azithromycin [Watch] based on AWaRe book, by country, data from the IQVIA MIDAS Quarterly sales data for the year 2019

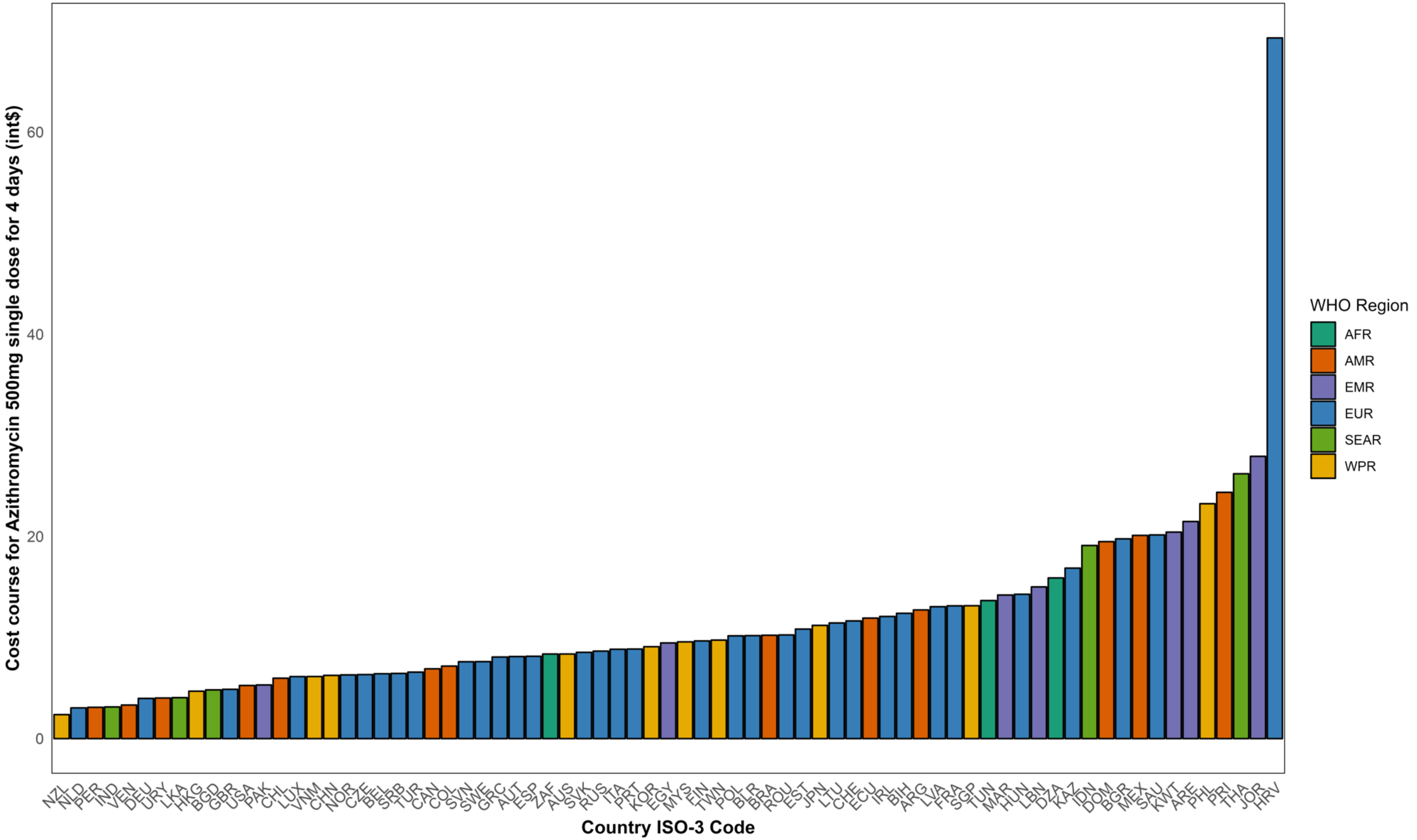

Notes: int$= International dollars. WHO= World Health Organization. AFRO= African region, AMRO= Americas region, EMRO= East Mediterranean region, EURO= Europe, SEARO= Southeast Asia region, WPRO= Western-pacific region. Table A1 indicates the abbreviations of country-ISO3-codes. Authors' analysis of IQVIA MIDAS Quarterly Sales data for the year 2019, reflecting estimates of real-world activity. Copyright IQVIA. All Rights Reserved.

**Figure A19.** Price of treatment course (in int$) for oral Cefixime [Watch] based on AWaRe book, by country, data from the IQVIA MIDAS Quarterly sales data for the year 2019

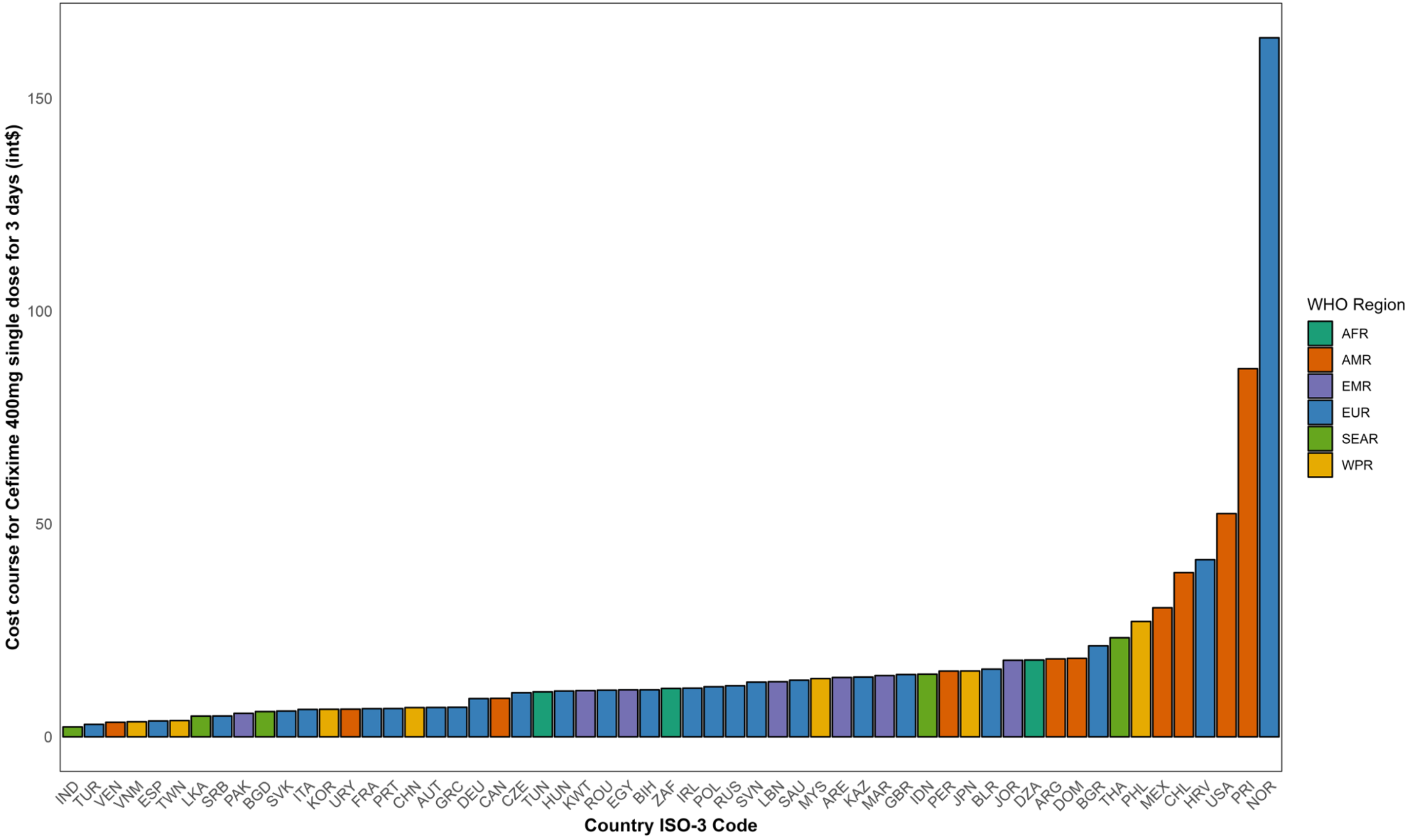

Notes: int$= International dollars. WHO= World Health Organization. AFRO= African region, AMRO= Americas region, EMRO= East Mediterranean region, EURO= Europe, SEARO= Southeast Asia region, WPRO= Western-pacific region. Table A1 indicates the abbreviations of country-ISO3-codes. Authors' analysis of IQVIA MIDAS Quarterly Sales data for the year 2019, reflecting estimates of real-world activity. Copyright IQVIA. All Rights Reserved.

**Figure A20.** Price of treatment course (in int$) for oral Ciprofloxacin [Watch] based on AWaRe book, by country, data from the IQVIA MIDAS Quarterly sales data for the year 2019

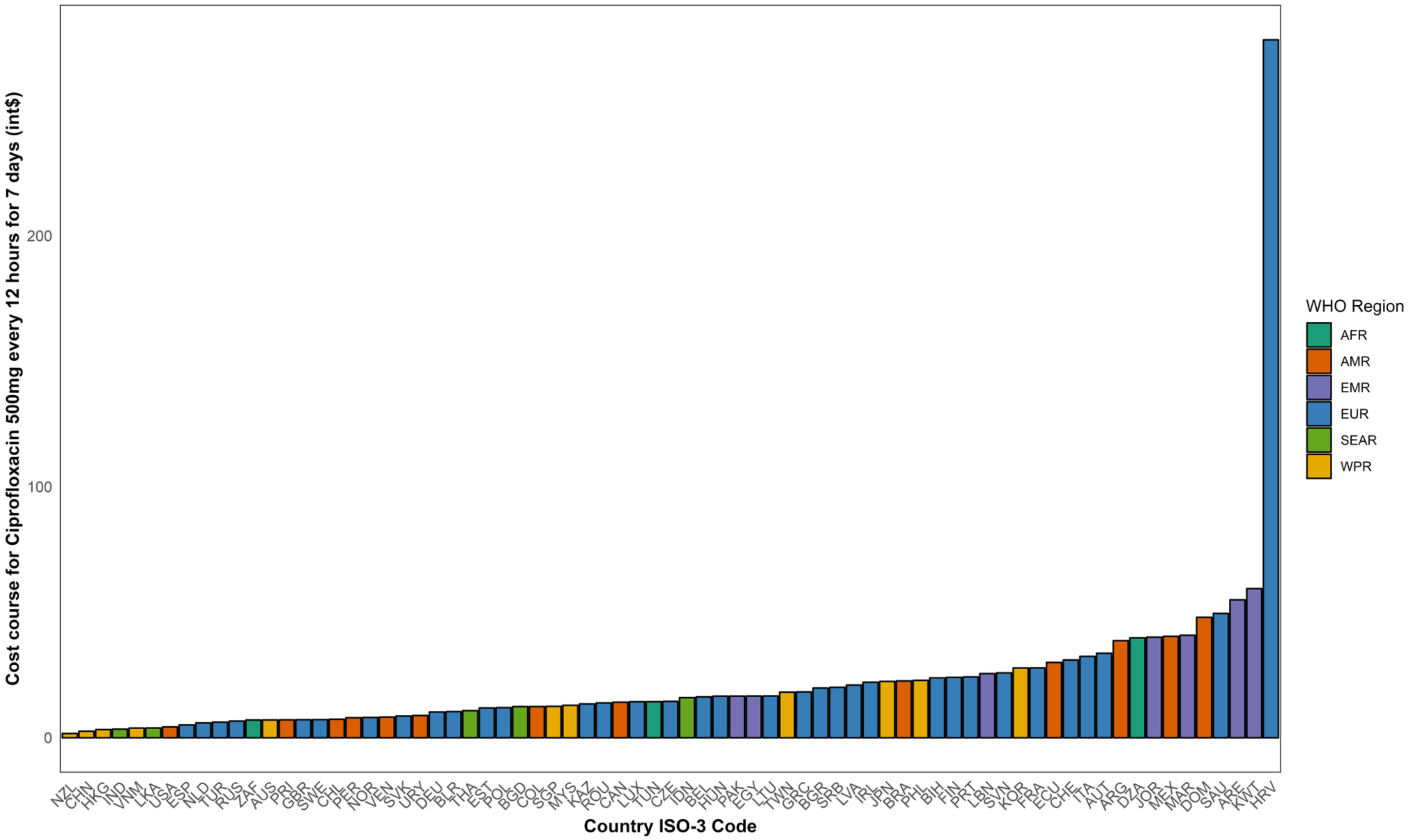

Notes: int$= International dollars. WHO= World Health Organization. AFRO= African region, AMRO= Americas region, EMRO= East Mediterranean region, EURO= Europe, SEARO= Southeast Asia region, WPRO= Western-pacific region. Table A1 indicates the abbreviations of country-ISO3-codes. Authors' analysis of IQVIA MIDAS Quarterly Sales data for the year 2019, reflecting estimates of real-world activity. Copyright IQVIA. All Rights Reserved.

**Figure A21.** Price of treatment course (in int$) for oral Clindamycin [Watch] based on AWaRe book, by country, data from the IQVIA MIDAS Quarterly sales data for the year 2019

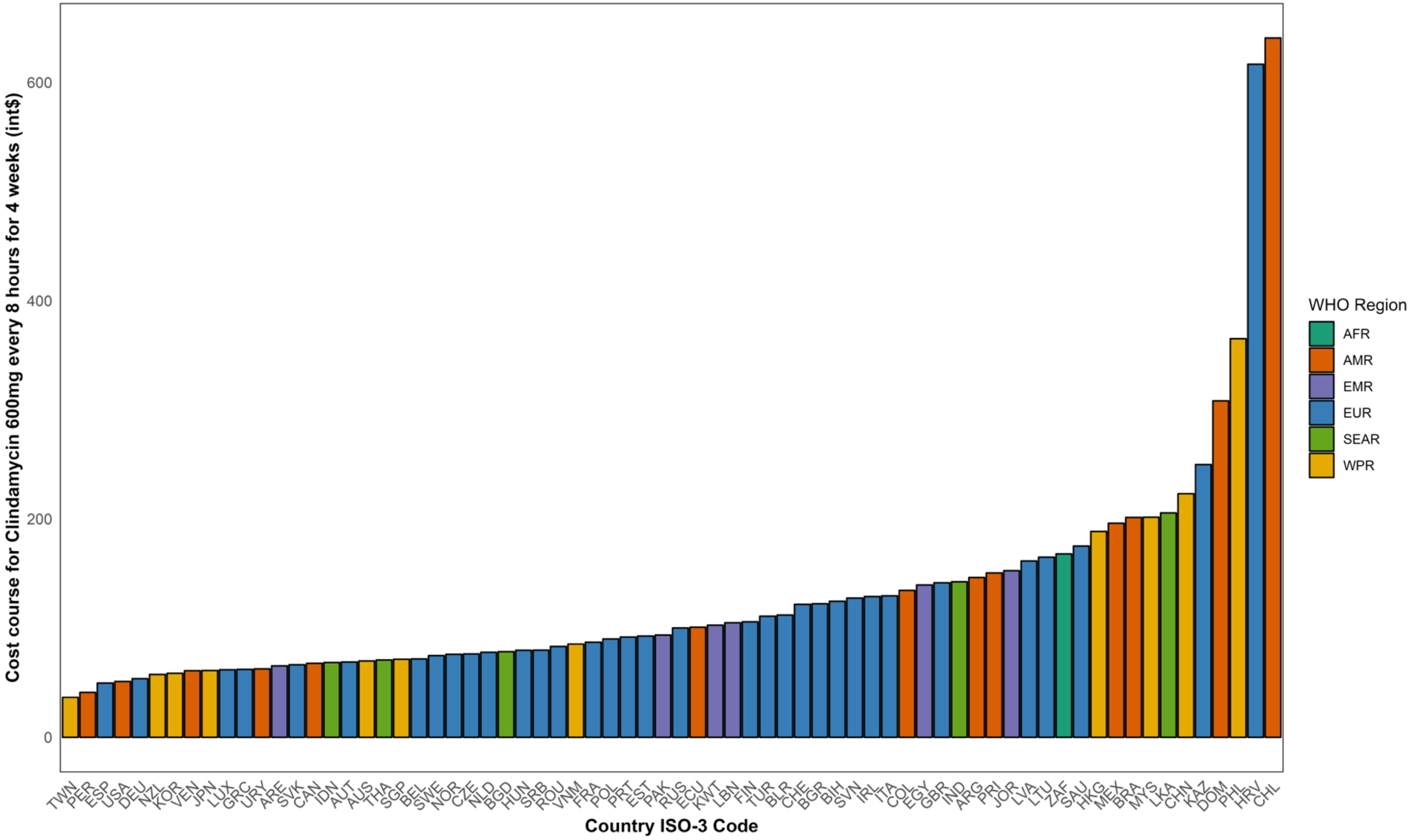

Notes: int$= International dollars. WHO= World Health Organization. AFRO= African region, AMRO= Americas region, EMRO= East Mediterranean region, EURO= Europe, SEARO= Southeast Asia region, WPRO= Western-pacific region. Table A1 indicates the abbreviations of country-ISO3-codes. Authors' analysis of IQVIA MIDAS Quarterly Sales data for the year 2019, reflecting estimates of real-world activity. Copyright IQVIA. All Rights Reserved.

**Figure A22.** Price of treatment course (in int$) for oral Linezolid **[Reserve]** based on AWaRe book, by country, data from the IQVIA MIDAS Quarterly sales data for the year 2019

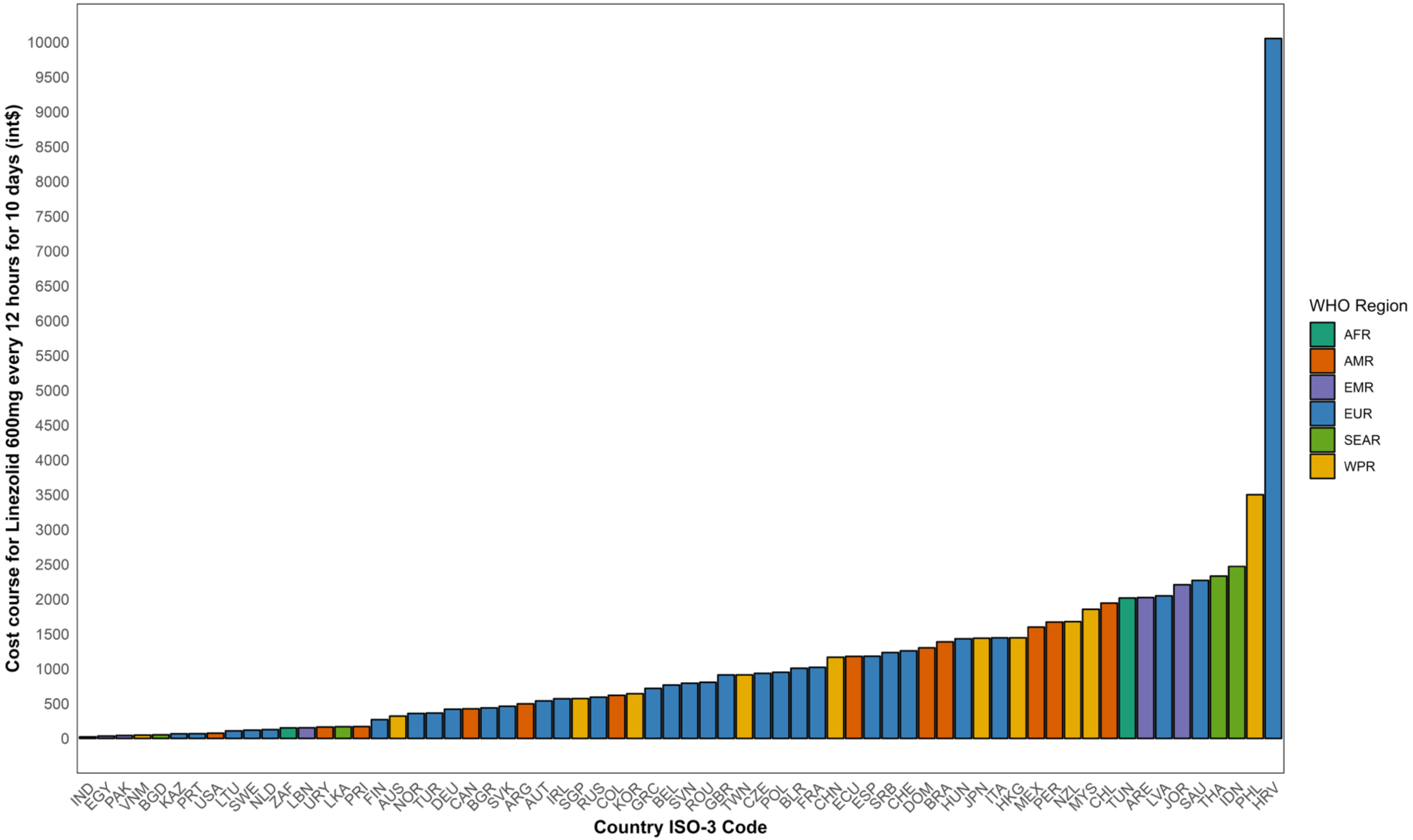

Notes: int$= International dollars. WHO= World Health Organization. AFRO= African region, AMRO= Americas region, EMRO= East Mediterranean region, EURO= Europe, SEARO= Southeast Asia region, WPRO= Western-pacific region. Table A1 indicates the abbreviations of country-ISO3-codes. Authors' analysis of IQVIA MIDAS Quarterly Sales data for the year 2019, reflecting estimates of real-world activity. Copyright IQVIA. All Rights Reserved.

**Table A22.** Pricing treatment courses for most common infections and most reported oral antibiotics across countries, following the AWaRe book (int$), data from the IQVIA MIDAS Quarterly sales data for the year 2019

| Antibiotic treatment course | AWaRe category | Treatment | Indications | Median price | p25th | p75th | IQR |
|---|---|---|---|---|---|---|---|
| Amoxicillin | Access | 500mg, every 8 hours for 5 days | Pharyngitis, Acute otitis media, COPD exacerbation (mild) | 5.02 | 3.68 | 6.54 | 2.86 |
| Amoxicillin + Clavulanic acid | Access | 500mg + 125mg, every 8 hours for 5 days | Acute otitis media, Acute sinusitis | 14.32 | 8.47 | 22.19 | 13.72 |
| Cefalexin | Access | 500mg, every 8 hours for 5 days | Respiratory infections | 6.22 | 4.46 | 8.45 | 3.99 |
| Doxycycline | Access | 500mg, every 12 hours for 5 days | CAP (mild), COPD exacerbation (mild) | 3.07 | 1.75 | 4.91 | 3.16 |
| Nitrofurantoin | Access | 50mg every 6 hours for 5 days | Lower UTIs | 3.08 | 2.22 | 5.78 | 3.56 |
| Phenoxymethylpenicillin | Access | 500mg, every 6 hours for 5 days | Respiratory infections | 10.16 | 5.84 | 15.70 | 9.86 |
| Azithromycin | Watch | 500mg, single dose for 4 days | Enteric fever(mild), Infectious acute diarrhoea | 9.46 | 6.33 | 13.15 | 6.81 |
| Cefixime | Watch | 400mg single dose for 3 days | Bacterial Infections | 11.37 | 6.61 | 15.45 | 8.85 |
| Ciprofloxacin | Watch | 500mg, every 12 hours for 7 days | Upper UTI (mild), Enteric fever (mild), Febrile neutropenia (low risk) | 14.50 | 8.08 | 24.26 | 16.17 |
| Clindamycin | Watch | 600mg every 8 hours for 4 weeks | Bacterial Infections | 96.95 | 70.18 | 145.46 | 75.28 |
| Linezolid | Reserve | 600mg every 12 hours for 10 days | Mild-severe bacterial infections | 780.47 | 284.28 | 1439.18 | 1154.90 |

Notes: UTI= Urinary tract infections. IQR= Interquartile range. P25th= 25th percentile. Mg= milligrams. COPD= Chronic obstructive pulmonary disease. CAP= Community-acquired pneumonia. Authors' analysis of IQVIA MIDAS Quarterly Sales data for the year 2019, reflecting estimates of real-world activity. Copyright IQVIA. All Rights Reserved.

**Table A23.** Pricing courses by country (int$), treatment courses defined in Table A21*, data from the IQVIA MIDAS Quarterly sales data for the year 2019

| Country ISO-3 code | Amoxicillin | Amoxicillin Clavulanic acid | Doxycycline | Nitrofurantoin | Ciprofloxacin | Azithromycin | Cefalexin | Clindamycin | Phenoxymethy lpenicillin | Cefixime | Linezolid |
|---|---|---|---|---|---|---|---|---|---|---|---|
| NZL | 0.92 | 1.93 | 0.51 | 2.91 | 1.71 | 2.37 | 4.66 | 57.54 | 2.82 | | 1678.17 |
| HKG | 1.49 | 3.77 | 1.97 | 0.67 | 3.26 | 4.69 | 1.53 | 188.63 | 10.01 | | 1446.69 |
| AUS | 1.82 | 3.07 | 1.48 | 5.02 | 7.09 | 8.37 | 1.29 | 69.95 | 4.43 | | 322.15 |
| USA | 2.04 | 5.30 | 8.96 | 10.05 | 4.31 | 5.25 | 2.07 | 51.13 | 5.46 | 52.44 | 74.87 |
| KOR | 2.06 | 12.27 | 1.45 | 4.04 | 27.80 | 9.09 | 15.18 | 58.59 | | 6.46 | 642.88 |
| GBR | 2.25 | 4.67 | 1.58 | 10.21 | 7.17 | 4.87 | 2.26 | 141.58 | 10.04 | 14.63 | 912.80 |
| TWN | 2.42 | 8.59 | 0.98 | | 18.16 | 9.75 | 2.19 | 36.61 | | 3.82 | 913.24 |
| ESP | 2.60 | 4.77 | 1.95 | 2.58 | 5.09 | 8.14 | 4.47 | 49.70 | 19.62 | 3.73 | 1180.45 |
| CHN | 2.85 | 19.71 | 1.60 | 0.24 | 2.60 | 6.25 | 1.76 | 223.21 | 5.67 | 6.85 | 1165.74 |
| SGP | 2.87 | 8.88 | 0.73 | 1.64 | 12.54 | 13.15 | 3.23 | 71.53 | 0.70 | | 573.05 |
| VNM | 3.15 | 18.14 | 0.67 | 0.95 | 3.86 | 6.14 | 2.88 | 85.43 | 2.91 | 3.54 | 47.41 |
| DEU | 3.18 | 24.56 | 1.44 | 1.60 | 10.23 | 3.98 | 6.38 | 53.70 | 7.64 | 9.00 | 420.89 |
| NLD | 3.20 | 4.05 | 3.01 | 3.72 | 5.92 | 3.04 | 5.76 | 77.88 | 22.62 | | 125.92 |
| PER | 3.27 | 24.49 | 1.80 | 12.19 | 7.96 | 3.10 | 4.26 | 41.18 | 19.71 | 15.43 | 1671.96 |
| GRC | 3.37 | 8.58 | 3.55 | 2.63 | 18.26 | 8.07 | | 62.30 | 9.89 | 6.93 | 719.12 |
| MYS | 3.56 | 21.84 | 1.09 | 2.22 | 12.94 | 9.57 | 1.54 | 201.64 | 40.12 | 13.68 | 1855.50 |
| THA | 3.62 | 24.17 | 0.87 | 1.31 | 10.76 | 26.20 | 5.24 | 70.86 | 4.76 | 23.27 | 2331.44 |
| JPN | 3.66 | 22.19 | 1.75 | | 22.41 | 11.20 | 7.35 | 61.28 | | 15.45 | 1441.33 |
| LUX | 3.68 | 6.11 | 1.48 | 1.06 | 14.36 | 6.14 | 7.10 | 61.96 | 11.98 | | |
| LKA | 3.68 | 24.64 | 1.33 | 0.28 | 3.88 | 4.05 | 4.14 | 205.52 | 4.49 | 4.88 | 166.93 |
| SVK | 3.71 | 9.97 | 1.81 | | 8.63 | 8.53 | 4.95 | 66.51 | 5.90 | 6.04 | 463.29 |
| FRA | 3.73 | 7.27 | 2.45 | 2.88 | 27.84 | 13.13 | 6.79 | 87.18 | 13.36 | 6.61 | 1021.54 |
| ZAF | 3.79 | 17.25 | 0.62 | 12.36 | 7.07 | 8.37 | 3.76 | 168.06 | 6.89 | 11.37 | 150.84 |
| TUR | 3.80 | 5.60 | 2.22 | 2.30 | 6.21 | 6.58 | 4.45 | 111.01 | 12.32 | 2.92 | 365.29 |
| ITA | 3.88 | 9.25 | 4.08 | 6.43 | 32.40 | 8.84 | 6.05 | 129.58 | | 6.42 | 1446.33 |

| | | | | | | | | | | | |
|---|---|---|---|---|---|---|---|---|---|---|---|
| EGY | 3.99 | 13.29 | 3.07 | 1.68 | 16.63 | 9.46 | 4.96 | 139.59 | 12.03 | 11.03 | 33.78 |
| CZE | 4.14 | 7.87 | 3.32 | 3.19 | 14.50 | 6.33 | | 76.39 | 9.36 | 10.35 | 935.22 |
| BIH | 4.33 | 18.22 | 2.32 | 9.99 | 23.87 | 12.40 | 4.53 | 124.65 | 24.79 | 11.04 | |
| IDN | 4.38 | 47.57 | 3.58 | | 15.97 | 19.10 | 9.93 | 68.56 | 5.59 | 14.71 | 2471.05 |
| BEL | 4.47 | 6.58 | 2.25 | 1.19 | 16.29 | 6.42 | 7.97 | 71.83 | 12.60 | | 767.40 |
| SRB | 4.48 | 9.75 | 3.57 | 14.26 | 20.07 | 6.45 | 4.92 | 79.81 | | 4.90 | 1233.17 |
| PRI | 4.59 | 25.89 | 20.11 | 22.89 | 7.12 | 24.36 | 9.67 | 150.59 | 24.01 | 86.49 | 169.21 |
| PAK | 4.72 | 12.86 | 1.21 | 0.38 | 16.60 | 5.31 | 6.97 | 93.71 | | 5.51 | 41.88 |
| BGD | 4.75 | 22.30 | 0.66 | 5.38 | 12.43 | 4.82 | 5.73 | 78.45 | 5.48 | 5.92 | 51.49 |
| NOR | 4.89 | 14.32 | 3.36 | 7.34 | 8.08 | 6.30 | 3.53 | 76.07 | 4.74 | 164.23 | 360.18 |
| IRL | 4.93 | 6.60 | 3.40 | 3.00 | 22.09 | 12.08 | 2.72 | 128.95 | 7.47 | 11.43 | 570.68 |
| IND | 5.02 | 15.32 | 0.54 | 3.03 | 3.43 | 3.13 | 9.24 | 142.52 | | 2.32 | 22.37 |
| SWE | 5.04 | 6.19 | 3.57 | 2.74 | 7.21 | 7.61 | | 74.85 | 5.48 | | 118.34 |
| PRT | 5.05 | 7.51 | 4.20 | 2.47 | 24.26 | 8.86 | | 91.88 | | 6.63 | 68.04 |
| ROU | 5.08 | 8.01 | 1.61 | 3.14 | 13.89 | 10.26 | 7.34 | 83.18 | 5.93 | 10.96 | 807.18 |
| POL | 5.16 | 12.27 | 3.70 | 2.61 | 12.02 | 10.17 | 12.51 | 90.12 | 11.56 | 11.77 | 950.76 |
| TUN | 5.45 | 14.66 | 2.69 | 1.50 | 14.38 | 13.66 | | | 8.63 | 10.57 | 2017.35 |
| EST | 5.49 | 11.50 | 2.21 | 8.34 | 11.90 | 10.83 | | 92.77 | 15.05 | | |
| LBN | 5.54 | 10.46 | 4.85 | 2.29 | 25.58 | 15.01 | 12.51 | 104.94 | 12.61 | 12.95 | 152.21 |
| COL | 5.63 | 51.78 | 5.59 | 3.77 | 12.45 | 7.17 | 6.48 | 134.66 | 22.97 | | 619.81 |
| AUT | 5.71 | 8.47 | 2.58 | 2.39 | 33.64 | 8.12 | 7.73 | 69.01 | 13.15 | 6.88 | 539.70 |
| LTU | 6.02 | 15.56 | 3.01 | 4.52 | 16.64 | 11.44 | | 165.05 | 15.85 | | 108.03 |
| CHE | 6.04 | 18.16 | 4.79 | 1.53 | 30.99 | 11.64 | | 121.85 | 9.81 | | 1257.09 |
| CAN | 6.06 | 10.20 | 6.71 | 5.09 | 14.16 | 6.91 | 5.19 | 67.82 | 10.58 | 9.06 | 426.38 |
| VEN | 6.11 | 7.56 | 1.99 | 2.35 | 8.23 | 3.32 | 4.69 | 61.08 | | 3.41 | |
| HUN | 6.19 | 12.34 | 1.94 | 5.78 | 16.58 | 14.28 | 6.72 | 79.71 | 11.86 | 10.76 | 1432.75 |
| DZA | 6.28 | 17.12 | 3.08 | | 39.80 | 15.89 | 7.71 | | 10.02 | 18.05 | |
| LVA | 6.41 | 11.59 | 2.66 | 4.86 | 20.97 | 13.05 | | 161.60 | 21.69 | | 2048.23 |
| CHL | 6.44 | 21.09 | 15.07 | 4.17 | 7.37 | 5.98 | | 640.75 | 7.97 | 38.58 | 1944.27 |
| SVN | 6.54 | 16.11 | 3.44 | 18.11 | 25.85 | 7.60 | | 127.57 | 21.98 | 12.84 | 793.53 |
| RUS | 6.75 | 13.76 | 6.99 | 2.40 | 6.64 | 8.65 | 2.26 | 100.19 | 5.33 | 12.00 | 593.18 |
| BGR | 6.79 | 23.34 | 5.22 | | 19.82 | 19.76 | 7.48 | 122.38 | 10.28 | 21.38 | 439.15 |

| | | | | | | | | | | | |
|---|---|---|---|---|---|---|---|---|---|---|---|
| URY | 7.05 | 8.38 | 5.60 | 2.67 | 8.84 | 4.02 | 17.34 | 62.75 | | 6.49 | 162.93 |
| ARG | 7.20 | 15.51 | 8.55 | 9.22 | 38.69 | 12.73 | 8.61 | 146.44 | 25.22 | 18.32 | 498.03 |
| ECU | 7.40 | 24.94 | 3.26 | 3.40 | 29.99 | 11.92 | 6.91 | 100.89 | 10.36 | | 1178.13 |
| PHL | 7.89 | 36.54 | 10.44 | 28.22 | 22.84 | 23.24 | 14.35 | 365.24 | 30.21 | 27.12 | 3500.40 |
| MAR | 8.28 | 15.78 | 4.91 | | 40.83 | 14.20 | 13.07 | | 15.66 | 14.38 | |
| BRA | 8.78 | 32.30 | 4.63 | 1.18 | 22.60 | 10.23 | 11.73 | 201.45 | 19.30 | | 1389.23 |
| JOR | 9.25 | 17.82 | 5.77 | 2.66 | 40.07 | 27.92 | 6.72 | 152.65 | | 18.00 | 2207.85 |
| BLR | 9.62 | 17.66 | 5.76 | 0.96 | 10.38 | 10.19 | 5.31 | 112.10 | | 15.89 | 1008.27 |
| FIN | 9.85 | 11.16 | 3.05 | | 24.09 | 9.66 | 6.05 | 105.76 | 4.99 | | 271.66 |
| KWT | 9.87 | 22.13 | 7.66 | 3.00 | 59.39 | 20.43 | 12.40 | 102.73 | | 10.85 | |
| ARE | 11.02 | 26.84 | 7.85 | 2.58 | 54.96 | 21.48 | 5.78 | 65.47 | | 13.93 | 2024.10 |
| DOM | 12.10 | 44.62 | 11.78 | 13.62 | 48.03 | 19.49 | 19.73 | 308.33 | | 18.45 | 1300.64 |
| KAZ | 12.26 | 30.75 | 3.81 | 0.32 | 13.47 | 16.86 | 13.65 | 249.86 | | 14.04 | 66.58 |
| SAU | 13.79 | 26.87 | 10.46 | 3.12 | 49.56 | 20.15 | 6.89 | 175.23 | | 13.30 | 2269.60 |
| MEX | 15.82 | 25.45 | 20.06 | 8.75 | 40.40 | 20.10 | 18.01 | 196.17 | 6.61 | 30.32 | 1600.13 |
| HRV | 29.98 | 66.71 | 21.80 | 53.59 | 278.12 | 69.32 | 44.34 | 616.70 | 185.96 | 41.61 | 10053.59 |

Notes: *Values less than 0.005 were approximated to 0.00. Blank spaces mean country did not have data. int$= international dollars, 2019. Table A1 indicates the abbreviations of country-ISO3-codes. Authors' analysis of IQVIA MIDAS Quarterly Sales data for the year 2019, reflecting estimates of real-world activity. Copyright IQVIA. All Rights Reserved.

**Figure A23.** Percentage of the population at risk of failing under national poverty lines due to out-of-pocket expenditure for clindamycin and linezolid oral treatments following Table A21, by country, data from the IQVIA MIDAS Quarterly sales data for the year 2019

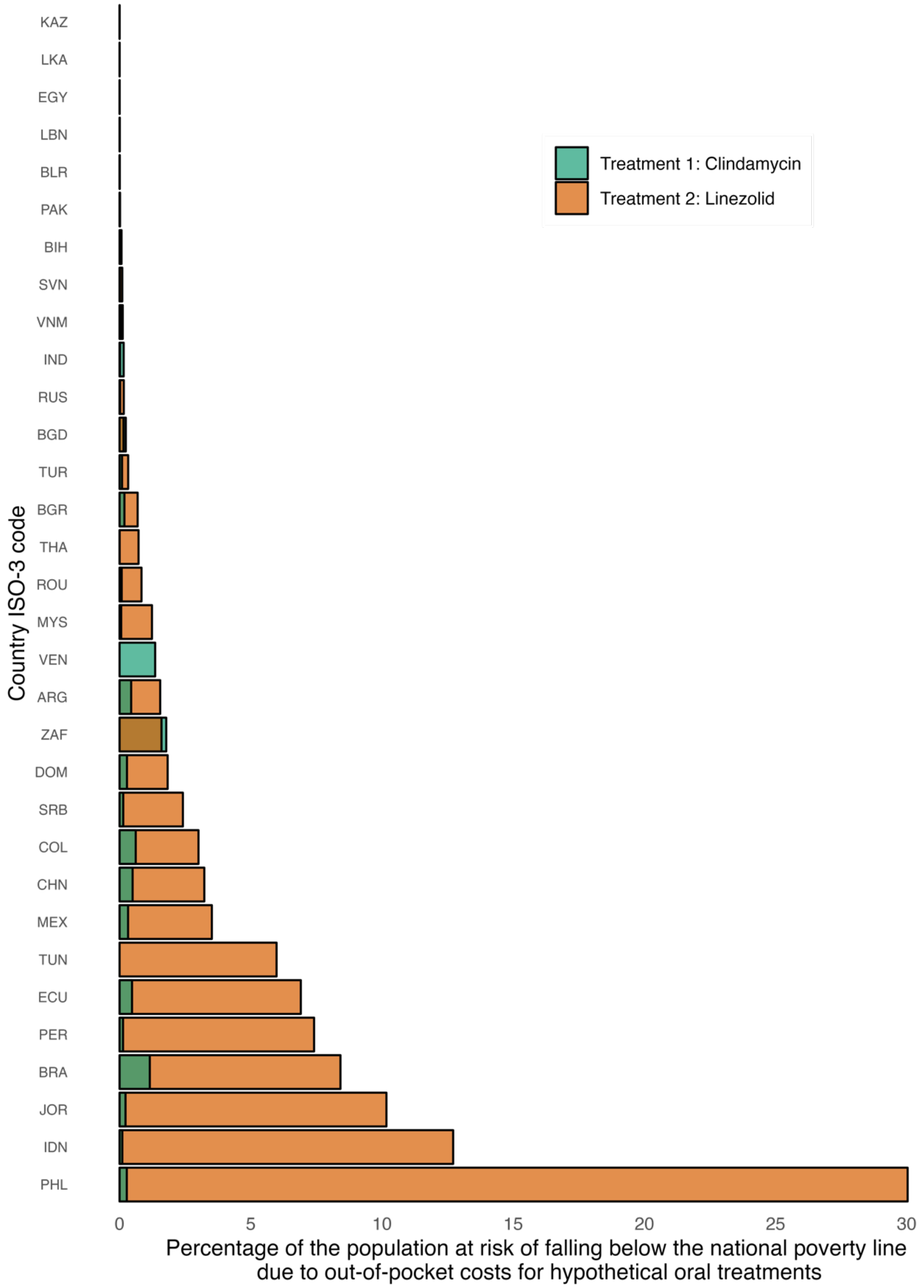

Notes: Countries without data on the selected specific antibiotics were excluded. Table A1 indicates the abbreviations of country-ISO3-codes. Authors' analysis of IQVIA MIDAS Quarterly Sales data for the year 2019, reflecting estimates of real-world activity. Copyright IQVIA. All Rights Reserved.

**Table A24.** Percentage of the population at risk of falling below the national poverty line due to out-of-pocket expenditure for hypothetical oral treatments (clindamycin and linezolid), data from the IQVIA MIDAS Quarterly sales data for the year 2019

| Country ISO-3 code | WB income | WHO region | Treatment 1 | Proportion at risk of poverty (%) | Number of people at risk | Treatment 2 | Proportion at risk of poverty (%) | Number of people at risk |
|---|---|---|---|---|---|---|---|---|
| PHL | LMIC | WPR | Clindamycin | 0.28 | 298040 | Linezolid | 30.03 | 32,471,825 |
| IDN | LMIC | SEAR | Clindamycin | 0.10 | 279003 | Linezolid | 12.71 | 33,986,308 |
| JOR | LMIC | EMR | Clindamycin | 0.23 | 23152 | Linezolid | 10.17 | 1,040,758 |
| BRA | LMIC | AMR | Clindamycin | 1.15 | 2359595 | Linezolid | 8.42 | 17,230,292 |
| PER | LMIC | AMR | Clindamycin | 0.13 | 41427 | Linezolid | 7.41 | 2,321,795 |
| ECU | LMIC | AMR | Clindamycin | 0.48 | 79634 | Linezolid | 6.91 | 1,157,609 |
| TUN | LMIC | AFR | Clindamycin | | | | 5.98 | |
| MEX | LMIC | AMR | Clindamycin | 0.32 | 400409 | Linezolid | 3.52 | 4,339,700 |
| CHN | LMIC | WPR | Clindamycin | 0.50 | 7022072 | Linezolid | 3.23 | 45,603,251 |
| COL | LMIC | AMR | Clindamycin | 0.62 | 296642 | Linezolid | 3.01 | 1,447,320 |
| SRB | LMIC | EUR | Clindamycin | 0.14 | 9933 | Linezolid | 2.41 | 170,685 |
| DOM | LMIC | AMR | Clindamycin | 0.28 | 30163 | Linezolid | 1.83 | 195,425 |
| ZAF | LMIC | AFR | Clindamycin | 1.78 | 1024637 | Linezolid | 1.60 | 921,333 |
| ARG | LMIC | AMR | Clindamycin | 0.44 | 195589 | Linezolid | 1.55 | 685,281 |
| VEN | LMIC | AMR | Clindamycin | 1.36 | 414361 | Linezolid | | |
| MYS | LMIC | WPR | Clindamycin | 0.06 | 20005 | Linezolid | 1.23 | 399,325 |
| ROU | LMIC | EUR | Clindamycin | 0.08 | 16038 | Linezolid | 0.84 | 164,539 |
| THA | LMIC | SEAR | Clindamycin | 0.00 | 346 | Linezolid | 0.72 | 514,236 |
| BGR | LMIC | EUR | Clindamycin | 0.19 | 13115 | Linezolid | 0.69 | 48,694 |
| TUR | LMIC | EUR | Clindamycin | 0.09 | 78648 | Linezolid | 0.33 | 271,849 |
| BGD | LMIC | SEAR | Clindamycin | 0.24 | 384188 | Linezolid | 0.15 | 243,928 |
| RUS | LMIC | EUR | Clindamycin | 0.02 | 32665 | Linezolid | 0.16 | 235,134 |
| IND | LMIC | SEAR | Clindamycin | 0.16 | 2121076 | Linezolid | 0.02 | 258,110 |
| VNM | LMIC | WPR | Clindamycin | 0.12 | 113618 | Linezolid | 0.06 | 61,545 |
| SVN | LMIC | EUR | Clindamycin | 0.01 | 301 | Linezolid | 0.10 | 2,112 |
| BIH | LMIC | EUR | Clindamycin | 0.07 | 2540 | Linezolid | | |
| PAK | LMIC | EMR | Clindamycin | 0.02 | 38311 | Linezolid | 0.01 | 14,014 |
| BLR | LMIC | EUR | Clindamycin | 0.00 | 28 | Linezolid | 0.01 | 728 |
| LBN | LMIC | EMR | Clindamycin | 0.00 | 304 | Linezolid | 0.01 | 461 |
| EGY | LMIC | EMR | Clindamycin | 0.00 | 3567 | Linezolid | 0.00 | 739 |

| | | | | | | | | |
|---|---|---|---|---|---|---|---|---|
| LKA | LMIC | SEAR | Clindamycin | 0.00 | 456 | Linezolid | 0.00 | 329 |
| KAZ | LMIC | EUR | Clindamycin | 0.00 | 0 | Linezolid | 0.00 | 0 |
| | | | **Median** | **0.13** | 38,311 | **Median** | **1.03** | 264,980 |
| | | | **Mean** | **0.29** | 493,544 | **Mean** | **3.44** | 4,816,143 |
| | | | **IQR** | **0.28** | 290,591 | **IQR** | **3.37** | 1,076,490 |

Notes: LMICs= low-and-middle income countries. WHO= World health organization. Table A1 indicates the abbreviations of country-ISO3-codes. Table A30 explains treatment details for clindamycin and linezolid. Authors' analysis of IQVIA MIDAS Quarterly Sales data for the year 2019, reflecting estimates of real-world activity. Copyright IQVIA. All Rights Reserved.

**Table A25.** Price for a full course of antibiotic treatment for sepsis of unknown origin when following the AWaRe guidance, by income level and adults and neonates (int$), data from the IQVIA MIDAS Quarterly sales data for the year 2019

| Treatment group/WB income group | HICs | | | | LMICs | | | |
|---|---|---|---|---|---|---|---|---|
| | Median | p25th | p75th | IQR | Median | p25th | p75th | IQR |
| **I. Adults (reference body weight of 70 kilograms), treatment course duration of 7 days** | | | | | | | | |
| Group 1: Ceftriaxone, Amikacin/Gentamicin | 204.53 | 119.00 | 413.12 | 294.12 | 126.11 | 79.93 | 264.19 | 184.27 |
| Group 2: Piperacillin+Tazobactam | 369.21 | 230.40 | 543.25 | 312.85 | 557.41 | 330.85 | 1108.30 | 777.46 |
| Group 3: Meropenem | 759.26 | 437.64 | 1181.87 | 744.24 | 1567.06 | 718.67 | 2127.84 | 1409.16 |
| Group 4: Colistin | 557.71 | 378.95 | 891.94 | 512.99 | 1076.80 | 428.67 | 1662.41 | 1233.73 |
| **II. Neonates (reference body weight of 3.5 kilograms), treatment course duration of 7 days** | | | | | | | | |
| **Treatment group/WB income group** | **HICs** | | | | **LMICs** | | | |
| | Median | p25th | p75th | IQR | Median | p25th | p75th | IQR |
| Group 1: Ampicillin/Benzylpenicillin, Gentamicin | 10.66 | 5.06 | 21.54 | 16.49 | 3.98 | 1.90 | 6.50 | 4.60 |
| Group 2: Ceftriaxone | 9.63 | 4.21 | 21.79 | 17.58 | 12.83 | 5.19 | 20.82 | 15.63 |
| Group 3: Piperacillin+Tazobactam | 21.47 | 13.40 | 31.58 | 18.19 | 32.41 | 19.24 | 64.44 | 45.20 |
| Group 4: Meropenem | 53.15 | 30.63 | 82.73 | 52.10 | 109.69 | 50.31 | 148.95 | 98.64 |
| Group 5: Colistin | 21.47 | 14.59 | 34.34 | 19.75 | 41.46 | 16.50 | 64.00 | 47.50 |

Notes: WB= World bank income groups. HIC= high-income countries. LMIC= Low-and middle-income countries (here it is just middle-income countries as there is no low-income countries). IQR= interquartile range. Int$= International dollars, 2019. Authors' analysis of IQVIA MIDAS Quarterly Sales data for the year 2019, reflecting estimates of real-world activity. Copyright IQVIA. All Rights Reserved.

**Table A26.** Hypothetical treatment prices for unknown sepsis, by WHO region and adults and neonates (int$), data from the IQVIA MIDAS Quarterly sales data for the year 2019

**I. Adults (reference body weight of 70 kilograms), treatment course duration of 7 days**

| Treatment group/ WHO region | AFRO | | | | AMRO | | | | EMRO | | | |
|---|---|---|---|---|---|---|---|---|---|---|---|---|
| | Median | p25th | p75th | IQR | Median | p25th | p75th | IQR | Median | p25th | p75th | IQR |
| Group 1: Ceftriaxone, Amikacin/Gentamicin | 117.62 | 100.40 | 160.85 | 60.45 | 181.43 | 91.87 | 297.99 | 206.12 | 283.24 | 213.83 | 361.57 | 147.74 |
| Group 2: Piperacillin+Tazobactam | 479.38 | 349.31 | 538.51 | 189.20 | 466.00 | 209.47 | 1472.89 | 1263.42 | 774.76 | 569.84 | 1006.28 | 436.44 |
| Group 3: Meropenem | 756.73 | 717.16 | 1011.56 | 294.40 | 1567.06 | 400.16 | 2625.31 | 2225.15 | 1807.81 | 1617.78 | 1941.73 | 323.95 |
| Group 4: Colistin | 361.29 | 207.63 | 572.90 | 365.27 | 457.32 | 370.45 | 1582.88 | 1212.43 | 1362.72 | 1125.36 | 1766.51 | 641.16 |

| Treatment group/ WHO region | EURO | | | | SEARO | | | | WPRO | | | |
|---|---|---|---|---|---|---|---|---|---|---|---|---|
| | Median | p25th | p75th | IQR | Median | p25th | p75th | IQR | Median | p25th | p75th | IQR |
| Group 1: Ceftriaxone, Amikacin/Gentamicin | 128.83 | 80.65 | 292.78 | 212.12 | 121.86 | 113.98 | 129.48 | 15.50 | 258.91 | 131.46 | 470.02 | 338.56 |
| Group 2: Piperacillin+Tazobactam | 389.74 | 271.37 | 538.55 | 267.18 | 1424.55 | 763.56 | 2105.00 | 1341.44 | 408.02 | 260.46 | 596.85 | 336.39 |
| Group 3: Meropenem | 737.23 | 472.46 | 992.80 | 520.34 | 2020.44 | 1718.02 | 2106.40 | 388.38 | 822.37 | 614.43 | 1625.94 | 1011.51 |
| Group 4: Colistin | 610.56 | 375.33 | 1148.14 | 772.80 | 401.15 | 304.30 | 610.08 | 305.77 | 891.94 | 748.19 | 1295.89 | 547.70 |

**II. Neonates (reference body weight of 3.5 kilograms), treatment course duration of 7 days**

| Treatment group/ WHO region | AFRO | | | | AMRO | | | | EMRO | | | |
|---|---|---|---|---|---|---|---|---|---|---|---|---|
| | Median | p25th | p75th | IQR | Median | p25th | p75th | IQR | Median | p25th | p75th | IQR |
| Group 1: Ampicillin/Benzylpenicillin, Gentamicin | 5.09 | 4.93 | 5.53 | 0.60 | 4.54 | 2.82 | 8.81 | 5.99 | 5.64 | 2.19 | 23.48 | 21.28 |

| | | | | | | | | | | | | |
|---|---|---|---|---|---|---|---|---|---|---|---|---|
| Group 2: Ceftriaxone | 12.03 | 8.79 | 17.86 | 9.07 | 14.80 | 4.83 | 34.70 | 29.87 | 24.11 | 21.77 | 34.20 | 12.43 |
| Group 3: Piperacillin+Tazobactam | 27.87 | 20.31 | 31.31 | 11.00 | 27.09 | 12.18 | 85.63 | 73.45 | 45.04 | 33.13 | 58.50 | 25.37 |
| Group 4: Meropenem | 52.97 | 50.20 | 70.81 | 20.61 | 109.69 | 28.01 | 183.77 | 155.76 | 126.55 | 113.24 | 135.92 | 22.68 |
| Group 5: Colistin | 13.91 | 7.99 | 22.06 | 14.06 | 17.61 | 14.26 | 60.94 | 46.68 | 52.46 | 43.33 | 68.01 | 24.68 |

| **Treatment group/ WHO region** | **EURO** | | | | **SEARO** | | | | **WPRO** | | | |
|---|---|---|---|---|---|---|---|---|---|---|---|---|
| | Median | p25th | p75th | IQR | Median | p25th | p75th | IQR | Median | p25th | p75th | IQR |
| Group 1: Ampicillin/Benzylpenicillin, Gentamicin | 8.84 | 4.76 | 19.07 | 14.31 | 2.05 | 1.43 | 2.90 | 1.47 | 4.33 | 1.44 | 14.98 | 13.54 |
| Group 2: Ceftriaxone | 7.94 | 3.66 | 11.90 | 8.23 | 12.30 | 12.06 | 15.71 | 3.65 | 16.86 | 10.29 | 19.92 | 9.63 |
| Group 3: Piperacillin+Tazobactam | 22.66 | 15.78 | 31.31 | 15.53 | 82.82 | 44.39 | 122.38 | 77.99 | 23.72 | 15.14 | 34.70 | 19.56 |
| Group 4: Meropenem | 51.61 | 33.07 | 69.50 | 36.42 | 141.43 | 120.26 | 147.45 | 27.19 | 57.57 | 43.01 | 113.82 | 70.81 |
| Group 5: Colistin | 23.51 | 14.45 | 44.20 | 29.75 | 15.44 | 11.72 | 23.49 | 11.77 | 34.34 | 28.81 | 49.89 | 21.09 |

Notes: WB= World bank income groups. HIC= high-income countries. LMIC= Lower middle-income countries. IQR= interquartile range. WHO= World Health Organization. AFRO= African region, AMRO= Americas region, EMRO= East Mediterranean region, EURO= Europe, SEARO= Southeast Asia region, WPRO= Western-pacific region. Int$= International dollars, 2019. Authors' analysis of IQVIA MIDAS Quarterly Sales data for the year 2019, reflecting estimates of real-world activity. Copyright IQVIA. All Rights Reserved.

**Table A27.** Antibiotic treatment price for a 7-day duration sepsis, unknown causative agent, by country and regimen (int$), among adults (70 kg adult as an example), data from the IQVIA MIDAS Quarterly sales data for the year 2019

| Country ISO-3 code | I. Adults | | | | | | | |
|---|---|---|---|---|---|---|---|---|
| | Ceftriaxone + Gentamicin/Amikacin | | Piperacillin+Tazobactam | | Meropenem | | Colistin | |
| | **per day** | **full course** | **per day** | **full course** | **per day** | **full course** | **per day** | **full course** |
| ARE | 42.57 | 297.99 | 92.33 | 646.28 | 243.42 | 1703.92 | | |
| ARG | 64.87 | 454.08 | 210.41 | 1472.89 | 713.15 | 4992.06 | 61.94 | 433.61 |
| AUS | 75.70 | 529.90 | 16.95 | 118.62 | 32.82 | 229.72 | 137.72 | 964.07 |
| AUT | 54.41 | 380.90 | 136.72 | 957.01 | 230.76 | 1615.35 | 208.44 | 1459.05 |
| BEL | 41.83 | 292.78 | 49.74 | 348.21 | 90.55 | 633.87 | 60.81 | 425.65 |
| BGD | 16.28 | 113.98 | 130.43 | 913.01 | 236.51 | 1655.56 | 171.25 | 1198.76 |
| BGR | 16.16 | 113.13 | 37.75 | 264.26 | 86.79 | 607.54 | 103.24 | 722.68 |
| BIH | 12.36 | 86.55 | 117.17 | 820.17 | 131.62 | 921.36 | 206.65 | 1446.52 |
| BLR | 4.22 | 29.55 | 523.58 | 3665.03 | 86.23 | 603.59 | 167.47 | 1172.27 |
| BRA | 25.92 | 181.43 | 160.26 | 1121.81 | 401.82 | 2812.74 | 40.68 | 284.77 |
| CAN | 137.71 | 963.98 | 14.37 | 100.56 | 64.87 | 454.09 | 52.92 | 370.45 |
| CHE | 62.01 | 434.09 | 75.59 | 529.14 | 144.31 | 1010.20 | | |
| CHL | 87.81 | 614.65 | 212.76 | 1489.32 | 239.96 | 1679.74 | | |
| CHN | 11.76 | 82.34 | 58.29 | 408.02 | 211.41 | 1479.84 | | |
| COL | 13.12 | 91.87 | 29.92 | 209.47 | 33.00 | 231.03 | 586.29 | 4104.06 |
| CZE | 18.47 | 129.32 | 63.08 | 441.54 | 118.19 | 827.30 | 49.55 | 346.84 |
| DEU | 31.04 | 217.31 | 15.73 | 110.09 | 38.26 | 267.84 | 52.07 | 364.48 |
| DOM | 41.66 | 291.63 | 231.62 | 1621.31 | 478.88 | 3352.17 | | |
| DZA | | | | | | | | |
| ECU | 23.13 | 161.88 | 53.96 | 377.73 | 223.87 | 1567.06 | 140.19 | 981.33 |
| EGY | 15.98 | 111.86 | 68.48 | 479.38 | 180.91 | 1266.40 | 19.22 | 134.56 |
| ESP | 15.99 | 111.94 | 44.07 | 308.50 | 123.29 | 863.06 | 63.12 | 441.81 |
| EST | 8.60 | 60.23 | | | | | 55.41 | 387.90 |
| FIN | 141.10 | 987.73 | 46.15 | 323.05 | 42.15 | 295.06 | | |
| FRA | 41.72 | 292.05 | 78.28 | 547.95 | 105.32 | 737.23 | 113.91 | 797.39 |
| GBR | 51.62 | 361.37 | 88.86 | 622.05 | 139.34 | 975.40 | 22.80 | 159.62 |
| GRC | 24.99 | 174.91 | 32.76 | 229.30 | 82.59 | 578.14 | 114.04 | 798.30 |
| HKG | 38.85 | 271.96 | 16.90 | 118.32 | 86.50 | 605.52 | 116.81 | 817.65 |
| HRV | 321.52 | 2250.61 | 792.45 | 5547.12 | 1167.56 | 8172.92 | 1197.24 | 8380.65 |
| HUN | 17.22 | 120.54 | 57.25 | 400.76 | 168.30 | 1178.10 | 54.14 | 378.95 |
| IDN | 67.56 | 472.93 | 373.11 | 2611.78 | 288.63 | 2020.44 | 7.41 | 51.87 |
| IND | 9.65 | 67.54 | 45.03 | 315.22 | 245.43 | 1718.02 | 59.12 | 413.85 |
| IRL | 27.39 | 191.75 | 57.10 | 399.69 | 224.95 | 1574.62 | 27.47 | 192.26 |

| | | | | | | | | |
|---|---|---|---|---|---|---|---|---|
| ITA | 19.31 | 135.16 | 55.68 | 389.74 | 111.61 | 781.30 | 162.87 | 1140.09 |
| JOR | 40.92 | 286.43 | 152.54 | 1067.80 | 266.91 | 1868.36 | 194.67 | 1362.72 |
| JPN | 19.79 | 138.50 | 41.03 | 287.23 | 58.70 | 410.89 | 59.79 | 418.54 |
| KAZ | 8.25 | 57.72 | 66.74 | 467.18 | 319.32 | 2235.24 | 363.28 | 2542.95 |
| KOR | 17.77 | 124.41 | 33.38 | 233.68 | 104.82 | 733.74 | 129.21 | 904.46 |
| KWT | 55.23 | 386.61 | 103.97 | 727.80 | 249.61 | 1747.26 | | |
| LBN | 40.01 | 280.06 | 182.88 | 1280.19 | 280.88 | 1966.19 | 311.96 | 2183.69 |
| LKA | 18.50 | 129.48 | | | 602.69 | 4218.83 | | |
| LTU | 6.92 | 48.44 | 20.88 | 146.13 | 36.20 | 253.37 | 49.61 | 347.24 |
| LUX | 24.75 | 173.23 | | | | | | |
| LVA | 10.49 | 73.46 | 75.43 | 528.02 | 61.74 | 432.15 | 55.75 | 390.28 |
| MAR | 39.03 | 273.24 | | | 96.80 | 677.59 | 70.08 | 490.59 |
| MEX | 31.95 | 223.64 | 342.12 | 2394.84 | 375.04 | 2625.31 | 376.72 | 2637.05 |
| MYS | 36.99 | 258.91 | 115.46 | 808.21 | 117.48 | 822.37 | 101.61 | 711.30 |
| NLD | 58.59 | 410.14 | 75.48 | 528.34 | 158.42 | 1108.97 | 127.42 | 891.94 |
| NOR | 82.62 | 578.31 | 44.52 | 311.67 | 89.05 | 623.33 | 112.15 | 785.08 |
| NZL | 142.35 | 996.42 | 21.47 | 150.28 | 37.19 | 260.34 | 92.86 | 649.99 |
| PAK | 27.28 | 190.97 | 73.88 | 517.19 | 280.92 | 1966.43 | 252.36 | 1766.51 |
| PER | 11.11 | 77.75 | 25.91 | 181.37 | 57.17 | 400.16 | 226.13 | 1582.88 |
| PHL | 101.70 | 711.88 | 208.29 | 1458.00 | 479.58 | 3357.07 | 463.78 | 3246.44 |
| POL | 12.19 | 85.30 | 39.89 | 279.24 | 71.30 | 499.10 | 81.59 | 571.13 |
| PRI | 25.72 | 180.07 | 66.57 | 466.00 | 169.02 | 1183.13 | 65.33 | 457.32 |
| PRT | 7.44 | 52.05 | 14.44 | 101.07 | 40.13 | 280.91 | 79.67 | 557.71 |
| ROU | 18.40 | 128.83 | 39.59 | 277.15 | 184.80 | 1293.58 | 96.40 | 674.77 |
| RUS | 3.60 | 25.23 | 90.00 | 630.02 | 126.02 | 882.12 | | |
| SAU | 60.29 | 422.06 | 117.39 | 821.73 | 187.41 | 1311.87 | 160.77 | 1125.36 |
| SGP | 8.24 | 57.70 | 31.31 | 219.14 | 117.81 | 824.65 | | |
| SRB | 12.30 | 86.08 | 49.81 | 348.68 | 87.04 | 609.26 | 179.59 | 1257.13 |
| SVK | 37.31 | 261.18 | 138.23 | 967.59 | 97.23 | 680.62 | 45.11 | 315.77 |
| SVN | 76.88 | 538.15 | 67.91 | 475.35 | 185.16 | 1296.15 | 172.29 | 1206.03 |
| SWE | 79.89 | 559.26 | 72.44 | 507.05 | 175.97 | 1231.79 | 146.84 | 1027.90 |
| THA | 17.41 | 121.86 | 276.58 | 1936.08 | 300.91 | 2106.40 | 55.49 | 388.44 |
| TUN | 17.63 | 123.39 | 85.38 | 597.64 | | | 117.12 | 819.85 |
| TUR | 11.52 | 80.65 | 37.94 | 265.59 | 63.69 | 445.81 | 24.93 | 174.48 |
| TWN | 24.15 | 169.08 | 95.05 | 665.37 | 411.50 | 2880.52 | 101.19 | 708.34 |
| URY | 4.01 | 28.08 | 28.92 | 202.44 | 48.07 | 336.49 | 47.90 | 335.32 |
| USA | 16.34 | 114.39 | 47.99 | 335.92 | 51.94 | 363.57 | 33.11 | 231.75 |
| VEN | 6.23 | 43.59 | 33.41 | 233.89 | 46.55 | 325.87 | | |
| VNM | 10.78 | 75.45 | 66.04 | 462.27 | 253.15 | 1772.03 | 232.53 | 1627.70 |
| ZAF | 9.43 | 66.04 | 31.32 | 219.24 | 108.10 | 756.73 | 33.14 | 231.99 |

Notes: Blanks indicate data no available. Int$= International dollars, 2019. Table A1 indicates the abbreviations of country-ISO3-codes. Authors' analysis of IQVIA MIDAS Quarterly Sales data for the year 2019, reflecting estimates of real-world activity. Copyright IQVIA. All Rights Reserved.

**Table A28.** Antibiotic treatment price for a 7-day duration sepsis, unknown causative agent, by country and regimen (int$), among neonates (3.5kg neonate as an example), data from the IQVIA MIDAS Quarterly sales data for the year 2019

| **Country ISO-3 code** | **II. Children** | | | | | | | | | |
|---|---|---|---|---|---|---|---|---|---|---|
| | Ampicillin/ Benzyhpenicillin + gentamicin | | Ceftriaxone | | Piperacillin+Tazobactam | | Meropenem | | Colistin | |
| | **per day** | **full course** | **per day** | **full course** | **per day** | **full course** | **per day** | **full course** | **per day** | **full course** |
| ARE | | | 5.24 | 36.67 | 5.37 | 37.57 | 17.04 | 119.27 | | |
| ARG | 1.17 | 8.19 | 5.10 | 35.72 | 12.23 | 85.63 | 49.92 | 349.44 | 2.38 | 16.69 |
| AUS | 0.74 | 5.19 | 0.24 | 1.65 | 0.99 | 6.90 | 2.30 | 16.08 | 5.30 | 37.12 |
| AUT | 2.72 | 19.02 | 3.61 | 25.30 | 7.95 | 55.64 | 16.15 | 113.07 | 8.02 | 56.17 |
| BEL | 3.09 | 21.63 | 1.70 | 11.90 | 2.89 | 20.24 | 6.34 | 44.37 | 2.34 | 16.39 |
| BGD | 0.55 | 3.86 | 1.76 | 12.30 | 7.58 | 53.08 | 16.56 | 115.89 | 6.59 | 46.15 |
| BGR | 2.17 | 15.20 | 0.50 | 3.52 | 2.19 | 15.36 | 6.08 | 42.53 | 3.97 | 27.82 |
| BIH | 0.27 | 1.90 | 0.52 | 3.66 | 6.81 | 47.68 | 9.21 | 64.50 | 7.96 | 55.69 |
| BLR | 0.14 | 1.01 | 0.37 | 2.57 | 30.44 | 213.08 | 6.04 | 42.25 | 6.45 | 45.13 |
| BRA | 0.58 | 4.09 | 3.13 | 21.90 | 9.32 | 65.22 | 28.13 | 196.89 | 1.57 | 10.96 |
| CAN | 3.66 | 25.64 | 0.61 | 4.26 | 0.84 | 5.85 | 4.54 | 31.79 | 2.04 | 14.26 |
| CHE | 38.15 | 267.07 | 3.22 | 22.56 | 4.39 | 30.76 | 10.10 | 70.71 | | |
| CHL | 0.31 | 2.17 | 11.84 | 82.90 | 12.37 | 86.59 | 16.80 | 117.58 | | |
| CHN | 0.10 | 0.71 | 1.48 | 10.34 | 3.39 | 23.72 | 14.80 | 103.59 | 288.43 | 2019.01 |
| COL | 0.29 | 2.02 | 1.35 | 9.47 | 1.74 | 12.18 | 2.31 | 16.17 | 22.57 | 158.01 |
| CZE | 1.30 | 9.12 | 1.13 | 7.94 | 3.67 | 25.67 | 8.27 | 57.91 | 1.91 | 13.35 |
| DEU | 1.09 | 7.64 | 0.58 | 4.07 | 0.91 | 6.40 | 2.68 | 18.75 | 2.00 | 14.03 |
| DOM | 0.93 | 6.50 | 4.96 | 34.70 | 13.47 | 94.26 | 33.52 | 234.65 | | |
| DZA | 0.73 | 5.09 | | | | | | | | |
| ECU | 0.25 | 1.73 | 2.11 | 14.80 | 3.14 | 21.96 | 15.67 | 109.69 | 5.40 | 37.78 |
| EGY | 0.32 | 2.26 | 1.53 | 10.70 | 3.98 | 27.87 | 12.66 | 88.65 | 0.74 | 5.18 |
| ESP | 0.57 | 4.01 | 1.35 | 9.47 | 2.56 | 17.94 | 8.63 | 60.41 | 2.43 | 17.01 |
| EST | 1.49 | 10.45 | 0.51 | 3.58 | | | | | 2.13 | 14.93 |
| FIN | 12.45 | 87.16 | 1.09 | 7.66 | 2.68 | 18.78 | 2.95 | 20.65 | | |
| FRA | 2.75 | 19.23 | 3.06 | 21.44 | 4.55 | 31.86 | 7.37 | 51.61 | 4.39 | 30.70 |
| GBR | 1.55 | 10.87 | 3.50 | 24.53 | 5.17 | 36.17 | 9.75 | 68.28 | 0.88 | 6.15 |
| GRC | 2.03 | 14.21 | 1.40 | 9.79 | 1.90 | 13.33 | 5.78 | 40.47 | 4.39 | 30.73 |
| HKG | 0.17 | 1.17 | 2.62 | 18.31 | 0.98 | 6.88 | 6.06 | 42.39 | 4.50 | 31.48 |
| HRV | 22.07 | 154.48 | 9.82 | 68.71 | 46.07 | 322.51 | 81.73 | 572.10 | 46.09 | 322.66 |
| HUN | 0.96 | 6.73 | 0.92 | 6.46 | 3.33 | 23.30 | 11.78 | 82.47 | 2.08 | 14.59 |
| IDN | 0.37 | 2.58 | 2.24 | 15.71 | 21.69 | 151.85 | 20.20 | 141.43 | 0.29 | 2.00 |
| IND | 0.17 | 1.16 | 0.56 | 3.94 | 2.62 | 18.33 | 17.18 | 120.26 | 2.28 | 15.93 |

| | | | | | | | | | | |
|---|---|---|---|---|---|---|---|---|---|---|
| IRL | 1.24 | 8.69 | 3.40 | 23.80 | 3.32 | 23.24 | 15.75 | 110.22 | 1.06 | 7.40 |
| ITA | 3.32 | 23.26 | 1.59 | 11.14 | 3.24 | 22.66 | 7.81 | 54.69 | 6.27 | 43.89 |
| JOR | 0.37 | 2.59 | 3.49 | 24.41 | 8.87 | 62.08 | 18.68 | 130.79 | 7.49 | 52.46 |
| JPN | 2.03 | 14.22 | 0.89 | 6.24 | 2.39 | 16.70 | 4.11 | 28.76 | 2.30 | 16.11 |
| KAZ | 0.13 | 0.94 | 0.52 | 3.66 | 3.88 | 27.16 | 22.35 | 156.47 | 13.99 | 97.90 |
| KOR | 0.44 | 3.11 | 1.81 | 12.69 | 1.94 | 13.59 | 7.34 | 51.36 | 4.97 | 34.82 |
| KWT | 4.06 | 28.41 | 5.35 | 37.46 | 6.04 | 42.31 | 17.47 | 122.31 | | |
| LBN | 0.29 | 2.06 | 3.01 | 21.09 | 10.63 | 74.43 | 19.66 | 137.63 | 12.01 | 84.07 |
| LKA | | | 2.49 | 17.45 | | | 42.19 | 295.32 | | |
| LTU | 0.72 | 5.01 | 0.33 | 2.33 | 1.21 | 8.50 | 2.53 | 17.74 | 1.91 | 13.37 |
| LUX | | | 1.62 | 11.36 | | | | | | |
| LVA | 0.55 | 3.83 | 1.34 | 9.41 | 4.39 | 30.70 | 4.32 | 30.25 | 2.15 | 15.03 |
| MAR | 0.70 | 4.93 | 4.48 | 31.35 | | | 6.78 | 47.43 | 2.70 | 18.89 |
| MEX | 1.52 | 10.67 | 3.10 | 21.69 | 19.89 | 139.23 | 26.25 | 183.77 | 14.50 | 101.53 |
| MYS | 0.25 | 1.72 | 3.69 | 25.81 | 6.71 | 46.99 | 8.22 | 57.57 | 3.91 | 27.39 |
| NLD | 3.12 | 21.82 | 3.08 | 21.53 | 4.39 | 30.72 | 11.09 | 77.63 | 4.91 | 34.34 |
| NOR | 2.68 | 18.78 | 2.58 | 18.08 | 2.59 | 18.12 | 6.23 | 43.63 | 4.32 | 30.23 |
| NZL | 2.47 | 17.28 | 0.01 | 0.08 | 1.25 | 8.74 | 2.60 | 18.22 | 3.57 | 25.02 |
| PAK | 0.27 | 1.89 | 2.02 | 14.17 | 4.30 | 30.07 | 19.66 | 137.65 | 9.72 | 68.01 |
| PER | 0.71 | 4.98 | 1.02 | 7.13 | 1.51 | 10.54 | 4.00 | 28.01 | 8.71 | 60.94 |
| PHL | 2.25 | 15.74 | 8.17 | 57.19 | 12.11 | 84.77 | 33.57 | 234.99 | 17.86 | 124.99 |
| POL | 3.04 | 21.29 | 0.56 | 3.91 | 2.32 | 16.24 | 4.99 | 34.94 | 3.14 | 21.99 |
| PRI | 1.71 | 11.99 | 0.69 | 4.83 | 3.87 | 27.09 | 11.83 | 82.82 | 2.52 | 17.61 |
| PRT | 1.22 | 8.56 | 0.45 | 3.14 | 0.84 | 5.88 | 2.81 | 19.66 | 3.07 | 21.47 |
| ROU | 0.30 | 2.08 | 1.72 | 12.06 | 2.30 | 16.11 | 12.94 | 90.55 | 3.71 | 25.98 |
| RUS | 0.07 | 0.52 | 0.21 | 1.49 | 5.23 | 36.63 | 8.82 | 61.75 | | |
| SAU | 4.90 | 34.29 | 6.06 | 42.44 | 6.83 | 47.78 | 13.12 | 91.83 | 6.19 | 43.33 |
| SGP | 0.90 | 6.28 | 1.15 | 8.06 | 1.82 | 12.74 | 8.25 | 57.73 | | |
| SRB | 0.36 | 2.54 | 0.57 | 4.01 | 2.90 | 20.27 | 6.09 | 42.65 | 6.91 | 48.40 |
| SVK | 0.96 | 6.73 | 2.07 | 14.47 | 8.04 | 56.26 | 6.81 | 47.64 | 1.74 | 12.16 |
| SVN | 2.72 | 19.02 | 2.96 | 20.73 | 3.95 | 27.64 | 12.96 | 90.73 | 6.63 | 46.43 |
| SWE | 6.77 | 47.41 | 1.35 | 9.43 | 4.21 | 29.48 | 12.32 | 86.23 | 5.65 | 39.57 |
| THA | 0.22 | 1.52 | 1.72 | 12.06 | 16.08 | 112.56 | 21.06 | 147.45 | 2.14 | 14.96 |
| TUN | 0.79 | 5.53 | 1.91 | 13.36 | 4.96 | 34.75 | | | 4.51 | 31.56 |
| TUR | 0.93 | 6.50 | 0.80 | 5.60 | 2.21 | 15.44 | 4.46 | 31.21 | 0.96 | 6.72 |
| TWN | 0.62 | 4.33 | 2.41 | 16.86 | 5.53 | 38.68 | 28.81 | 201.64 | 3.90 | 27.27 |
| URY | 0.43 | 3.04 | 0.23 | 1.61 | 1.68 | 11.77 | 3.36 | 23.55 | 1.84 | 12.91 |
| USA | 1.05 | 7.35 | 0.50 | 3.53 | 2.79 | 19.53 | 3.64 | 25.45 | 1.27 | 8.92 |
| VEN | 0.59 | 4.10 | 0.21 | 1.44 | 1.94 | 13.60 | 3.26 | 22.81 | | |

| | | | | | | | | | |
|---|---|---|---|---|---|---|---|---|---|
| VNM | 0.10 | 0.72 | 1.46 | 10.24 | 3.84 | 26.88 | 17.72 | 124.04 | 8.95 | 62.67 |
| ZAF | 1.43 | 10.03 | 0.44 | 3.05 | 1.82 | 12.75 | 7.57 | 52.97 | 1.28 | 8.93 |

Notes: Blanks indicate data no available. Int$= International dollars, 2019. Table A1 indicates the abbreviations of country-ISO3-codes. Authors' analysis of IQVIA MIDAS Quarterly Sales data for the year 2019, reflecting estimates of real-world activity. Copyright IQVIA. All Rights Reserved.

**Figure A24.** Percentage of the population at risk of failing under national poverty lines due to out-of-pocket expenditure for sepsis treatment of unknown causative agent, by country, data from the IQVIA MIDAS Quarterly sales data for the year 2019

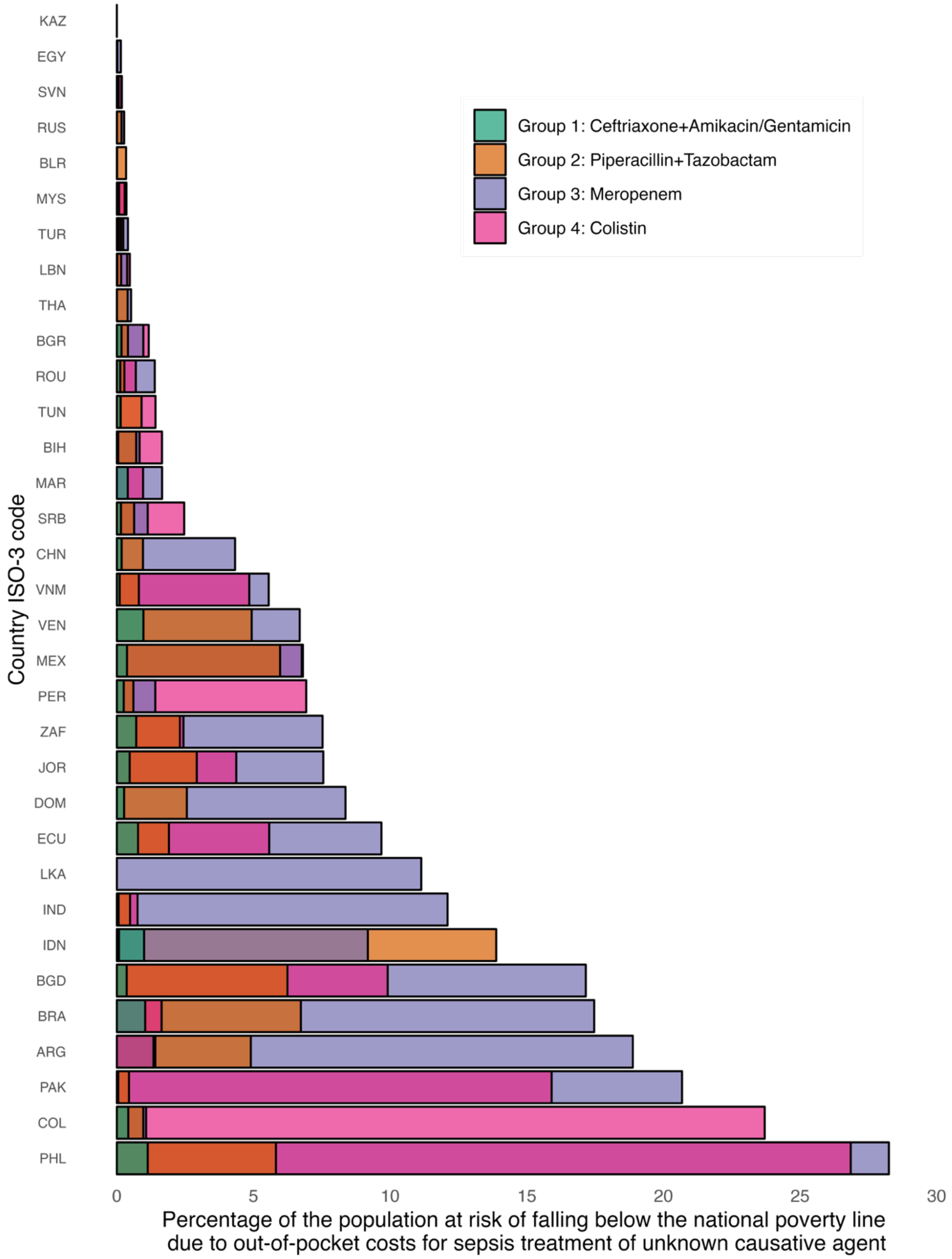

Notes: Countries without data on the selected specific antibiotics were excluded. Table A1 indicates the abbreviations of country-ISO3-codes. Authors' analysis of IQVIA MIDAS Quarterly Sales data for the year 2019, reflecting estimates of real-world activity. Copyright IQVIA. All Rights Reserved.

**Table A29.** Percentage of the population at risk of falling below the national poverty line due to out-of-pocket expenditure for sepsis treatment of unknown causative agent among adults, by treatment group†, data from the IQVIA MIDAS Quarterly sales data for the year 2019

| ISO code | Income level | Group 1† | Group 2† | Group 3† | Group 4† | N people Group 1† | N people Group 2† | N people Group 3† | N people Group 4† |
|---|---|---|---|---|---|---|---|---|---|
| ARG | LMIC | 1.406% | 4.903% | 18.879% | 1.340% | 637882 | 2224669 | 8566839 | 608107 |
| BGD | LMIC | 0.359% | 6.243% | 17.157% | 9.910% | 601708 | 10451558 | 28725010 | 16591242 |
| BGR | LMIC | 0.171% | 0.406% | 0.969% | 1.166% | 11869 | 28184 | 67187 | 80867 |
| BIH | LMIC | 0.050% | 0.706% | 0.833% | 1.653% | 1670 | 23419 | 27629 | 54848 |
| BLR | LMIC | 0.000% | 0.337% | 0.003% | 0.011% | 7 | 31570 | 269 | 1007 |
| BRA | LMIC | 1.037% | 6.733% | 17.467% | 1.638% | 2210385 | 14355489 | 37238442 | 3492982 |
| CHN | LMIC | 0.176% | 0.957% | 4.325% | 95.306% | 2479778 | 13499895 | 61027992 | |
| COL | LMIC | 0.418% | 0.968% | 1.071% | 23.706% | 212928 | 493094 | 545370 | 12073788 |
| DOM | LMIC | 0.265% | 2.561% | 8.366% | | 29178 | 281744 | 920209 | |
| DZA | LMIC | | | | | | | | |
| ECU | LMIC | 0.775% | 1.905% | 9.681% | 5.577% | 136248 | 335128 | 1702713 | 980880 |
| EGY | LMIC | 0.003% | 0.019% | 0.146% | 0.003% | 2843 | 20830 | 157273 | 3537 |
| IDN | LMIC | 0.997% | 13.880% | 9.184% | 0.078% | 2709561 | 37734907 | 24966554 | 211523 |
| IND | LMIC | 0.063% | 0.484% | 12.100% | 0.758% | 882002 | 6758523 | 168961340 | 10582658 |
| JOR | LMIC | 0.470% | 2.923% | 7.554% | 4.370% | 51374 | 319487 | 825510 | 477576 |
| KAZ | LMIC | 0.000% | 0.000% | 0.000% | 0.000% | | | 8 | 27 |
| LBN | LMIC | 0.015% | 0.158% | 0.377% | 0.474% | 877 | 8922 | 21324 | 26844 |
| LKA | LMIC | 0.001% | | 11.139% | | 225 | | 2441530 | |
| MAR | LMIC | 0.400% | | 1.656% | 0.960% | 146685 | | 607584 | 352127 |
| MEX | LMIC | 0.372% | 5.974% | 6.766% | 6.807% | 469078 | 7526813 | 8524913 | 8576901 |
| MYS | LMIC | 0.082% | 0.342% | 0.351% | 0.287% | 27226 | 113611 | 116412 | 95260 |
| PAK | LMIC | 0.051% | 0.444% | 20.680% | 15.909% | 115767 | 1009212 | 46984317 | 36144293 |
| PER | LMIC | 0.252% | 0.604% | 1.405% | 6.928% | 83969 | 201183 | 468034 | 2307242 |
| PHL | LMIC | 1.129% | 5.820% | 28.247% | 26.855% | 1266742 | 6529696 | 31690301 | 30128741 |
| ROU | LMIC | 0.127% | 0.276% | 1.385% | 0.692% | 24395 | 53109 | 266894 | 133289 |
| RUS | LMIC | 0.005% | 0.173% | 0.266% | | 7924 | 251632 | 386990 | |
| SRB | LMIC | 0.152% | 0.630% | 1.129% | 2.465% | 10455 | 43479 | 77874 | 170048 |
| SVK | LMIC | | | | | | | | |
| SVN | LMIC | 0.066% | 0.058% | 0.183% | 0.168% | 1394 | 1217 | 3851 | 3526 |
| THA | LMIC | 0.001% | 0.396% | 0.520% | 0.006% | 677 | 282900 | 371691 | 4080 |
| TUN | LMIC | 0.138% | 0.905% | | 1.412% | 16766 | 110072 | | 171688 |
| TUR | LMIC | 0.068% | 0.233% | 0.405% | 0.151% | 57004 | 194632 | 338043 | 125622 |
| TWN | LMIC | | | | | | | | |
| VEN | LMIC | 0.972% | 4.934% | 6.691% | | 281691 | 1429401 | 1938449 | |
| VNM | LMIC | 0.105% | 0.807% | 5.559% | 4.847% | 101257 | 780381 | 5372227 | 4684807 |
| ZAF | LMIC | 0.706% | 2.307% | 7.526% | 2.438% | 415242 | 1356464 | 4425529 | 1433411 |
| **Median** | | 0.15% | 0.71% | 2.99% | 1.41% | 70487 | 301194 | 716547 | 281825 |
| **p25** | | 0.05% | 0.34% | 0.49% | 0.29% | 9822 | 67350 | 147058 | 74362 |

| | | | | | | | | |
|---|---|---|---|---|---|---|---|---|
| **p75** | 0.42% | 2.74% | 9.31% | 5.58% | 428701 | 2025852 | 8535395 | 3790938 |
| **IQR** | 0.37% | 2.40% | 8.82% | 5.29% | 418879 | 1958502 | 8388337 | 3716576 |
| **Average** | 0.33% | 2.16% | 6.31% | 7.45% | 406088 | 3548374 | 13680260 | 4625604 |
| **Total** | | | | | 12994807 | 106451221 | 437768308 | 129516921 |

Notes: Group 1 is for ceftriaxone +Amikacin/Gentamicin, Group 2 is for Piperazillin+Tazobactam, Group 3 is for Meropenem and Group 4 is for Colistin. Countries with missing data either did not report the highlighted antibiotics or national poverty lines were not found. Table A1 indicates the abbreviations of country-ISO3-codes. Authors' analysis of IQVIA MIDAS Quarterly Sales data for the year 2019, reflecting estimates of real-world activity. Copyright IQVIA. All Rights Reserved.

**Text A11.** Approximation for ex-factory-to-retail prices across AWaRe categories

We approximated end-buyer prices paid in the retail sector. As IQVIA MIDAS Quarterly data provides ex-manufacturer (ex-manufacturer) prices only, we estimated the ratio between volume-weighted retail (end-buyer) and ex-manufacturer prices per DDD across AWaRe categories using Pharma 14 data,[11] among 28 available countries, most of which were HIC (71%, n=20, see list of countries below). Pharma14 is a global pharmaceutical database launched in 2016, tracking pricing and consumption patterns across multiple countries. It provides data on prescription and over-the-counter medicines, covering ex-factory, wholesale, and retail prices, while also offering insights into consumption trends, medicine shortages, and European parallel trade. Next, we computed ex-factory-to-retail ratios at the country-level, whenever available, and per HIC- or MIC-aggregated values if country-specific ratios were not available. After doing the calculations, retail antibiotic prices averaged 1·03-1·30 times higher prices than ex-factory (ex-manufacturer) prices (Figure Text A11.1, Table Text A11.1), with differences being larger in MICs (1·21-times greater, compared to HICs, 1·12).

<u>List of countries included in Pharma14</u> (n=28): AUS – Australia; AUT – Austria; BEL - Belgium; BGR - Bulgaria; BRA - Brazil; CAN - Canada; CZE - Czech Republic; DEU - Germany; EST - Estonia; FIN - Finland; GBR - United Kingdom; GRC - Greece; HRV - Croatia; HUN - Hungary; IRL - Ireland; LTU - Lithuania; LUX - Luxembourg; LVA - Latvia; MEX - Mexico; POL - Poland; ROU - Romania; RUS - Russia; SAU - Saudi Arabia; SVK - Slovakia; SVN - Slovenia; SWE - Sweden; TUR - Turkey; ZAF - South Africa.

**Figure Text A11.1.** Comparison between ex-factory (ex-manufacturer) and retail prices per DDD among available antibiotics on Pharma14

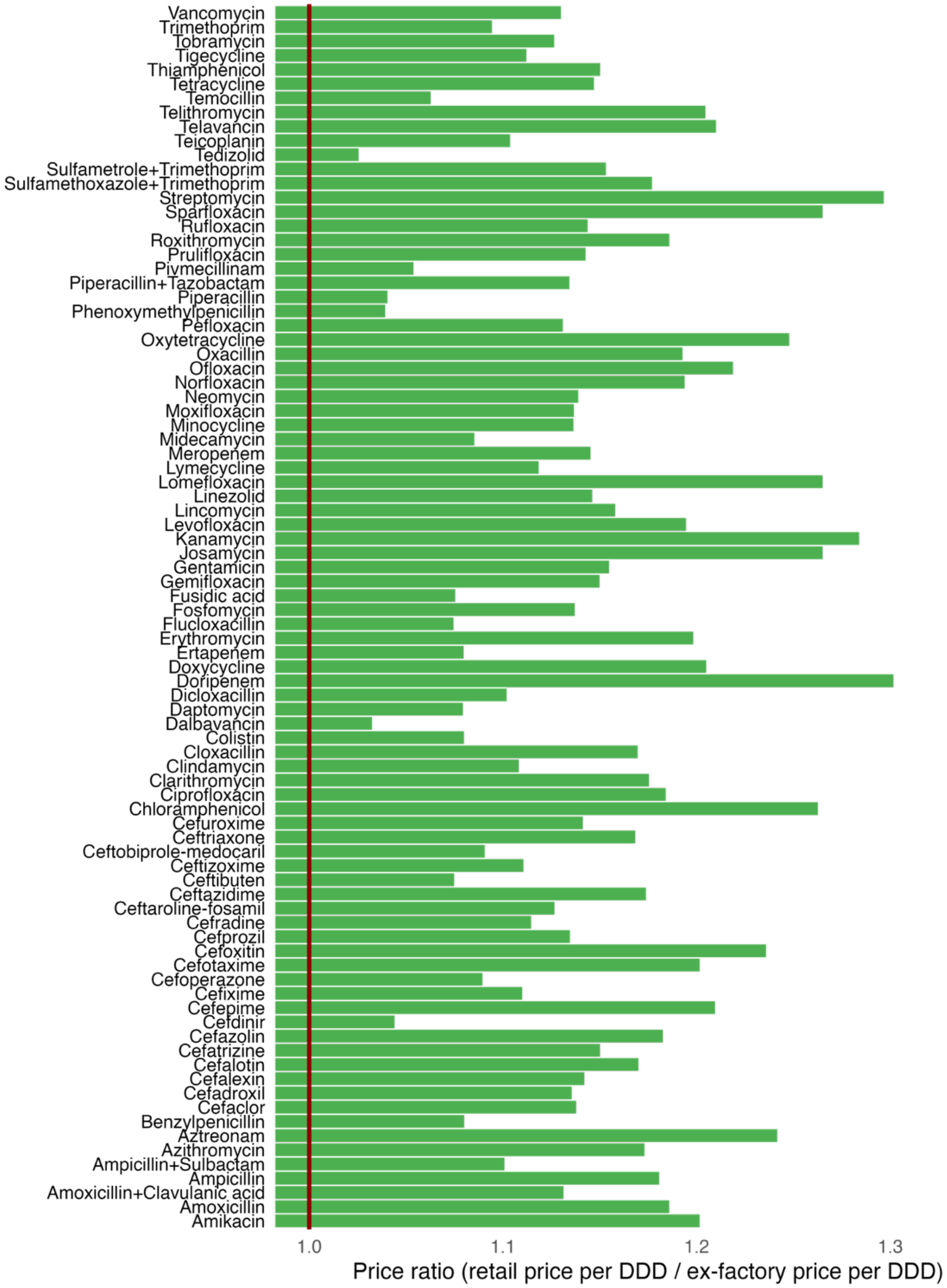

Notes: DDD= Daily defined doses. Pharma14.[11]

**Table Text A11.1.** Ex-factory to retail price ratio using Pharma 14, by WHO region, income group and country

| WHO region | Median | p25th | p75th |
|---|---|---|---|
| AFR | 1.295 | 1.270 | 1.315 |
| AMR | 1.226 | 1.252 | 1.253 |
| EUR | 1.147 | 1.063 | 1.210 |
| WPR | 1.075 | 1.074 | 1.075 |
| **WB income group** | **Median** | **p25th** | **p75th** |
| HIC | 1.120 | 1.060 | 1.150 |
| LMIC | 1.210 | 1.140 | 1.265 |
| **Country ISO-3 code** | **Median** | **p25th** | **p75th** |
| AUS | 1.075 | 1.074 | 1.075 |
| AUT | 1.135 | 1.125 | 1.154 |
| BEL | 1.148 | 1.149 | 1.150 |
| BGR | 1.058 | 1.046 | 1.069 |
| BRA | 1.252 | 1.253 | 1.253 |
| CAN | 1.080 | 1.080 | 1.080 |
| CZE | 1.050 | 1.050 | 1.050 |
| DEU | 1.166 | 1.046 | 1.221 |
| EST | 1.131 | 1.100 | 1.151 |
| FIN | 1.040 | 1.040 | 1.040 |
| GBR | 1.072 | 1.062 | 1.065 |
| GRC | 1.150 | 1.149 | 1.150 |
| HRV | 1.055 | 1.040 | 1.060 |
| HUN | 1.051 | 1.044 | 1.053 |
| IRL | 1.080 | 1.080 | 1.080 |
| LTU | 1.084 | 1.069 | 1.091 |
| LUX | 1.164 | 1.149 | 1.167 |
| LVA | 1.241 | 1.207 | 1.265 |
| MEX | 1.153 | 1.150 | 1.160 |
| POL | 1.083 | 1.050 | 1.120 |
| ROU | 1.132 | 1.138 | 1.140 |
| RUS | 1.252 | 1.210 | 1.265 |
| SAU | 1.145 | 1.150 | 1.150 |
| SVK | 1.094 | 1.075 | 1.110 |
| SVN | 1.060 | 1.050 | 1.065 |
| SWE | 1.027 | 1.027 | 1.027 |
| TUR | 1.071 | 1.070 | 1.081 |
| ZAF | 1.295 | 1.270 | 1.315 |

Notes: Notes: WHO= World Health Organization. AFRO= African region, AMRO= Americas region, EMRO= East Mediterranean region, EURO= Europe, SEARO= Southeast Asia region, WPRO= Western-pacific region. WB= World Bank. HIC= high-income countries. LMIC= Low- and middle-income countries (here it is just middle-income countries as there is no low-income countries). P25 and 75 mean percentiles. Table A1 indicates the abbreviations of country-ISO3-codes. Pharma14.[11]

### STROBE guidelines

STROBE Statement[12]—checklist of items that should be included in reports of observational studies.

| | Item No | Recommendation | Page No |
|---|---|---|---|
| **Title and abstract** | 1 | (*a*) Indicate the study's design with a commonly used term in the title or the abstract | 1-2 |
| | | (*b*) Provide in the abstract an informative and balanced summary of what was done and what was found | 2-3 |
| **Introduction** | | | |
| Background/rationale | 2 | Explain the scientific background and rationale for the investigation being reported | 4 |
| Objectives | 3 | State specific objectives, including any prespecified hypotheses | 4 |
| **Methods** | | | |
| Study design | 4 | Present key elements of study design early in the paper | 5 |
| Setting | 5 | Describe the setting, locations, and relevant dates, including periods of recruitment, exposure, follow-up, and data collection | 5 |
| Participants | 6 | (*a*) *Cohort study*—Give the eligibility criteria, and the sources and methods of selection of participants. Describe methods of follow-up<br>*Case-control study*—Give the eligibility criteria, and the sources and methods of case ascertainment and control selection. Give the rationale for the choice of cases and controls<br>*Cross-sectional study*—Give the eligibility criteria, and the sources and methods of selection of participants | |
| | | (*b*) *Cohort study*—For matched studies, give matching criteria and number of exposed and unexposed<br>*Case-control study*—For matched studies, give matching criteria and the number of controls per case | |
| Variables | 7 | Clearly define all outcomes, exposures, predictors, potential confounders, and effect modifiers. Give diagnostic criteria, if applicable | 5-6 |
| Data sources/ measurement | 8* | For each variable of interest, give sources of data and details of methods of assessment (measurement). Describe comparability of assessment methods if there is more than one group | *5-6* |
| Bias | 9 | Describe any efforts to address potential sources of bias | 6 |
| Study size | 10 | Explain how the study size was arrived at | 6 |
| Quantitative variables | 11 | Explain how quantitative variables were handled in the analyses. If applicable, describe which groupings were chosen and why | 6 |
| Statistical methods | 12 | (*a*) Describe all statistical methods, including those used to control for confounding | 6 |
| | | (*b*) Describe any methods used to examine subgroups and interactions | 6 |
| | | (*c*) Explain how missing data were addressed | 6 |
| | | (*d*) *Cohort study*—If applicable, explain how loss to follow-up was addressed<br>*Case-control study*—If applicable, explain how matching of cases and controls was addressed<br>*Cross-sectional study*—If applicable, describe analytical methods taking account of sampling strategy | 6 |
| | | (*e*) Describe any sensitivity analyses | 6 |

Continued on next page

| | | | |
|---|---|---|---|
| **Results** | | | |
| Participants | 13* | (a) Report numbers of individuals at each stage of study—eg numbers potentially eligible, examined for eligibility, confirmed eligible, included in the study, completing follow-up, and analysed | 7 |
| | | (b) Give reasons for non-participation at each stage | |
| | | (c) Consider use of a flow diagram | |
| Descriptive data | 14* | (a) Give characteristics of study participants (eg demographic, clinical, social) and information on exposures and potential confounders | |
| | | (b) Indicate number of participants with missing data for each variable of interest | 5-7 |
| | | (c) *Cohort study*—Summarise follow-up time (eg, average and total amount) | |
| Outcome data | 15* | *Cohort study*—Report numbers of outcome events or summary measures over time | |
| | | *Case-control study*—Report numbers in each exposure category, or summary measures of exposure | |
| | | *Cross-sectional study*—Report numbers of outcome events or summary measures | *7-9* |
| Main results | 16 | (*a*) Give unadjusted estimates and, if applicable, confounder-adjusted estimates and their precision (eg, 95% confidence interval). Make clear which confounders were adjusted for and why they were included | |
| | | (*b*) Report category boundaries when continuous variables were categorized | |
| | | (*c*) If relevant, consider translating estimates of relative risk into absolute risk for a meaningful time period | |
| Other analyses | 17 | Report other analyses done—eg analyses of subgroups and interactions, and sensitivity analyses | 8-9 |
| **Discussion** | | | |
| Key results | 18 | Summarise key results with reference to study objectives | 10 |
| Limitations | 19 | Discuss limitations of the study, taking into account sources of potential bias or imprecision. Discuss both direction and magnitude of any potential bias | 11 |
| Interpretation | 20 | Give a cautious overall interpretation of results considering objectives, limitations, multiplicity of analyses, results from similar studies, and other relevant evidence | 10-11 |
| Generalisability | 21 | Discuss the generalisability (external validity) of the study results | 11 |
| **Other information** | | | |
| Funding | 22 | Give the source of funding and the role of the funders for the present study and, if applicable, for the original study on which the present article is based | 12 |

*Give information separately for cases and controls in case-control studies and, if applicable, for exposed and unexposed groups in cohort and cross-sectional studies.

**Note:** An Explanation and Elaboration article discusses each checklist item and gives methodological background and published examples of transparent reporting. The STROBE checklist is best used in conjunction with this article (freely available on the Web sites of PLoS Medicine at http://www.plosmedicine.org/, Annals of Internal Medicine at http://www.annals.org/, and Epidemiology at http://www.epidem.com/). Information on the STROBE Initiative is available at www.strobe-statement.org.
